## Supplementary Figures and Tables for "Adjuvant nivolumab, capecitabine or the combination in patients with residual triple-negative breast cancer: the OXEL randomized phase II study": Supplementary Figures and Tables.pdf

#### Supplementary Figures, Tables, Protocol and References

##### Supplementary Fig. 1

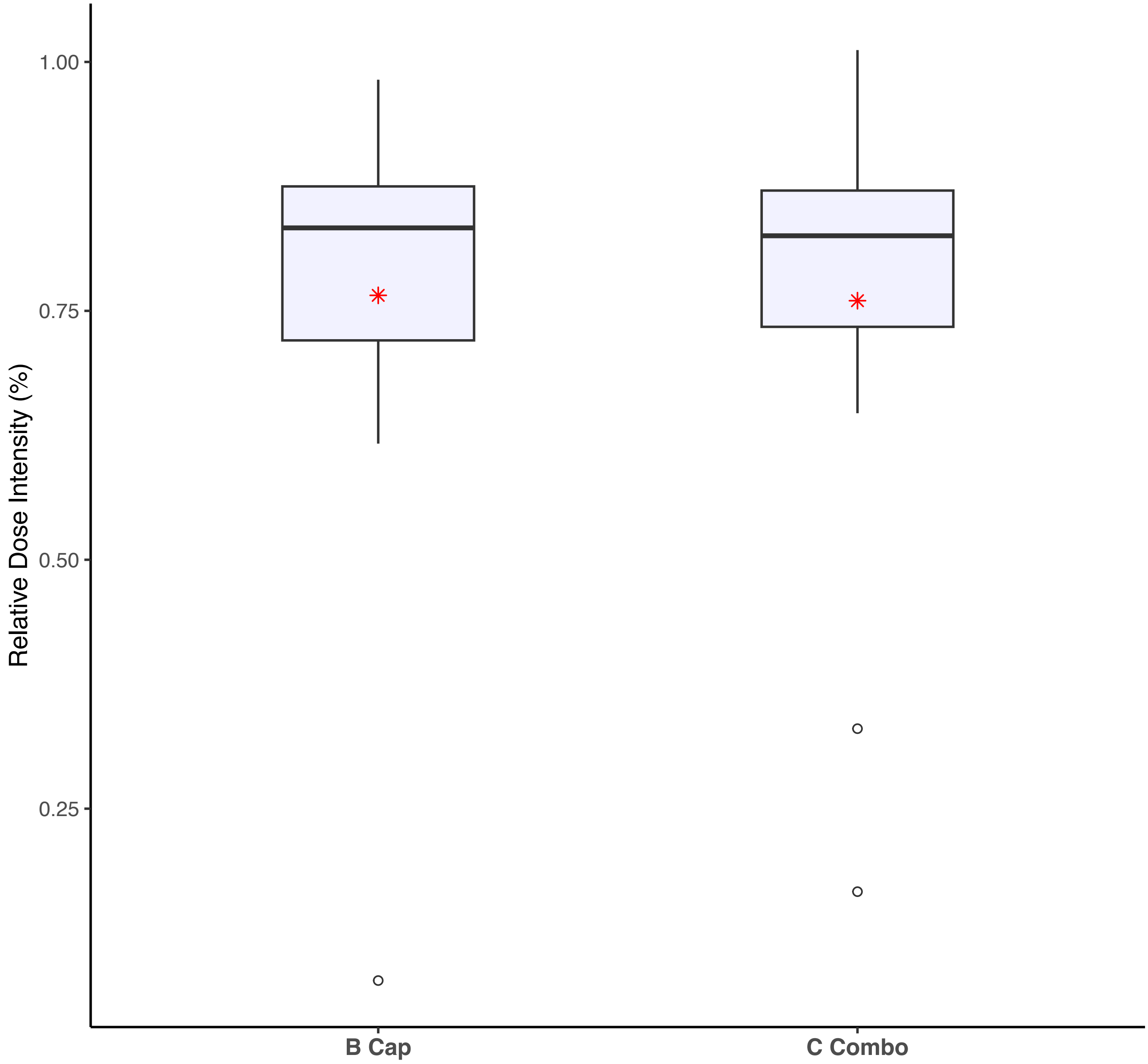

**Supplementary Figure 1. Boxplot of capecitabine relative dose intensity among all 30 patients who received capecitabine (Arm B, n = 15 patients; Arm C, n = 15 patients).** The red asterisk (*) represents the mean value. Source data are provided as a source data file.

### Supplementary Fig. 2

a

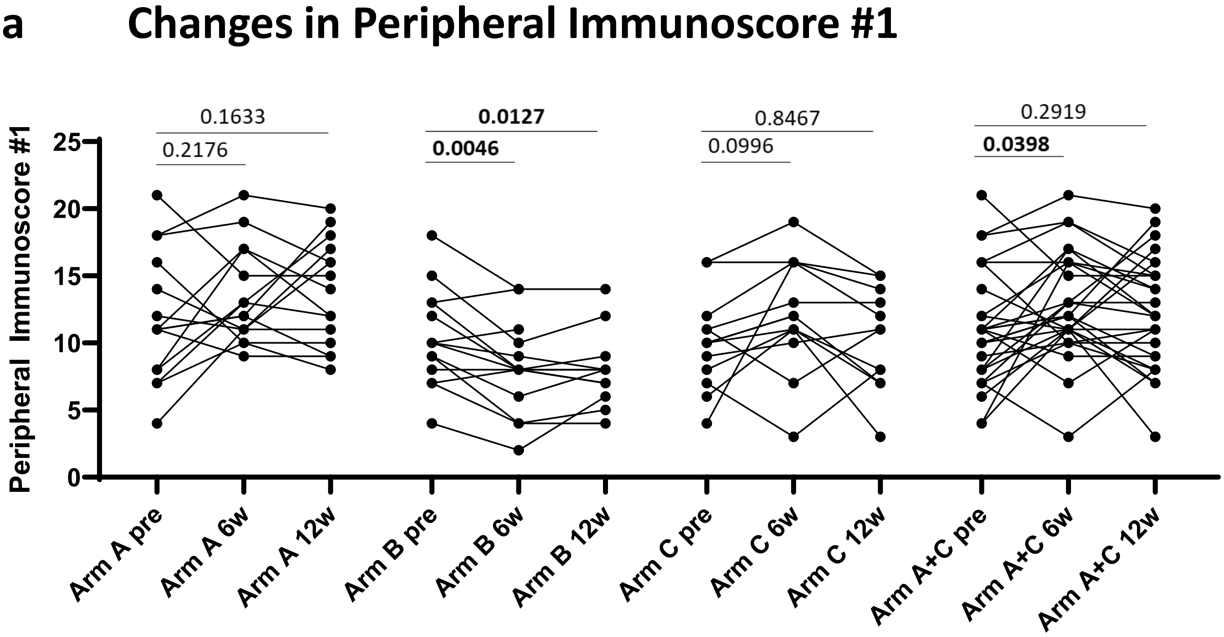

b

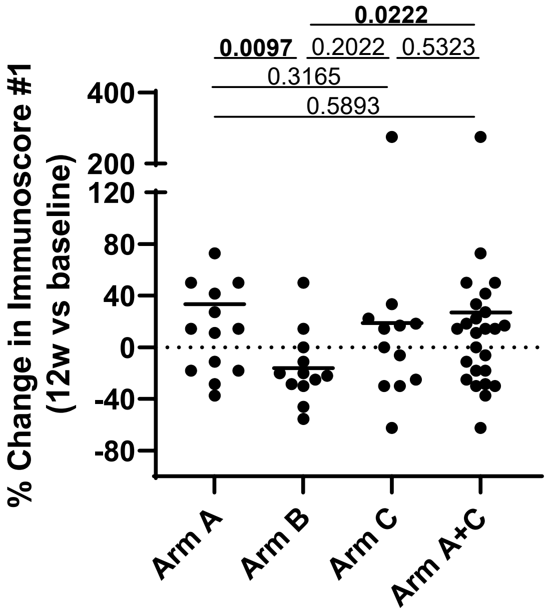

**Supplementary Fig. 2. Changes in peripheral immunoscore #1 after 6 and 12 weeks of therapy vs. landmark.** (**A**) The peripheral immunoscore based on refined PBMC subsets of cell types reflecting known function (Supplementary Table 3, Immunoscore #1) before and after 6 and 12 weeks of therapy in Arm A (n=15), Arm B (n=14), Arm C (n=13), and Arms A and C combined (n=28). The peripheral immunoscore is the sum of points assigned to each subset based on tertile distribution as previously described [1]. (**B**) Comparison of the percent change after 12 weeks vs. baseline in the peripheral immunoscore in in Arm A (n=15), Arm B (n=14), Arm C (n=13), and Arms A and C combined (n=28). *p* values are shown; *p* values were calculated by a 2 tailed Wilcoxon test in A and a 2 tailed Mann-Whitney test in B. Source data are provided as a source data file.

### Supplementary Fig. 3

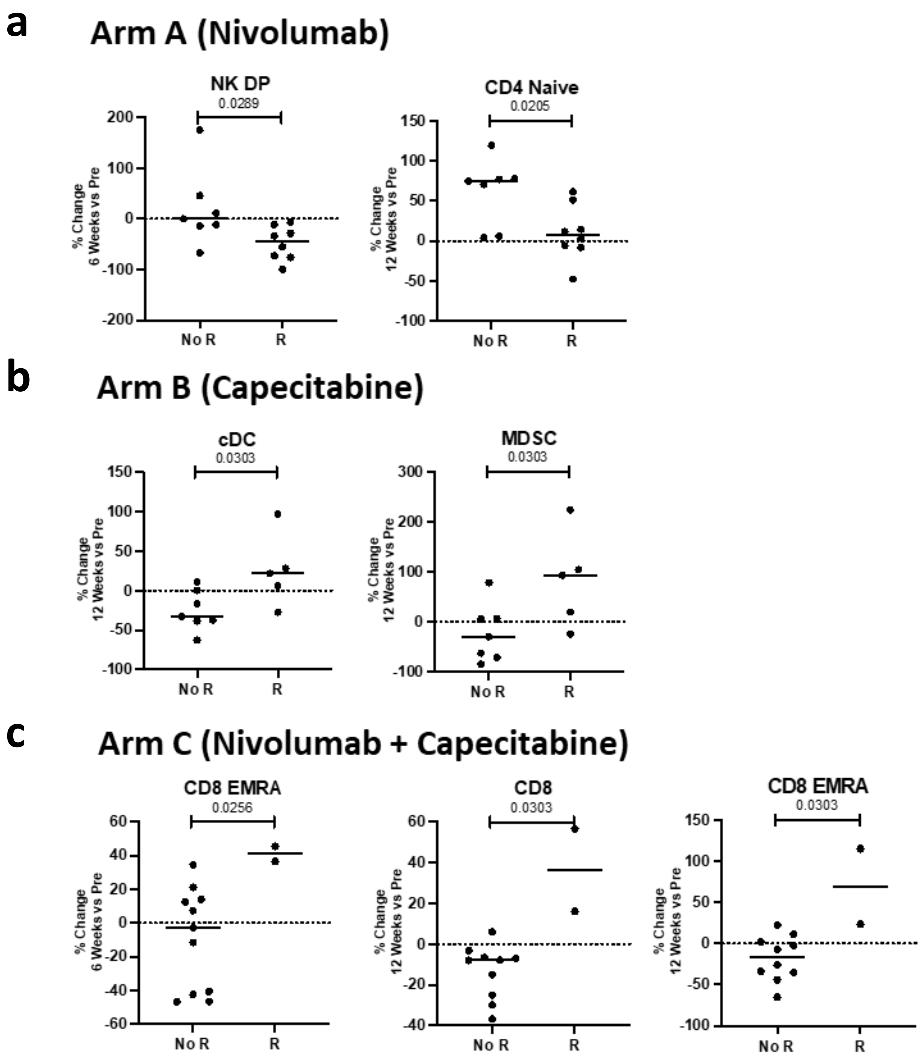

**Supplementary Fig. 3. Changes in the peripheral immune profile after 6 and 12 weeks of therapy vs. landmark associated with the development of recurrence (R).** The percent change in peripheral blood mononuclear cell (PBMC) subsets after 6 and 12 weeks of treatment in each arm in patients with disease recurrence (R) and in patients without disease recurrence (no R). Comparisons were made in patients treated with (A) nivolumab (n=8 R, n=7 NR at 6 and 12 weeks), (B) capecitabine (n=5 R, n=7 NR at 6 at 12 weeks), and (C) nivolumab plus capecitabine (n=2 R and n=11 NR at 6 weeks and n=2 R and n=10 NR at 12 weeks). Changes in 10 classic PBMC cell types and 148 refined PBMC subsets reflective of maturation and function were analyzed. Notable subsets with significant differences displayed and include those with $p < 0.05$ (calculated by a 2 tailed Mann-Whitney test), and a difference in medians > 0.05% of PBMCs. No adjustment was made for multiple comparisons. Source data are provided as a source data file.

### Supplementary Fig. 4

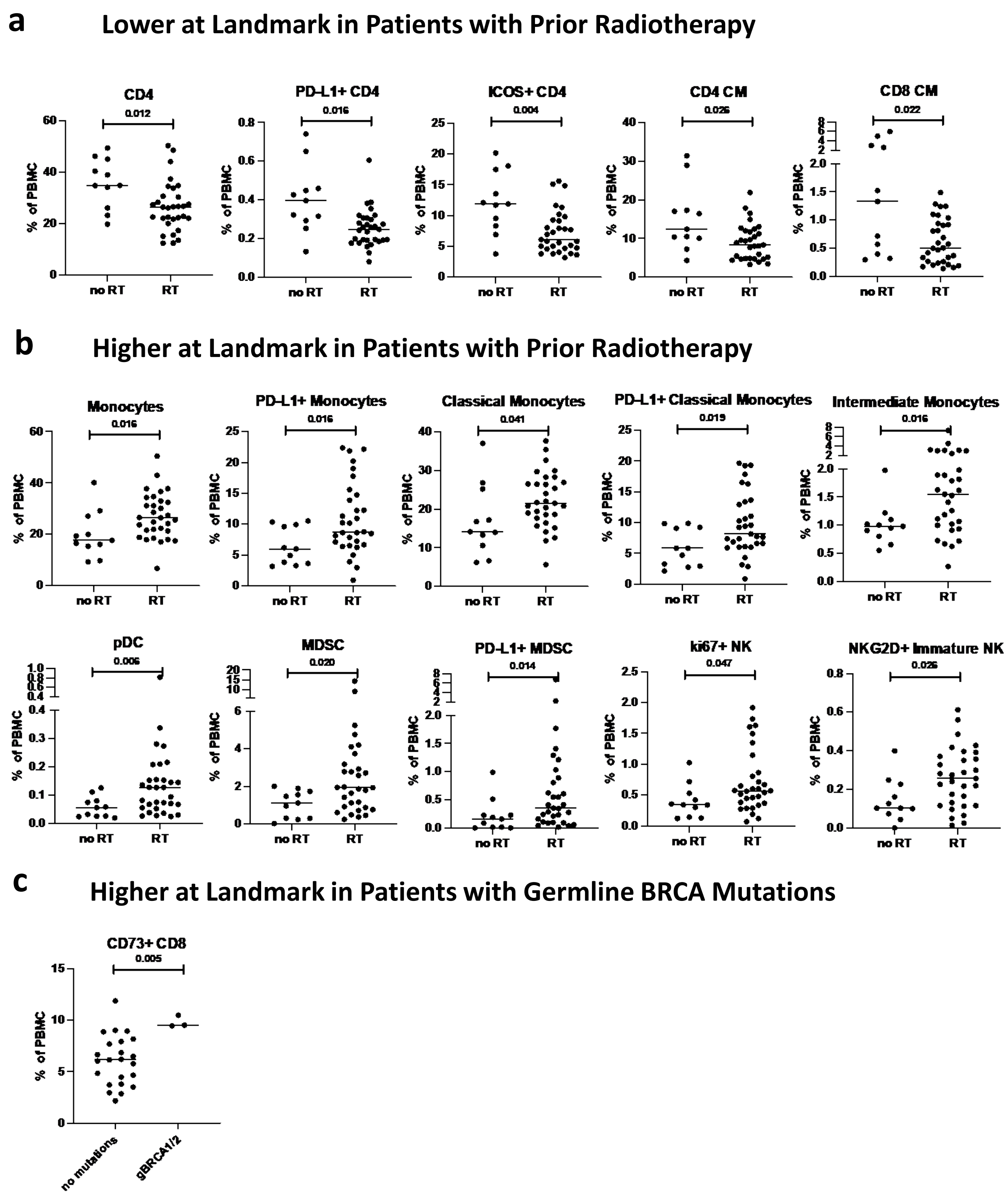

**Supplementary Fig. 4. Association between prior radiotherapy (RT) and *BRCA1/2* mutation status and the landmark peripheral immune profile.** The peripheral immune profile was compared at landmark and evaluated for subsets that were lower (**A**) or higher (**B**) as percent of PBMC in patients with prior radiotherapy (RT, n=31) compared to patients without prior RT (no RT, n=11). Landmark differences in the peripheral immune profile were analyzed in patients with germline *BRCA1* or *BRCA2* (*gBRCA1/2*) mutations (n=3) and compared to those with no deleterious mutations (n=23) (**C**). Analyses were performed in all arms combined, and differences were analyzed in 10 classic PBMC cell types and 148 refined PBMC subsets reflective of maturation and function. Notable subsets with significant differences are displayed and include those with $p < 0.05$ (calculated by a 2 tailed Mann-Whitney test), and a difference in medians > 0.05 of PBMCs. No adjustment was made for multiple comparisons. Source data are provided as a source data file.

### Supplementary Fig. 5

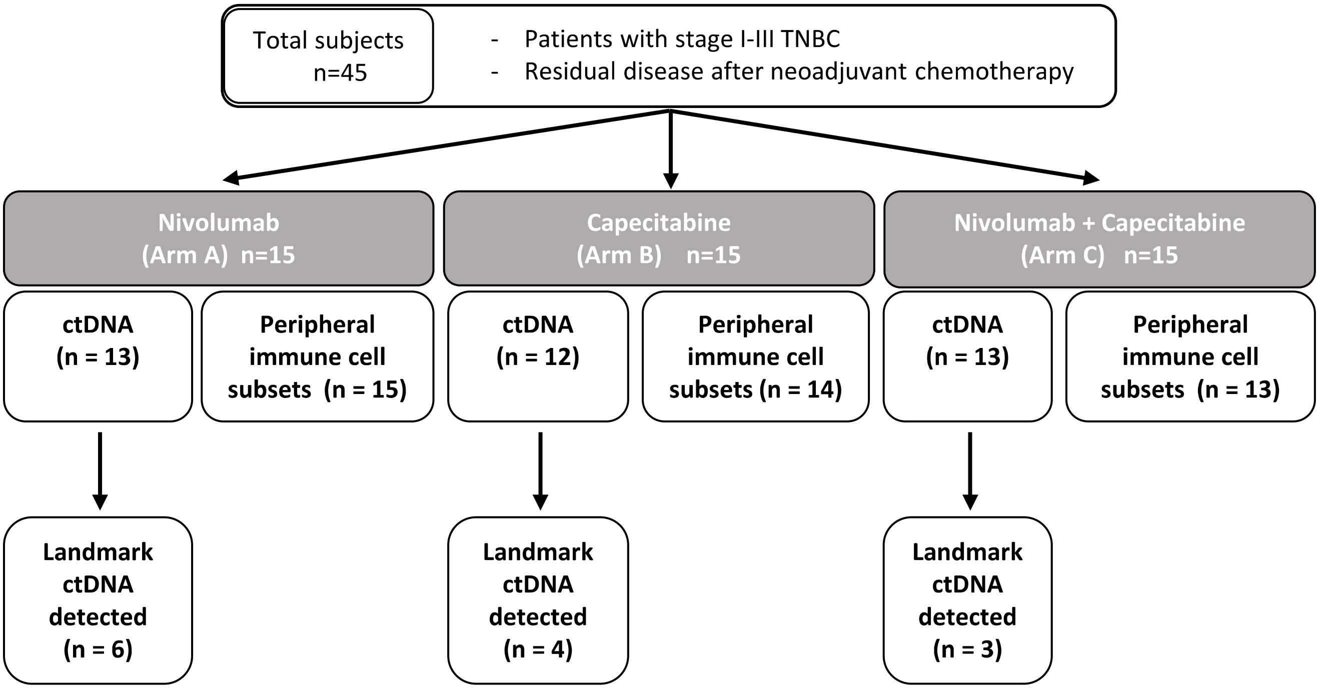

Consort diagram of number of patients in each arm with available landmark circulating tumor DNA (ctDNA) results and detected ctDNA at landmark.

Source data are provided as a source data file.

### Supplementary Fig. 6

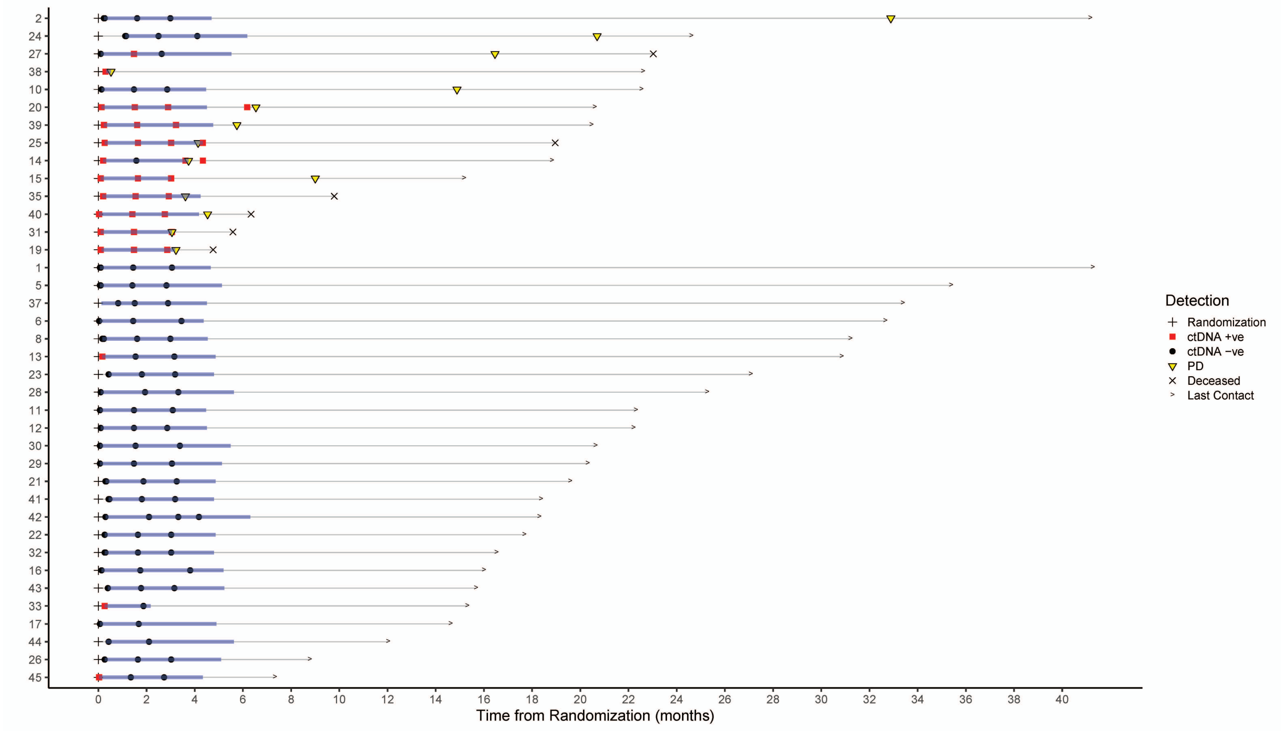

**Supplementary Figure 6. Swimmer plot depicting patients' disease courses, including recurrence, ctDNA assay samples and results among all 38 patients who underwent successful ctDNA testing.** Source data are provided as a source data file.

### Supplementary Fig. 7

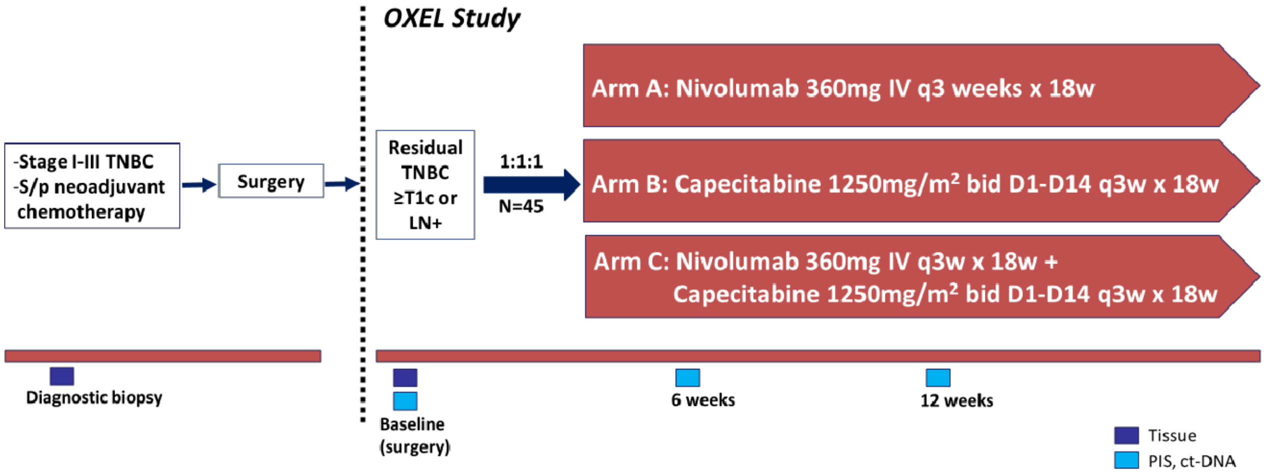

**OXEL Study Design**

### Supplementary Fig. 8

**a**

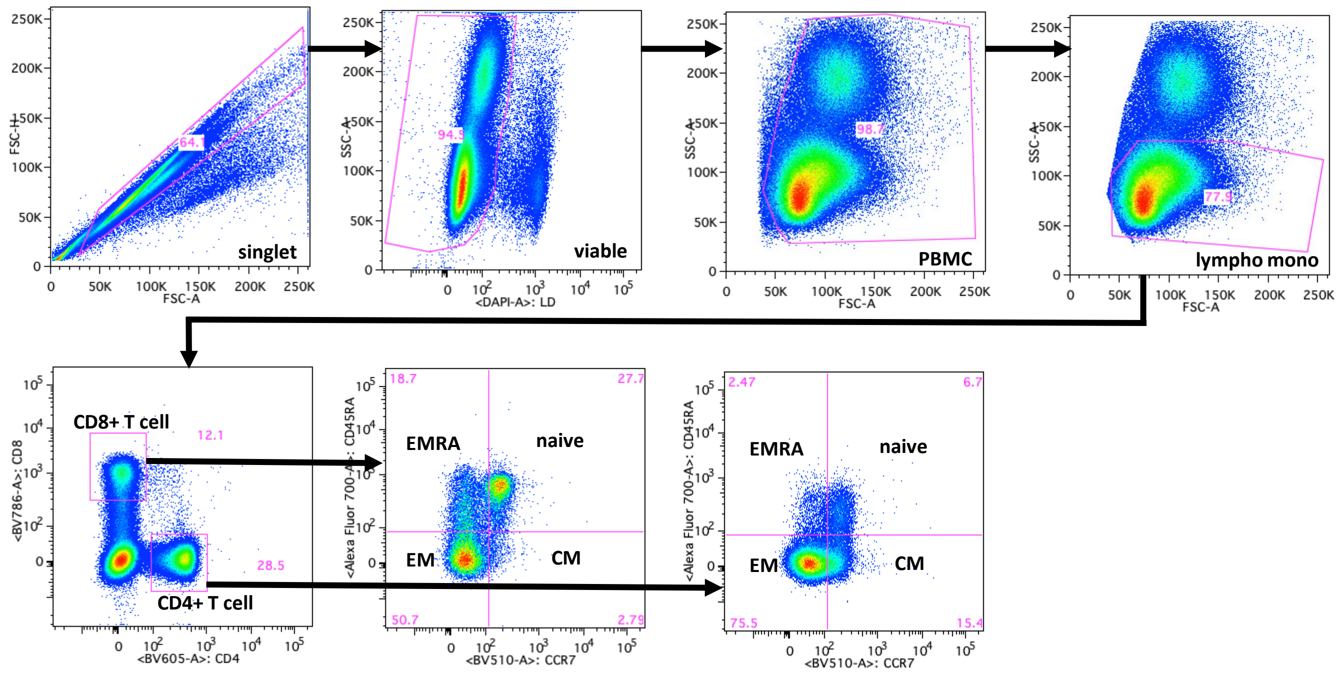

**Markers in Total and Memory CD4**

CTLA4, 41bb, PD-1, PD-L1, Tim3, ki67, CD73

**Markers in Total and Memory CD8**

CTLA4, 41bb, PD-1, PD-L1, Tim3, ki67, CD73

**b**

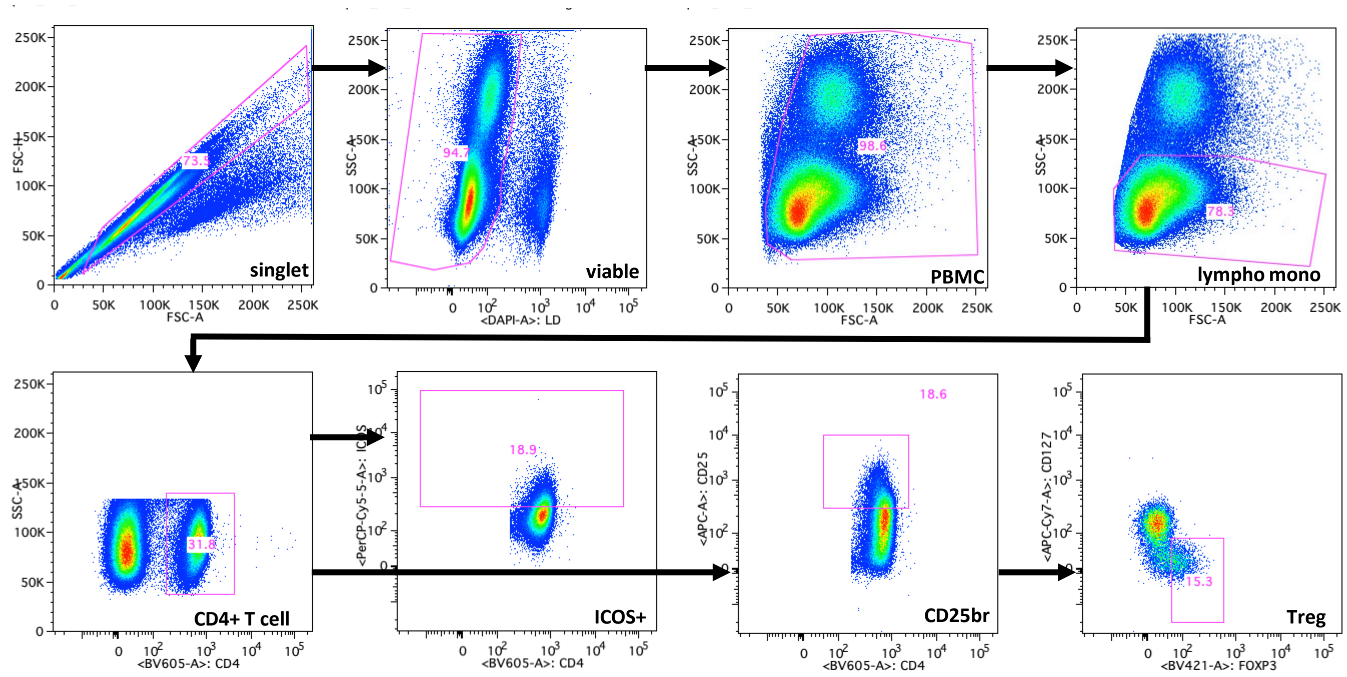

**Markers in ICOS+ CD4**

PD-1, PD-L1

**Markers in Treg**

CTLA4, PD-1, PD-L1, CD38, CD45RA, CD49d, HLA DR, ICOS, ki67

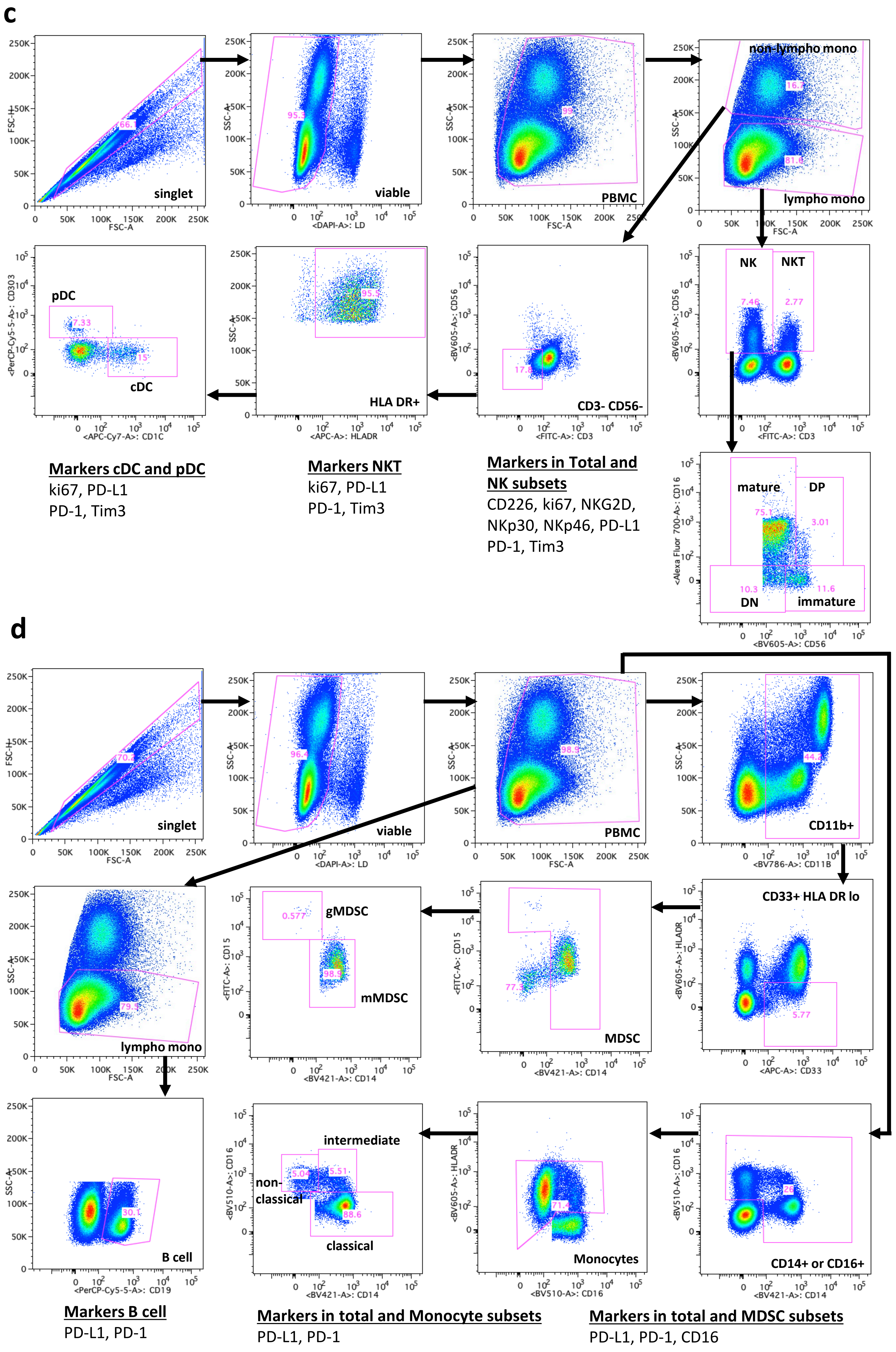
c
singlet
viable
PBMC
non-lympho mono
lympho mono
pDC
cDC
HLA DR+
CD3- CD56-
NK
NKT
mature
DP
DN
immature
Markers cDC and pDC
ki67, PD-L1
PD-1, Tim3
Markers NKT
ki67, PD-L1
PD-1, Tim3
Markers in Total and NK subsets
CD226, ki67, NKG2D, NKp30, NKp46, PD-L1
PD-1, Tim3
d
singlet
viable
PBMC
CD11b+
lympho mono
gMDSC
mMDSC
MDSC
CD33+ HLA DR lo
B cell
intermediate
non-classical
classical
Monocytes
CD14+ or CD16+
Markers B cell
PD-L1, PD-1
Markers in total and Monocyte subsets
PD-L1, PD-1
Markers in total and MDSC subsets
PD-L1, PD-1, CD16

**Supplementary Fig. 8. Gating strategy to identity 158 peripheral immune cell subsets. Four representative immune flow cytometry panels using PBMC from a single participant are shown.** Classic immune cell types included CD4$^+$ T cells and CD8$^+$ T cells (a), Tregs (b), natural killer (NK) cells, NK-T cells, conventional dendritic cells (cDCs), and plasmacytoid DCs (pDCs) (c), and myeloid derived suppressor cells (MDSCs), monocytes, and B cells (d). Refined immune subsets were determined within parental classic immune cell types using the markers indicated.

| Supplementary Table 1. Capecitabine dose intensity for patients in Arm B and C. | | | |
|---|---|---|---|
| **Variable** | **Overall (N=30)** | **Arm B (N=15)** | **Arm C (N=15)** |
| RDI (mean (SD), median [IQR], [min, max]) | 0.76 (0.22), 0.83 [0.71 - 0.88], [0.08, 1.01] | 0.77 (0.21), 0.83 [0.72 - 0.88], [0.08, 0.98] | 0.76 (0.23), 0.83 [0.73 - 0.87], [0.17, 1.01] |
| Cumulative dose of capecitabine (mean (SD), median [IQR], [min, max]) | 272600 (81378.51), 286750 [261000 - 316000], [27950, 408000] | 269966.67 (75282.78), 282000 [263050 - 299250], [27950, 354000] | 275233.33 (89641.52), 291500 [262000 - 324000], [70000, 408000] |
| Cycle 1 | 57186.67 (10427.71), 56000 [54000 - 66650], [27950, 77000] | 53593.33 (10113.83), 56000 [50800 - 56000], [27950, 70000] | 60780 (9761.31), 56000 [56000 - 68750], [39000, 77000] |
| Cycle 2 | 50043.33 (15591.33), 56000 [48250 - 56000], [0, 70000] | 49773.33 (14309.81), 54000 [50800 - 56000], [0, 56000] | 50313.33 (17280.50), 56000 [42750 - 56000], [0, 70000] |
| Cycle 3 | 44153.33 (15350.38), 43000 [42000 - 55425], [0, 67200] | 45226.67 (14823.46), 47500 [42000 - 55950], [0, 60000] | 43080 (16306.32), 42000 [42000 - 54000], [0, 67200] |
| Cycle 4 | 40863.33 (17335.20), 42000 [42000 - 55500], [0, 67200] | 41013.33 (16110.42), 42000 [42000 - 51000], [0, 60200] | 40713.33 (19049.67), 42000 [42000 - 55000], [0, 67200] |
| Cycle 5 | 41940 (16726.12), 42000 [42000 - 55500], [0, 72000] | 41746.67 (13385.17), 42000 [42000 - 44000], [0, 60200] | 42133.33 (20006.67), 42000 [42000 - 56000], [0, 72000] |
| Cycle 6 | 38413.33 (18971.26), 42000 [36750 - 51000], [0, 67200] | 38613.33 (17542.19), 42000 [38500 - 45000], [0, 60200] | 38213.33 (20921.55), 42000 [35000 - 54000], [0, 67200] |

Abbreviations: RDI, relative dose intensity; SD, standard deviation; IQR, interquartile range

**Supplementary Table 2.** 158 peripheral immune cell subsets analyzed by flow cytometry. Ten parental phenotypes are identified as well as refined subsets of each relating to maturation and function. Expected function based on expression of specific markers within each subset is indicated in italics.

#### 1. Total CD4$^+$ T cells

- PD-L1$^+$ CD4 - *activation/inhibition*
- PD-1$^+$ CD4 - *activation/inhibition*
- CTLA-4$^+$ CD4 - *inhibition*
- Tim-3$^+$ CD4 - *inhibition*
- 41bb$^+$ CD4 - *co-stimulation*
- Ki67$^+$ CD4 - *proliferation*
- CD73$^+$ CD4 - exhausted/suppressive
- ICOS$^+$ CD4 - activation
  - ICOS$^+$ PD-L1$^+$ CD4 - activation/inhibition
  - ICOS$^+$ PD-1$^+$ CD4 - activation/inhibition
- Naïve (CCR7$^+$CD45RA$^+$) CD4
- Central memory (CCR7$^+$ CD45RA$^-$) CD4
  - PD-L1$^+$ CM CD4 - activation/inhibition
  - PD-1$^+$ CM CD4 - activation/inhibition
  - CTLA-4$^+$ CM CD4 - inhibition
  - Tim-3$^+$ CM CD4 - inhibition
  - 41bb$^+$ CM CD4 - co-stimulation
  - Ki67$^+$ CM CD4 - proliferation
  - CD73$^+$ CM CD4 - exhausted/suppressive
- Effector memory (CCR7$^-$ CD45RA$^-$) CD4
  - PD-L1$^+$ EM CD4 - activation/inhibition
  - PD-1$^+$ EM CD4 - activation/inhibition
  - CTLA-4$^+$ EM CD4 - inhibition
  - Tim-3$^+$ EM CD4 - inhibition
  - 41bb$^+$ EM CD4 - co-stimulation
  - Ki67$^+$ EM CD4 - proliferation
  - CD73$^+$ EM CD4 - exhausted/suppressive
- EMRA (CCR7$^-$CD45RA$^+$) CD4
  - PD-L1$^+$ EMRA CD4 - activation/inhibition
  - PD-1$^+$ EMRA CD4 - activation/inhibition
  - CTLA-4$^+$ EMRA CD4 - inhibition
  - Tim-3$^+$ EMRA CD4 - inhibition
  - 41bb$^+$ EMRA CD4 - co-stimulation
  - Ki67$^+$ EMRA CD4 - proliferation
  - CD73$^+$ EMRA CD4 - exhausted/suppressive

#### 2. Total CD8$^+$ T cells

- PD-L1$^+$ CD8 - activation/inhibition
- PD-1$^+$ CD8 - activation/inhibition
- CTLA-4$^+$ CD8 - inhibition
- Tim-3$^+$ CD8 - inhibition
- 41bb$^+$ CD8 - co-stimulation
- Ki67$^+$ CD8 - proliferation
- CD73$^+$ CD8 - exhausted/suppressive
- Naïve (CCR7$^+$CD45RA$^+$) CD8
- Central memory (CCR7$^+$CD45RA$^-$) CD8
  - PD-L1$^+$ CM CD8 - activation/inhibition
  - PD-1$^+$ CM CD8 - activation/inhibition
  - CTLA-4$^+$ CM CD8 - inhibition
  - Tim-3$^+$ CM CD8 - inhibition
  - 41bb$^+$ CM CD8 - co-stimulation
  - Ki67$^+$ CM CD8 - proliferation
  - CD73$^+$ CM CD8 - exhausted/suppressive
- Effector memory (CCR7$^-$ CD45RA$^-$) CD8
  - PD-L1$^+$ EM CD8 - activation/inhibition
  - PD-1$^+$ EM CD8 - activation/inhibition
  - CTLA-4$^+$ EM CD8 - inhibition
  - Tim-3$^+$ EM CD8 - inhibition
  - 41bb$^+$ EM CD8 - co-stimulation
  - Ki67$^+$ EM CD8 - proliferation
  - CD73$^+$ EM CD8 - exhausted/suppressive
- EMRA (CCR7$^-$ CD45RA$^+$) CD8
  - PD-L1$^+$ EMRA CD8 - activation/inhibition
  - PD-1$^+$ EMRA CD8 - activation/inhibition
  - CTLA-4$^+$ EMRA CD8 - inhibition
  - Tim-3$^+$ EMRA CD8 - inhibition
  - 41bb$^+$ EMRA CD8 - co-stimulation
  - Ki67$^+$ EMRA CD8 - proliferation
  - CD73$^+$ EMRA CD8 - exhausted/suppressive

#### 3. Total Tregs

- PD-L1$^+$ Tregs - activation/inhibition
- PD-1$^+$ Tregs - suppression
- CTLA-4$^+$ Tregs - suppression
- ICOS$^+$ Tregs - suppression
- CD45RA$^+$ Tregs - highly expandable in vitro
- CD49d$^-$ Tregs - suppression
- Ki67$^+$ Tregs - proliferation
- CD38$^+$ Tregs - suppression
- HLA-DR$^+$ Tregs - suppression

#### 4. Total B cells

- PD-L1$^+$ B cells - activation/inhibition
- PD-1$^+$ B cells- activation/inhibition

#### 5. Total NK

- PD-L1$^+$ NK - inhibition
- PD-1$^+$ NK - activation/inhibition
- Tim-3$^+$ NK - activation/inhibition
- Ki67$^+$ NK - proliferation
- NKp30$^+$ NK - activation
- NKp46$^+$ NK - activation
- NKG2D$^+$ NK - activation
- CD226$^+$ NK - adhesion/activation
- Mature (CD16$^+$ CD56$^{dim}$) NK - lytic
  - PD-L1$^+$ mature NK - inhibition
  - PD-1$^+$ mature NK - activation/inhibition
  - Tim-3$^+$ mature NK - activation/inhibition
  - Ki67$^+$ mature NK - proliferation
  - NKp30$^+$ mature NK - activation
  - NKp46$^+$ mature NK - activation
  - NKG2D$^+$ mature NK - activation
  - CD226$^+$ mature NK - adhesion/activation
- Functional intermediate (CD16$^+$ CD56$^{br}$) NK - *lytic, cytokine production*
  - PD-L1$^+$ functional intermediate NK - inhibition
  - PD-1$^+$ functional intermediate NK - activation/inhibition
  - Tim-3$^+$ functional intermediate NK - activation/inhibition
  - Ki67$^+$ functional intermediate NK - proliferation
  - NKp30$^+$ functional intermediate NK - activation
  - NKp46$^+$ functional intermediate NK - activation
  - NKG2D$^+$ functional intermediate NK - activation
  - CD226$^+$ functional intermediate NK - adhesion/activation
- Immature (CD16$^-$ CD56$^{br}$) NK – cytokine production
  - PD-L1$^+$ immature NK - inhibition
  - PD-1$^+$ immature NK - activation/inhibition
  - Tim-3$^+$ immature NK - activation/inhibition
  - Ki67$^+$ immature NK - proliferation
  - NKp30$^+$ immature NK - activation
  - NKp46$^+$ immature NK - activation
  - NKG2D$^+$ immature NK - activation
  - CD226$^+$ immature NK - adhesion/activation
- Unconventional (CD16$^-$ CD56$^{dim}$) NK - non-lytic, non-cytokine production

#### 6. Total NK-T

- PD-L1$^+$ NK-T - inhibition
- PD-1$^+$ NK-T - activation/inhibition
- Tim-3$^+$ NK-T - inhibition
- Ki67$^+$ NK-T - proliferation

#### 7. Total cDC

- PD-L1$^+$ cDC - inhibition
- PD-1$^+$ cDC - activation/inhibition
- Tim-3$^+$ cDC - inhibition
- Ki67$^+$ cDC - proliferation

#### 8. Total pDC

- PD-L1$^+$ pDC - inhibition
- PD-1$^+$ pDC - activation/inhibition
- Tim-3$^+$ pDC - inhibition
- Ki67$^+$ pDC - proliferation

#### 9. Total MDSC

- PD-L1$^+$ MDSC - inhibition
- PD-1$^+$ MDSC - activation/inhibition
- CD16$^+$ MDSC - immature/suppression
- Monocytic (CD14$^+$ CD15$^-$) MDSC
  - PD-L1$^+$ mMDSC - inhibition
  - PD-1$^+$ mMDSC - activation/inhibition
  - CD16$^+$ mMDSC - immature/suppression
- Granulocytic (CD14$^-$ CD15$^+$) MDSC
  - PD-L1$^+$ gMDSC - inhibition
  - PD-1$^+$ gMDSC - activation/inhibition
  - CD16$^+$ gMDSC - immature/suppression

#### 10. Total Monocytes

- PD-L1$^+$ Monocytes - inhibition
- PD-1$^+$ Monocytes - activation/inhibition
- Classical Monocytes - phagocytic
  - PD-L1$^+$ Classical Monocytes - inhibition
  - PD-1$^+$ Classical Monocytes - activation/inhibition
- Intermediate Monocytes - *phagocytic/proinflammatory*
  - PD-L1$^+$ Intermediate Monocytes - inhibition
  - PD-1$^+$ Intermediate Monocytes - activation/inhibition
- Non-Classical Monocytes - *proinflammatory*
  - PD-L1$^+$ Non-Classical Monocytes - inhibition
  - PD-1$^+$ Non-Classical Monocytes - activation/inhibition

cDC, conventional dendritic cells; CM, central memory; CTLA-4, cytotoxic T lymphocyte-associated protein-4; EM, effector memory; EMRA, terminally differentiated effector memory; FoxP3, forkhead box P3; gMDSCs, granulocytic myeloid derived suppressor cells; ICOS, inducible T cell co-stimulator; mMDSCs, monocytic MDSCs; NK, natural killer; pDC, plasmacytoid DC; PD-1, programmed cell death-1; PD-L1, programmed cell death ligand-1; Tim-3, T cell immunoglobulin and mucin domain-3; Tregs, regulatory T cells.

**Supplementary Table 3: PBMC subsets included in development of a peripheral immunoscore (Immunoscore #1) that are indicative of enhanced immune function and changes with immunotherapy.**

| "Classic Cell Types" | | "Refined Cell Subsets" reflecting immune function included in Peripheral Immunoscore #1 | | |
|---|---|---|---|---|
| **Description** | **Expected effect on outcome** | **Description** | **Expected effect on outcome** | **Reference *** |
| % CD4 | Positive | % CD73+ CD4 | Negative | (2) |
| | | % ki67+ Effector Memory[(a)] CD4 | Positive | (3, 4) |
| % CD8 | Positive | % CD73+ CD8 | Negative | (2, 5) |
| | | % ki67+ Effector Memory[(a)] CD8 | Positive | (3, 4) |
| % NK[(b)] | Positive | % Immature (CD16- CD56br) NK[(b)] | Negative | (6, 7) |
| % Treg[(c)] | Negative | % CD49d- Treg[(c)] | Negative | (8, ) |
| % MDSC[(d)] | Negative | % CD16+ MDSC[(d)] | Negative | (10, 11) |
| Ratio CD4:Treg | Positive | Ratio ki67+ EM CD4: CD49d- Treg | Positive | |
| Ratio CD8:Treg | Positive | Ratio ki67+ EM CD8: CD49d- Treg | Positive | |
| Ratio CD4:MDSC | Positive | Ratio ki67+ EM CD4: CD16+ MDSC | Positive | |
| Ratio CD8:MDSC | Positive | Ratio ki67+ EM CD8: CD16+ MDSC | Positive | |

Frequencies of PBMC subsets represented as a percentage of total PBMCs. EM, effector memory; NK, natural killer cell; MDSC, myeloid derived suppressor cell; PBMC, peripheral blood mononuclear cell; Treg, regulatory T cell.

(a) CD45RA-, CCR7-, (b) CD3-, CD56+, (c) CD4+, CD25br, FoxP3+, CD127-,
(d) CD11b+, CD33+, HLA-DR-, CD14+ or CD15+

* See the page before the protocol for a list of references.

#### Supplementary Table 4. Changes in peripheral immune profile induced after 6 and 12 weeks of therapy.

##### Arm A: Nivolumab

**6 Weeks vs Baseline**

| Subset | | Direction of Change | Median Baseline (n=15) | Median Week 6 (n=15) | % with >25% Increase | % with >25% Decrease | p-value |
|---|---|---|---|---|---|---|---|
| Classic | CD4 | = | 26.61 | 24.38 | 27 | 20 | 0.8904 |
| | CD8 | = | 19.73 | 19.37 | 13 | 7 | 0.7197 |
| | Treg | = | 0.71 | 0.61 | 33 | 27 | >0.9999 |
| | NK | = | 4.56 | 3.17 | 27 | 53 | 0.3591 |
| | NKT | = | 2.50 | 1.37 | 20 | 53 | 0.0730 |
| | cDC | = | 0.43 | 0.48 | 40 | 40 | 0.6788 |
| | pDC | = | 0.14 | 0.09 | 60 | 33 | >0.9999 |
| | MDSC | = | 1.17 | 0.68 | 40 | 53 | 0.4212 |
| | Monocytes | = | 22.33 | 21.77 | 27 | 27 | 0.7197 |
| | B cells | = | 8.03 | 9.42 | 40 | 20 | 0.2769 |
| Refined | **NK DN** | ↓ | 0.78 | 0.52 | 13 | 60 | **0.0248** |

**12 Weeks vs Baseline**

| Subset | | Direction Of Change | Median Baseline (n=15) | Median Week 12 (n=15) | % with >25% Increase | % with >25% Decrease | p-value |
|---|---|---|---|---|---|---|---|
| Classic | CD4 | = | 26.61 | 25.64 | 20 | 40 | 0.4543 |
| | CD8 | = | 19.73 | 16.62 | 33 | 0 | 0.3303 |
| | Treg | = | 0.71 | 0.54 | 27 | 40 | 0.7615 |
| | NK | = | 4.56 | 5.98 | 47 | 20 | 0.0946 |
| | NKT | = | 2.50 | 2.14 | 27 | 33 | >0.9999 |
| | cDC | = | 0.43 | 0.45 | 27 | 27 | 0.4212 |
| | pDC | = | 0.14 | 0.09 | 47 | 27 | 0.6603 |
| | MDSC | = | 1.17 | 1.50 | 40 | 27 | 0.8040 |
| | Monocytes | = | 22.33 | 23.15 | 20 | 47 | 0.4212 |
| | B cells | = | 8.03 | 11.70 | 53 | 20 | 0.3028 |
| Refined | | | | | | | |

##### Arm B: Capecitabine

**6 Weeks vs Baseline**

| Subset | | Direction of Change | Median Baseline (n=14) | Median Week 6 (n=14) | % with >25% Increase | % with >25% Decrease | p-value |
|---|---|---|---|---|---|---|---|
| Classic | CD4 | = | 28.92 | 29.04 | 29 | 14 | 0.8552 |
| | CD8 | = | 12.94 | 14.83 | 29 | 14 | 0.3575 |
| | Treg | = | 0.95 | 0.91 | 29 | 43 | 0.5508 |
| | NK | = | 4.68 | 6.82 | 50 | 7 | 0.2958 |
| | NKT | = | 1.51 | 1.40 | 29 | 21 | 0.5066 |
| | cDC | = | 0.45 | 0.50 | 50 | 21 | 0.4631 |
| | pDC | = | 0.07 | 0.08 | 50 | 43 | 0.6374 |
| | MDSC | = | 1.77 | 1.57 | 43 | 43 | 0.8552 |
| | Monocytes | = | 26.15 | 24.43 | 21 | 36 | 0.2166 |
| | B cells | = | 11.47 | 10.44 | 36 | 21 | 0.3258 |
| Refined | **CD4 naive** | ↑ | 3.29 | 4.78 | 50 | 0 | **0.0107** |
| | **CD8 CD73$^+$** | ↑ | 6.06 | 6.81 | 57 | 0 | **0.0203** |
| | **NK CD226$^+$** | ↑ | 0.56 | 0.87 | 57 | 21 | **0.0382** |
| | **CD8 ki67$^+$** | ↓ | 0.61 | 0.32 | 7 | 57 | **0.0185** |
| | **Intermediate Monocytes PD-L1$^+$** | ↓ | 0.58 | 0.45 | 14 | 57 | **0.0494** |

**12 Weeks vs Baseline**

| Subset | | Direction of Change | Median Baseline (n=12) | Median Week 12 (n=12) | % with >25% Increase | % with >25% Decrease | p-value |
|---|---|---|---|---|---|---|---|
| Classic | CD4 | = | 28.92 | 26.94 | 17 | 8 | 0.7910 |
| | CD8 | = | 12.94 | 15.18 | 42 | 25 | 0.5693 |
| | Treg | = | 0.95 | 0.88 | 17 | 42 | 0.6221 |
| | NK | = | 4.03 | 6.35 | 50 | 25 | 0.1763 |
| | NKT | = | 1.23 | 1.22 | 25 | 33 | 0.9063 |
| | cDC | = | 0.41 | 0.29 | 17 | 42 | 0.4767 |
| | pDC | = | 0.06 | 0.07 | 42 | 25 | 0.8336 |
| | MDSC | = | 1.85 | 2.10 | 33 | 33 | 0.9097 |
| | Monocytes | = | 26.15 | 27.32 | 25 | 33 | 0.4238 |
| | B cells | = | 9.12 | 10.10 | 33 | 17 | 0.7334 |
| Refined | **CD4 naive** | ↑ | 3.29 | 4.15 | 58 | 0 | **0.0161** |
| | **CD8 CD73$^+$** | ↑ | 6.06 | 7.51 | 67 | 8 | **0.0122** |
| | **CD8 naive** | ↑ | 3.43 | 4.47 | 58 | 8 | **0.0093** |
| | **NK NKp46$^+$** | ↑ | 1.56 | 2.34 | 58 | 17 | **0.0093** |
| | **CD8 ki67$^+$** | ↓ | 0.66 | 0.42 | 8 | 58 | **0.0280** |

##### Arm C: Nivolumab + Capecitabine

**6 Weeks vs Baseline**

| Subset | | Direction of Change | Median Baseline (n=13) | Median Week 6 (n=13) | % with >25% Increase | % with >25% Decrease | p-value |
|---|---|---|---|---|---|---|---|
| Classic | CD4 | = | 26.08 | 31.04 | 23 | 15 | 0.8926 |
| | CD8 | = | 16.79 | 15.41 | 8 | 15 | 0.1272 |
| | Treg | = | 1.02 | 1.09 | 31 | 38 | 0.4548 |
| | NK | = | 5.79 | 7.22 | 38 | 8 | 0.2734 |
| | NKT | = | 2.76 | 2.06 | 0 | 46 | 0.0081* |
| | **cDC** | ↑ | 0.36 | 0.49 | 54 | 0 | **0.0206** |
| | pDC | = | 0.08 | 0.09 | 38 | 23 | 0.4213 |
| | MDSC | = | 1.56 | 2.37 | 54 | 15 | 0.3670 |
| | Monocytes | = | 25.60 | 23.69 | 38 | 8 | 0.3396 |
| | B cells | = | 8.56 | 7.57 | 15 | 31 | 0.3757 |
| Refined | **CD4 EM ki67$^+$** | ↑ | 0.56 | 0.82 | 62 | 8 | **0.0367** |
| | **NK DN** | ↑ | 0.58 | 0.75 | 62 | 0 | **0.0327** |

**12 Weeks vs Baseline**

| Subset | | Direction of Change | Median Baseline (n=12) | Median Week 12 (n=12) | % with >25% Increase | % with >25% Decrease | p-value |
|---|---|---|---|---|---|---|---|
| Classic | CD4 | = | 24.46 | 30.97 | 25 | 8 | 0.9097 |
| | CD8 | = | 16.95 | 15.19 | 8 | 17 | 0.2334 |
| | Treg | = | 1.17 | 0.82 | 25 | 42 | 0.4800 |
| | NK | = | 4.98 | 6.10 | 50 | 17 | 0.2661 |
| | NKT | = | 3.10 | 1.98 | 8 | 42 | 0.2300 |
| | cDC | = | 0.36 | 0.32 | 42 | 42 | 0.7334 |
| | pDC | = | 0.09 | 0.08 | 42 | 33 | 0.9645 |
| | MDSC | = | 1.72 | 2.34 | 58 | 33 | 0.6221 |
| | Monocytes | = | 26.02 | 25.55 | 42 | 25 | 0.3804 |
| | B cells | = | 10.77 | 10.90 | 33 | 25 | >0.9999 |
| Refined | **NK DN** | ↑ | 0.58 | 0.71 | 50 | 0 | 0.0161 |

Changes in immune cell subsets after 6 and 12 weeks of therapy were calculated in each arm with a 2 tailed Wilcoxon signed-rank text. Subsets with significant changes at post timepoints vs. landmark include those with $p < 0.05$ (calculated by a two tailed Wilcoxon signed-rank test), difference in medians > 0.05, and ≥ 50% of patients having a > 25% change. Subsets meeting this criteria are indicated in bold text. The number of patients evaluated at each time point and in each arm are indicated. No adjustments for multiple comparisons were made. *Although significant p value, majority of patients do not change by >25%. cDC, conventional dendritic cells; DN, double negative; EM, effector memory; NK, natural killer cell; NKT, natural killer T cell; MDSC, myeloid derived suppressor cell; pDC, plasmacytoid dendritic cell; PD-L1, programmed cell death ligand-1; Treg, regulatory T cell. Source data are provided as a source data file.

#### Supplementary Table 5: Changes in peripheral immune profile across treatment arms

### **a** Changes after 6 weeks

| Subset (% of PBMC) | Arm A (Nivolumab) | | | Arm B (Capecitabine) | | | Arm C (Nivolumab + Capecitabine) | | | Arms A + C (Nivolumab +/- Capecitabine) | | |
|---|---|---|---|---|---|---|---|---|---|---|---|---|
| | Median Baseline (n=15) | Median Week 6 (n=15) | p | Median Baseline (n=14) | Median Week 6 (n=14) | p | Median Baseline (n=13) | Median Week 6 (n=13) | p | Median Baseline (n=28) | Median Week 6 (n=28) | p |
| CD4 | 26.61 | 24.38 | 0.890 | 28.92 | 29.04 | 0.855 | 26.08 | 31.04 | 0.893 | 26.57 | 26.22 | 0.762 |
| CD8 | 19.73 | 19.37 | 0.720 | 12.94 | 14.83 | 0.358 | 16.79 | 15.41 | 0.127 | 17.83 | 15.67 | 0.695 |
| Treg | 0.71 | 0.61 | >0.999 | 0.95 | 0.91 | 0.551 | 1.02 | 1.09 | 0.455 | 0.77 | 0.69 | 0.493 |
| NK | 4.56 | 3.17 | 0.359 | 4.68 | 6.82 | 0.296 | 5.79 | 7.22 | 0.273 | 4.57 | 5.00 | 0.955 |
| **NK-T** | 2.5 | 1.37 | 0.073 | 1.51 | 1.40 | 0.507 | 2.76 | 2.06 | 0.008* | 2.66 | 1.69 | **0.001** |
| **cDC** | 0.43 | 0.48 | 0.679 | 0.45 | 0.50 | 0.463 | 0.36 | 0.49 | **0.021** | 0.43 | 0.48 | 0.327 |
| pDC | 0.14 | 0.09 | >0.999 | 0.07 | 0.08 | 0.637 | 0.08 | 0.09 | 0.421 | 0.10 | 0.09 | 0.662 |
| Monocytes | 1.17 | 0.68 | 0.421 | 1.77 | 1.57 | 0.855 | 1.56 | 2.37 | 0.367 | 1.51 | 1.26 | 0.902 |
| MDSC | 22.33 | 21.77 | 0.720 | 26.15 | 24.43 | 0.217 | 25.6 | 23.69 | 0.340 | 24.52 | 23.63 | 0.728 |
| B cells | 8.03 | 9.42 | 0.277 | 11.47 | 10.44 | 0.326 | 8.56 | 7.57 | 0.376 | 8.30 | 9.16 | 0.779 |
| **NK DN** | 0.78 | 0.52 | **0.025** | 0.59 | 0.89 | 0.715 | 0.58 | 0.75 | **0.033** | 0.66 | 0.70 | 0.614 |
| **CD8 Ki67+** | 0.41 | 0.42 | 0.330 | 0.61 | 0.32 | **0.018** | 0.48 | 0.42 | 0.972 | 0.45 | 0.42 | 0.425 |
| **CD4 naïve** | 4.02 | 4.46 | 0.303 | 3.29 | 4.78 | **0.011** | 5.11 | 5.82 | 0.216 | 4.43 | 5.00 | 0.109 |
| **CD8 CD73+** | 8.76 | 6.94 | 0.890 | 6.06 | 6.81 | **0.020** | 7.95 | 6.53 | 0.273 | 7.99 | 6.56 | 0.508 |
| **NK CD226+** | 0.55 | 0.40 | 0.798 | 0.56 | 0.87 | **0.038** | 0.66 | 0.7 | 0.916 | 0.62 | 0.60 | 0.902 |
| **CD4 EM ki67+** | 0.55 | 0.56 | 0.303 | 0.66 | 0.51 | 0.079 | 0.56 | 0.82 | **0.037** | 0.55 | 0.66 | **0.018** |

### **b** Changes after 12 weeks

| Subset (% of PBMC) | Arm A (Nivolumab) | | | Arm B (Capecitabine) | | | Arm C (Nivolumab + Capecitabine) | | | Arms A + C (Nivolumab +/- Capecitabine) | | |
|---|---|---|---|---|---|---|---|---|---|---|---|---|
| | Median Baseline (n=15) | Median Week 12 (n=15) | p | Median Baseline (n=12) | Median Week 12 (n=12) | p | Median Baseline (n=12) | Median Week 12 (n=12) | p | Median Baseline (n=27) | Median Week 12 (n=27) | p |
| CD4 | 26.61 | 25.64 | 0.454 | 28.92 | 26.94 | 0.791 | 24.46 | 30.97 | 0.910 | 26.53 | 27.05 | 0.414 |
| CD8 | 19.73 | 16.62 | 0.330 | 12.94 | 15.18 | 0.569 | 16.95 | 15.19 | 0.233 | 17.87 | 16.54 | 0.804 |
| Treg | 0.71 | 0.54 | 0.762 | 0.95 | 0.88 | 0.622 | 1.17 | 0.82 | 0.480 | 0.73 | 0.64 | 0.374 |
| NK | 4.56 | 5.98 | 0.095 | 4.03 | 6.35 | 0.176 | 4.98 | 6.10 | 0.266 | 4.56 | 5.98 | 0.039* |
| NK-T | 2.5 | 2.14 | >0.999 | 1.23 | 1.22 | 0.906 | 3.10 | 1.98 | 0.230 | 2.7 | 2.14 | 0.354 |
| cDC | 0.43 | 0.45 | 0.421 | 0.41 | 0.29 | 0.477 | 0.36 | 0.32 | 0.733 | 0.43 | 0.39 | 0.374 |
| pDC | 0.14 | 0.09 | 0.660 | 0.06 | 0.07 | 0.834 | 0.09 | 0.08 | 0.965 | 0.11 | 0.09 | 0.657 |
| Monocytes | 1.17 | 1.5 | 0.804 | 1.85 | 2.10 | 0.910 | 1.72 | 2.34 | 0.622 | 1.56 | 1.89 | 0.732 |
| MDSC | 22.33 | 23.15 | 0.421 | 26.15 | 27.32 | 0.424 | 26.02 | 25.55 | 0.380 | 24.84 | 23.29 | 0.897 |
| B cells | 8.03 | 11.7 | 0.303 | 9.12 | 10.10 | 0.733 | 10.77 | 10.90 | >0.999 | 8.56 | 11.68 | 0.290 |
| **CD4 naïve** | 4.02 | 4.65 | 0.041* | 3.29 | 4.15 | **0.016** | 5.02 | 5.18 | 0.209 | 4.30 | 4.90 | 0.013* |
| **CD8 CD73+** | 8.76 | 6.57 | 0.804 | 6.06 | 7.51 | **0.012** | 7.83 | 7.25 | 0.910 | 7.95 | 6.57 | 0.915 |
| **CD8 naïve** | 4.76 | 4.53 | 0.851 | 3.43 | 4.47 | **0.009** | 5.03 | 5.19 | 0.380 | 4.93 | 4.53 | 0.568 |
| **NK NKp46+** | 1.26 | 1.55 | 0.489 | 1.56 | 2.34 | **0.009** | 2.60 | 2.33 | 0.970 | 1.76 | 1.98 | 0.540 |
| **CD8 ki67+** | 0.41 | 0.45 | 0.691 | 0.66 | 0.42 | **0.028** | 0.50 | 0.40 | 0.850 | 0.48 | 0.45 | 0.665 |
| **NK DN** | 0.78 | 0.41 | 0.616 | 0.55 | 0.92 | 0.875 | 0.58 | 0.71 | **0.016** | 0.65 | 0.65 | 0.310 |

Changes in immune cell subsets after 6 and 12 weeks of therapy were calculated in each arm with a 2 tailed Wilcoxon signed-rank text. The number of patients evaluated at each time point and in each arm are indicated. Subsets with significant changes at post timepoints vs. landmark include those with $p < 0.05$ (calculated by a two tailed Wilcoxon signed-rank test), difference in medians > 0.05, and $\geq$ 50% of patients having a > 25% change. Subsets meeting this criteria are indicated with bold text. No adjustments for multiple comparisons were made. *Although significant p value, majority of patients do not change by >25%. cDC, conventional dendritic cells; DN, double negative; EM, effector memory; NK, natural killer cell; NKT, natural killer T cell; MDSC, myeloid derived suppressor cell; pDC, plasmacytoid dendritic cell; PD-L1, programmed cell death ligand-1; Treg, regulatory T cell. Source data are provided as a source data file.

**Supplementary Table 6. Invasive disease-free survival (iDFS) stratified by treatment arm and baseline ctDNA.**

| Risk Variable | Level | Number of Patients | Median iDFS (months) | 95% CI | | Log-rank p-value | iDFS probability at 1 year (SE) | iDFS probability at 2 year (SE) | iDFS probability at 3 year (SE) |
|---|---|---|---|---|---|---|---|---|---|
| All patients | | | - | 20.66 | - | - | 0.73 (0.07) | 0.63 (0.08) | 0.54 (0.11) |
| By treatment arm (n = 45) | A Nivolumab | 15 | 14.85 | 3.61 | - | 0.08 | 0.53 (0.13) | 0.47 (0.13) | 0.47 (0.13) |
| | B Capecitabine | 15 | - | 4.52 | - | | 0.67 (0.12) | 0.53 (0.15) | 0.53 (0.15) |
| | C Combo | 15 | 32.82 | 32.82 | - | | 1 (0) | 0.91 (0.09) | - |
| Baseline ctDNA (n = 38) | No | 25 | - | 32.82 | - | <.0001 | 1 (0) | 0.82 (0.10) | 0.62 (0.19) |
| | Yes | 13 | 4.52 | 3.21 | 8.98 | | 0.21 (0.12) | 0.205 (0.12) | - |

**Abbreviations: iDFS, invasive disease-free survival; CI, confidence interval; ctDNA, circulating tumor DNA; -, Not reached yet; <0.0001, p-value was from the log-rank test; SE, standard error.**

**Supplementary Table 7: Landmark levels of immune subsets that associate with disease recurrence across treatment arms**

### Landmark Levels of subsets that associate with recurrence

| **Subset** (% of PBMC) | **Arm A (Nivolumab)** | | | **Arm B (Capecitabine)** | | | **Arm C (Nivolumab + Capecitabine)** | | | **Arms A + C (Nivolumab +/- Capecitabine)** | | |
|---|---|---|---|---|---|---|---|---|---|---|---|---|
| | Median R (n=8) | Median NR (n=7) | p | Median R (n=5) | Median NR (n=9) | p | Median R (n=2) | Median NR (n=11) | p | Median R (n=10) | Median NR (n=18) | p |
| CD4 | 27.05 | 26.35 | 0.397 | 27.52 | 30.32 | 1.000 | 32.20 | 22.83 | 0.641 | 27.81 | 24.46 | 0.382 |
| CD8 | 20.59 | 17.40 | 0.281 | 12.64 | 13.11 | 0.797 | 10.82 | 17.78 | 0.231 | 19.98 | 17.59 | 0.464 |
| **Treg** | 0.71 | 0.66 | 0.281 | 0.91 | 1.04 | 0.438 | 2.16 | 0.90 | **0.026** | 0.81 | 0.77 | 0.494 |
| NK | 3.18 | 4.59 | 0.463 | 6.11 | 4.37 | 0.699 | 6.25 | 5.79 | 1.000 | 3.22 | 5 | 0.286 |
| **NK-T** | 1.82 | 3.22 | 0.189 | 1.20 | 2.65 | 0.240 | 1.14 | 3.44 | 0.103 | 1.71 | 3.33 | **0.031** |
| cDC | 0.43 | 0.43 | 0.613 | 0.39 | 0.47 | 0.898 | 0.47 | 0.36 | 0.923 | 0.43 | 0.43 | 0.981 |
| pDC | 0.15 | 0.11 | 0.779 | 0.05 | 0.07 | 0.606 | 0.08 | 0.08 | 0.923 | 0.14 | 0.09 | 0.494 |
| Monocytes | 1.55 | 0.49 | 0.779 | 1.65 | 1.79 | 0.797 | 1.83 | 1.56 | 0.923 | 1.55 | 1.51 | 0.944 |
| MDSC | 21.83 | 24.84 | 0.397 | 30.15 | 21.98 | 0.438 | 25.38 | 25.60 | 0.923 | 23.27 | 25.22 | 0.796 |
| B cells | 9.20 | 7.10 | 0.779 | 7.67 | 12.37 | 0.797 | 14.45 | 5.28 | 0.308 | 9.46 | 7.01 | 0.382 |
| **NK-T PD-1+** | 0.32 | 0.88 | **0.021** | 0.40 | 0.43 | 0.606 | 0.46 | 1.25 | 0.154 | 0.37 | 1.11 | **0.001** |
| **CD8 naïve** | 5.55 | 2.62 | **0.021** | 3.37 | 3.67 | 0.797 | 5.50 | 5.13 | 1.000 | 5.55 | 4.33 | 0.099 |
| **Treg HLA-DR+** | 0.39 | 0.23 | **0.043** | 0.36 | 0.40 | 1.000 | 0.77 | 0.35 | **0.026** | 0.47 | 0.30 | 0.051 |
| **NK NKp30+** | 1.71 | 2.97 | **0.021** | 4.46 | 2.99 | 0.797 | 4.84 | 2.35 | 1.000 | 1.71 | 2.73 | 0.064 |
| **Int. Monocytes** | 1.07 | 1.22 | 1.000 | 1.89 | 1.00 | **0.042** | 2.04 | 1.41 | 0.513 | 1.09 | 1.31 | 0.524 |
| **Non class monocyte** | 1.71 | 1.65 | 0.779 | 2.63 | 0.93 | **0.042** | 2.48 | 1.91 | 0.923 | 1.70 | 1.74 | 0.869 |
| **Treg ICOS+** | 0.57 | 0.40 | 0.105 | 0.57 | 0.55 | 0.518 | 1.32 | 0.58 | **0.026** | 0.61 | 0.50 | 0.191 |
| **Treg CD49d-** | 0.54 | 0.48 | 0.271 | 0.66 | 0.77 | 0.518 | 1.80 | 0.74 | **0.026** | 0.60 | 0.57 | 0.464 |

The peripheral immune profile at landmark was compared between patients who developed a recurrence (R) and those that did not (NR). The number of patients compared with R or NR in each arm are indicated. Notable subsets with significant differences include those with $p < 0.05$ (calculated by a two tailed Mann-Whitney test), and difference in medians > 0.05 of PBMCs. Subsets meeting this criteria are indicated with bold text. No adjustments were performed for multiple comparisons. Source data are provided as a source data file.

#### Supplementary Table 8: Changes in immune subsets that associate with disease recurrence across treatment arms

**a** **% change at 6 weeks vs baseline in levels of immune subsets that associate with recurrence**

| **Subset** (% change at 6 weeks vs Baseline) | **Arm A (Nivolumab)** | | | **Arm B (Capecitabine)** | | | **Arm C (Nivolumab + Capecitabine)** | | |
|---|---|---|---|---|---|---|---|---|---|
| | Median R (n=8) | Median NR (n=7) | p | Median R (n=5) | Median NR (n=9) | p | Median R (n=2) | Median NR (n=11) | p |
| CD4 | -3.19 | -7.62 | 0.867 | -7.13 | -0.17 | 1.000 | 11.38 | -14.10 | 0.641 |
| CD8 | 6.33 | 11.38 | 0.867 | 17.59 | 4.88 | 0.147 | 32.53 | -13.00 | 0.051 |
| Treg | 12.56 | 1.17 | 0.867 | -4.15 | -15.45 | 1.000 | -38.82 | -6.58 | 0.154 |
| NK | -28.37 | -10.59 | 0.463 | -2.09 | 38.36 | 0.898 | 34.92 | 12.00 | 0.923 |
| NK-T | -48.64 | -13.85 | 0.121 | -5.16 | 4.22 | 0.699 | -42.03 | -6.07 | 0.410 |
| cDC | 31.05 | -15.37 | 0.336 | 33.00 | 22.96 | 0.898 | 91.45 | 8.19 | 0.231 |
| pDC | 58.17 | 26.30 | 0.779 | -49.53 | 39.36 | 0.518 | 115.25 | 15.81 | 0.308 |
| Monocytes | -53.74 | 25.27 | 0.152 | 65.02 | -7.39 | 0.898 | -6.78 | 36.46 | 0.513 |
| MDSC | 7.82 | -5.05 | 0.281 | -20.38 | -7.69 | 0.240 | 2.83 | 3.67 | 0.513 |
| B cells | 10.38 | 25.19 | 0.336 | 30.42 | -4.94 | 0.364 | -23.74 | -9.36 | 0.769 |
| **NK DP** | -44.19 | 0.00 | **0.029** | 12.44 | 7.76 | 0.438 | 54.34 | -19.35 | 0.308 |
| **CD8 EMRA** | 19.10 | 1.11 | 0.536 | 35.13 | 21.45 | 1.000 | 40.96 | -2.92 | **0.026** |

**b** **% change at 12 weeks vs baseline in levels of immune subsets that associate with recurrence**

| **Subset** (% change at 12 weeks vs Baseline) | **Arm A (Nivolumab)** | | | **Arm B (Capecitabine)** | | | **Arm C (Nivolumab + Capecitabine)** | | |
|---|---|---|---|---|---|---|---|---|---|
| | Median R (n=8) | Median NR (n=7) | p | Median R (n=5) | Median NR (n=8) | p | Median R (n=2) | Median NR (n=10) | p |
| CD4 | 6.80 | -25.82 | 0.613 | -4.33 | 1.52 | 0.755 | 4.88 | -12.73 | 0.758 |
| CD8 | 9.15 | 6.78 | 0.955 | 14.89 | 2.89 | 0.876 | 36.41 | -7.83 | **0.030** |
| Treg | 1.75 | -13.47 | 0.867 | 21.12 | -10.40 | 0.343 | -12.41 | -5.40 | 1.000 |
| NK | 27.67 | 19.86 | 0.867 | 32.65 | 17.47 | 1.000 | 66.62 | 18.62 | 0.485 |
| NK-T | 8.10 | -15.53 | 0.336 | 8.43 | -0.57 | 0.876 | 26.72 | -20.97 | 0.273 |
| **cDC** | -7.75 | -9.79 | 0.955 | 22.14 | -32.73 | **0.030** | 15.11 | -4.83 | 1.000 |
| pDC | 47.66 | -11.05 | 0.779 | 49.86 | -5.03 | 0.149 | 213.66 | -2.97 | 0.606 |
| **Monocytes** | -19.81 | 173.35 | 0.232 | 93.28 | -29.68 | **0.030** | 75.98 | 86.29 | 1.000 |
| MDSC | -19.05 | 2.77 | 0.779 | -13.73 | -26.46 | 0.343 | -14.71 | 27.14 | 0.273 |
| B cells | 19.15 | 91.31 | 0.189 | -13.00 | -6.08 | 0.530 | -12.99 | -2.88 | 0.758 |
| **CD4 naïve** | 7.34 | 74.69 | **0.021** | 8.34 | 36.13 | 0.755 | 6.16 | 7.58 | 1.000 |
| **CD8 EMRA** | 42.61 | 6.23 | 0.463 | 18.71 | -13.19 | 0.268 | 69.48 | -16.52 | **0.030** |

Changes in the peripheral immune profile at 6 weeks vs baseline (**A**) and 12 weeks vs baseline (**B**) were compared between patients who developed disease recurrence (R) and those that did not develop disease recurrence (NR). The number of patients compared with R or NR in each arm are indicated. Notable subsets with significant differences include those with $p < 0.05$ (calculated by a two tailed Mann-Whitney test), and difference in medians > 0.05 of PBMCs. Subsets meeting this criteria are indicated with bold text. No adjustments were performed for multiple comparisons. Source data are provided as a source data file.

**Supplementary Table 9: PBMC subsets included in development of a peripheral immunoscore (Immunoscore #2) at landmark that associates with disease recurrence in patients receiving immunotherapy**

| **"Classic Cell Types"** | | **"Refined Cell Subsets" reflecting immune function included in Peripheral Immunoscore #2** | | |
|---|---|---|---|---|
| **Description** | **Expected effect on outcome** | **Description** | **Expected effect on outcome** | **References*** |
| % CD4 | Positive | % PD-1+ CD4 | Negative | (12) |
| | | % Effector Memory[a] CD4 | Positive | (13, 14) |
| % CD8 | Positive | % PD-1+ CD8 | Negative | (15, 16) |
| | | % Effector Memory[a] CD8 | Positive | (13,14) |
| % NK[b] | Positive | % NKp30+ NK[b] | Positive | (17) |
| % Treg[c] | Negative | % ICOS+ Treg[c] | Negative | (18) |
| % MDSC[d] | Negative | % PD-L1+ MDSC[d] | Negative | (19, 20) |
| Ratio CD4:Treg | Positive | Ratio EM CD4: ICOS+ Treg | Positive | |
| Ratio CD8:Treg | Positive | Ratio EM CD8: ICOS+ Treg | Positive | |
| Ratio CD4:MDSC | Positive | Ratio EM CD4: PD-L1+ MDSC | Positive | |
| Ratio CD8:MDSC | Positive | Ratio EM CD8: PD-L1+ MDSC | Positive | |

Frequencies of PBMC subsets represented as a percentage of total PBMCs.
EM, effector memory; NK, natural killer cell; MDSC, myeloid derived suppressor cell; PBMC, peripheral blood mononuclear cell; Treg, regulatory T cell.

(a) CD45RA-, CCR7-, (b) CD3-, CD56+, (c) CD4+, CD25br, FoxP3+, CD127-,
(d) CD11b+, CD33+, LA-DR-, CD14+ or CD15+

* See the page before the protocol for a list of references.

**Supplementary Table 10: ctDNA dynamics by treatment arm in patients who had ctDNA collected.**

| Patient Number | Total | Arm A | Arm B | Arm C |
|---|---|---|---|---|
| **Landmark ctDNA (n=38)** | | | | |
| Yes | 13 (34%) | 6 (46%) | 4 (33%) | 3 (23%) |
| No | 25 (66%) | 7 (54%) | 8 (67%) | 10 (77%) |
| **6-week ctNDA (n=37)** | | | | |
| Yes | 9 (24%) | 6 (46%) | 2 (18%) | 1 (8%) |
| No | 28 (76%) | 7 (54%) | 9 (82%) | 12 (92%) |
| **12-week ctDNA (n=32)** | | | | |
| Yes | 7 (22%) | 4 (36%) | 3 (30%) | 0 (0%) |
| No | 25 (78%) | 7 (64%) | 7 (70%) | 11 (100%) |

**Supplementary Table 11: ctDNA dynamics in patients with ctDNA detected at any timepoint.**

| Patient Number | Baseline ctDNA | 6-week ctDNA | 12-week ctDNA | ctDNA at Recurrence | Distance Recurrence |
|---|---|---|---|---|---|
| 13 | Yes | No | No | - | No |
| 14 | Yes | No | Yes | Yes | Yes |
| 15 | Yes | Yes | - | Yes | Yes |
| 19 | Yes | Yes | Yes | - | Yes |
| 20 | Yes | Yes | Yes | Yes | Yes |
| 25 | Yes | Yes | Yes | Yes | Yes |
| 27 | No | Yes | No | - | Yes |
| 31 | Yes | Yes | - | Yes | Yes |
| 33 | Yes | No | - | - | No |
| 35 | Yes | Yes | Yes | - | Yes |
| 38 | Yes | - | - | - | Yes |
| 39 | Yes | Yes | Yes | - | Yes |
| 40 | Yes | Yes | Yes | - | Yes |
| 45 | Yes | No | No | - | No |

ctDNA, circulating tumor DNA

### Supplementary Table 12: Antibodies used for flow cytometry

| | Marker | Fluorophore | Company | Catalog # | Clone | Dilution |
|---|---|---|---|---|---|---|
| **Panel 1** | | | | | | |
| | **Marker** | **Fluorophore** | **Company** | **Catalog #** | **Clone** | **Dilution** |
| 1 | CTLA4 | FITC | LS Bio | ls-c62860 | A3.4H2.H12 | 1:20 |
| 2 | PD-1 | PE | Biolegend | 329906 | EH12.2H7 | 1:100 |
| 3 | 4-1BB | PerCP-Cy5.5 | Biolegend | 309814 | 4B4-1 | 1:10 |
| 4 | PD-L1 | PE-Cy7 | BD Bioscience | 558017 | MIH1 | 1:100 |
| 5 | Tim3 | BV421 | Biolegend | 345008 | F38-2E2 | 1:50 |
| 6 | CCR7 | BV510 | Biolegend | 353232 | G043H7 | 1:20 |
| 7 | CD4 | BV605 | Biolegend | 317438 | OKT4 | 1:100 |
| 8 | **<u>Ki67**</u>** | BV711 | Biolegend | 350516 | Ki-67 | 1:100 |
| 9 | CD8 | BV785 | Biolegend | 301046 | RPA-T8 | 1:200 |
| 10 | CD73 | APC | Biolegend | 344006 | AD2 | 1:100 |
| 11 | live/dead | dapi | Invitrogen | L23105 | N/A | 1:1000 |
| 12 | CD45RA | AF700 | Biolegend | 304120 | HI100 | 1:200 |
| **Panel 2** | | | | | | |
| | **Marker** | **Fluorophore** | **Company** | **Catalog #** | **Clone** | **Dilution** |
| 1 | CTLA4 | FITC | LS Bio | ls-c62860 | A3.4H2.H12 | 1:20 |
| 2 | PD-1 | PE | Biolegend | 329906 | EH12.2H7 | 1:100 |
| 3 | ICOS | PerCP-Cy5.5 | Biolegend | 313518 | C398.4A | 1:50 |
| 4 | PD-L1 | PE-Cy7 | BD Bioscience | 558017 | MIH1 | 1:100 |
| 5 | **<u>FoxP3**</u>** | Pacific Blue | Biolegend | 320116 | 206D | 1:20 |
| 6 | CD49d | BV510 | Biolegend | 304318 | 9F10 | 1:50 |
| 7 | CD4 | BV605 | Biolegend | 317438 | OKT4 | 1:100 |
| 8 | **<u>Ki67**</u>** | BV711 | Biolegend | 350516 | ki-67 | 1:100 |
| 9 | CD38 | BV785 | Biolegend | 303530 | HIT2 | 1:50 |
| 10 | HLA-DR | BUV395 | BD Bioscience | 564040 | G46-6 | 1:200 |
| 11 | live/dead | dapi | Invitrogen | L23105 | N/A | 1:1000 |
| 12 | CD25 | APC | Biolegend | 356110 | M-A251 | 1:100 |
| 13 | CD45RA | AF700 | Biolegend | 304120 | HI100 | 1:200 |
| 14 | CD127 | APC-eF780 | Ebioscience | 47-1278-42 | eBioRDR5 | 1:50 |
| **Panel 3** | | | | | | |
| | **Marker** | **Fluorophore** | **Company** | **Catalog #** | **Clone** | **Dilution** |
| 1 | CD3 | FITC | Biolegend | 300306 | HIT3a | 1:200 |
| 2 | PD-1 | PE | Biolegend | 329906 | EH12.2H7 | 1:100 |
| 3 | CD303 | PerCP-Cy5.5 | Biolegend | 354210 | 201a | 1:100 |
| 4 | PD-L1 | PE-Cy7 | BD Bioscience | 558017 | MIH1 | 1:100 |
| 5 | Tim3 | BV421 | Biolegend | 345008 | F38-2E2 | 1:50 |
| 6 | NKp30 | BV510 | BD Bioscience | 743170 | p30-15 | 1:50 |
| 7 | CD56 | BV605 | Biolegend | 318334 | HCD56 | 1:50 |
| 8 | NKp46 | BV650 | Biolegend | 331927 | 9.00E+02 | 1:50 |
| 9 | **<u>Ki67**</u>** | BV711 | Biolegend | 350516 | Ki-67 | 1:100 |
| 10 | NKG2D | BV785 | BD Bioscience | 743560 | 1D11 | 1:20 |
| 11 | CD226 | BUV395 | BD Bioscience | 742498 | DX11 | 1:100 |
| 12 | live/dead | dapi | Invitrogen | L23105 | N/A | 1:1000 |
| 13 | HLA-DR | APC | Biolegend | 307610 | L243 | 1:100 |
| 14 | CD16 | AF700 | Biolegend | 302026 | 3G8 | 1:200 |
| 15 | CD1c | APC-Cy7 | Biolegend | 331520 | L161 | 1:200 |
| **Panel 4** | | | | | | |
| | **Marker** | **Fluorophore** | **Company** | **Catalog #** | **Clone** | **Dilution** |
| 1 | CD15 | FITC | Biolegend | 301904 | HI98 | 1:10 |
| 2 | PD-1 | PE | Biolegend | 329906 | EH12.2H7 | 1:100 |
| 3 | CD19 | PerCP-Cy5.5 | Biolegend | 302230 | HIB19 | 1:100 |
| 4 | PD-L1 | PE-Cy7 | BD Bioscience | 558017 | MIH1 | 1:100 |
| 5 | CD14 | BV421 | Biolegend | 301830 | M5E2 | 1:50 |
| 6 | CD16 | BV510 | Biolegend | 302048 | 3G8 | 1:200 |
| 7 | HLA-DR | BV605 | Biolegend | 307640 | L243 | 1:100 |
| 8 | CD11b | BV785 | Biolegend | 101243 | M1/70 | 1:200 |
| 9 | live/dead | dapi | Invitrogen | L23105 | N/A | 1:1000 |
| 10 | CD33 | APC | Biolegend | 303408 | WM53 | 1:100 |

**<u>Bold Underlined - Intracellular staining</u>**

**OXEL: A pilot study of immune checkpoint or capecitabine or combination therapy as adjuvant therapy for triple negative breast cancer with residual disease following neoadjuvant chemotherapy**

**Georgetown Protocol #:** 2017-1535 (CRC # 112017-01) BMS CA209-8CL

**Clinicaltrials.gov #:** NCT03487666
**Protocol Version Date:** Version 6.1,6/25/2020

**Principal Investigator (PI):** Candace Mainor, MD
Lombardi Comprehensive Cancer Center
Georgetown University Medical Center
3800 Reservoir Road, NW
Washington, DC 20057


**Correlative Science PI:** Louis Weiner, MD, PhD
Lombardi Comprehensive Cancer Center
Georgetown University Medical Center
3800 Reservoir Road, NW
Washington, DC 20057

Jeffrey Schlom, PhD
Center for Cancer Research
National Cancer Institute (NCI)
Building 10, Room 8B09
Bethesda, MD 20892-1750


**Study Chair:** Paula Pohlmann, MD, MSc, PhD

Lombardi Comprehensive Cancer Center
Georgetown University Medical Center

**Co-Investigators:** Claudine Isaacs, MD
Lombardi Comprehensive Cancer Center
Georgetown University Medical Center

Sandra Swain, MD
Georgetown University

**Corporate Collaborators:** Bristol-Meyers Squibb

**Biostatistician**: Hongkun Wang, PhD
Lombardi Comprehensive Cancer Center
Georgetown University Medical Center

**Study Drug:** Nivolumab

**IND Number:** 137808

**Clinical Phase:** Phase II

**Number of Subjects:** 45

The study will be conducted according to the protocol and in compliance with Good Clinical Practice (GCP), with the Declaration of Helsinki, and with other applicable regulatory requirements including but not limited to Institutional Review Board/Ethics Committee (IRB/EC) approval.

**Confidentiality Statement**

All information contained in this document is privileged and confidential. Any distribution, copying, or disclosure is strictly prohibited without prior written approval by the Sponsor.

**SPONSOR SIGNATURE PAGE**

**Declaration of Sponsor or Responsible Medical Officer**

Title: OXEL study - A pilot study of immune checkpoint or capecitabine or combination therapy as adjuvant therapy for triple negative breast cancer with residual disease following neoadjuvant chemotherapy

This study protocol was subjected to critical review and has been approved by the Sponsor. The information it contains is consistent with the current risk/benefit evaluation of the investigational product as well as with the moral, ethical, and scientific principles governing clinical research as set out in the Declaration of Helsinki and the guidelines on Good Clinical Practice.

________________________________________ _______________________

Candace Mainor, MD                Date
Lombardi Comprehensive Cancer Center
Georgetown University Medical Center

#### INVESTIGATOR SIGNATURE PAGE

##### Declaration of the Investigator

Title: OXEL study - A pilot study of immune checkpoint or capecitabine or combination therapy as adjuvant therapy for triple negative breast cancer with residual disease following neoadjuvant chemotherapy

I have read this study protocol, including all appendices. By signing this protocol, I agree to conduct the clinical study, following approval by an Independent Ethics Committee (IEC)/Institutional Review Board (IRB), in accordance with the study protocol, the current International Conference on Harmonisation (ICH) Guideline for Good Clinical Practice (GCP), and applicable regulatory requirements. I will ensure that all personnel involved in the study under my direction will be informed about the contents of this study protocol and will receive all necessary instructions for performing the study according to the study protocol.

________________________________________ _______________________

Candace Mainor, MD Date
Lombardi Comprehensive Cancer Center
Georgetown University Medical Center

#### Document History

| Document | Version Date | Summary of changes and rationale |
|---|---|---|
| Version 3 | 07MAY2018 | Administrative changes and the removal of EKG tests.<br><br>**Protocol Updates:**<br>• Updated version date in footer.<br>• Page 1, Protocol version date: changed to v3, date 07MAY18<br>• Page 7, study synopsis: updated protocol number and NCT number<br>• Page 21, birth control: changed men must not donate sperm to seven months to be consistent with consent<br>• Page 28, 6.1 Table of events: removed 12 lead EKG<br><br>**ICF Updates:**<br>• Updated version date in footer.<br>• Page 3, "End of study": removed EKG<br>• Page 17: added Dr. Christopher Gallagher |
| Version 4 | 11JUL18 | **Protocol Updates:**<br>• Page 2: Added IND number<br>• Page 7, 16, 41: Made the distinction between secondary objectives and exploratory objectives. Note - no new objectives were added.<br>• Page 18 to 19: added additional information to clarify inclusion criteria after feedback received during the SIV:<br>-Added RCB II or III<br>-Added need to have tissue in the lead institution by C1D1<br>-Added timeline for surgery and radiation completeness<br>-Added info on radiation received<br>-Limited PS to 0-1<br>-Added need for normal thyroid function |

| | | |
|---|---|---|
| | | • Page 19 to 20: added additional information to clarify exclusion criteria after feedback received during the SIV:<br>-Added clarification on autoimmune diseases<br>-Added clarification on definition of active hepatitis B and C<br>• Page 20: included endocrine therapy to anti-cancer therapies excluded during study<br>• Page 26: added clarification on the dose of nivolumab (was already in the schema but was added to the text)<br>• Page 28: added clarification on the dose of capecitabine (was already in the schema but was added to the text). Added a table with dose modifications for capecitabine<br>• Page 30: table of events - added that pregnancy test is only required for women of child bearing potential only and clarification on type of hepatitis and thyroid function tests required<br><br>**ICF Updates:**<br>• Added the option of research blood work (2 tubes, 10ml each) on C1D2 or C1D3 of study. |
| Version 5 | 16JAN19 | **Protocol Updates:**<br>• Page 18: added clarification in terms of timing for one of the items of inclusion criteria to be consistent with other inclusion criteria.<br>• Page 27: added new information on preparation of nivolumab based on new IB<br>• Page 29: Table of events. Made changes for consistency with the text. Added clarification on time window for some labs and visits. Added clarification for correlative blood samples (at baseline and not screening) to be consistent with the text. |

| | | |
|---|---|---|
| | | • Page 31: added that surveillance can be done by phone or review of medical records to be consistent with table of events<br>• Page 32: removed sentence that was left from the protocol template<br>• Page 33: changed from 30 to 100 days for participation of SAEs after discontinuation of dosing per BMS updated protocol guidelines.<br><br>**ICF Updates:**<br>• Changes made to include Hackensack as a site |
| Version 6 | 16JUL2019 | **Protocol Updates:**<br>• Page 18, 20, Appendix E: Added Quality of Life Questionnaires<br>• Fixed minor administrative discrepancies throughout the protocol<br>• Page 34: Made clarifications regarding dose delays for correlative specimen collection<br>• Page 35: Clarifications on temporary suspension of treatment and dose interruptions<br>• Page 38: Updated the SAE reporting guidelines from BMS for subsites<br>• Page 48: Updated specimen labeling section to be consistent with the lab manual<br><br>**ICF Updates:**<br>• Page 3-4: Added information on biopsy(ies) |
| Version 6.1 | 09Mar2020 | **Protocol Updates:**<br>• PI changed from Dr. Lynce to Dr. Mainor throughout protocol.<br>• **Section 6.7** removed typo “atezolizumab or placebo”<br>• Other Administrative clarifications/corrections made throughout protocol to replace CRO with Multicenter Project Managers and SAE reporting<br>• Updates made to incorporate information from Protocol Clarification letters: |

| | | |
|---|---|---|
| | | • **Inclusion Criteria 2** changed “must have” to “should be” and a Note Added for clarification “(Note: As long as tissue sample is confirmed shipped, the subject will be eligible)”<br>• **Inclusion Criteria 5** typo correction made<br>• **Inclusion Criteria 8** added clarification “(Patients that meet the definition of asymptomatic subclinical hypothyroidism will be eligible to participate)”<br>• **Section 6.3** added clarification “Note: Assessments performed for clinical indications (not exclusively to determine study eligibility) may be used for screening/baseline values even if the studies were done before informed consent was obtained. |

**TABLE OF CONTENTS** **Page**

| | | |
|---|---|---|
| **Table of Contents** | | 8 |
| **Study Synopsis** | | 10 |
| **1.0** | **Background and Justification** | |
| | 1.1 Triple negative breast cancer and residual disease | 12 |
| | 1.2 Immunotherapy as a treatment option for TNBC | 12 |
| | 1.3 The immune milieu in TNBC | 13 |
| | 1.4. Evaluating anti-tumor immunity in the absence of measurable disease | 13 |
| | 1.5 Nivolumab Activity and Pharmacokinetic Profile | 14 |
| | 1.6 Patient reported outcomes (PRO) | 18 |
| | 1.7 Rationale for the Current Study | 19 |
| **2.0** | **Study Objectives** | |
| | 2.1 Primary Objectives | 20 |
| | 2.2 Secondary Objectives | 20 |
| **3.0** | **Study design** | 21 |
| **4.0** | **Subject Population** | |
| | 4.1 Subject Population, Number of Subjects, and Feasibility | 22 |
| | 4.2 Inclusion Criteria | 22 |
| | 4.3 Exclusion Criteria | 24 |
| | 4.4 Additional Study Restrictions | 24 |
| | 4.5 Other Prior and Concomitant Therapy | 26 |
| | 4.6 Removal/Replacement of Subjects from Therapy or Assessment | 27 |
| | 4.7 Multi-Institutional Trial Coordination | 27 |
| **5.0** | **Treatment Plan** | |
| | 5.1 Dose modifications – General Guidelines | 29 |
| | 5.2 Nivolumab | 29 |
| | 5.3. Capecitabine | 32 |
| **6.0** | **Study Procedures** | |
| | 6.1 Table of events | 33 |
| | 6.2 Pre-registration | 34 |
| | 6.3 Screening | 34 |
| | 6.4 Randomization | 35 |
| | 6.5. On Study Day 1 | 35 |
| | 6.6 On Treatment | 35 |
| | 6.7 Treatment discontinuation | 35 |
| | 6.8 Surveillance | 35 |
| **7.0** | **Safety Variables and Toxicity Assessment** | |
| | 7.1 Serious Adverse Events | 36 |
| | 7.2 Adverse Events | 39 |
| | 7.3. Adverse Events Associated with the Use of Immune Checkpoint Inhibitor | 40 |

7.4 Laboratory Test Abnormalities 41
7.5 Pregnancy 42
7.6 Overdose 42
7.7 Other Safety Considerations 42
7.8 Study Monitoring 42
7.9 Protocol Deviations and Violations 44
**8.0 Statistical Considerations**
8.1 Objectives 44
8.2 Sample Size Considerations 45
8.3 Randomization and Stratification Factors 45
8.4 Endpoints 45
8.5 Analysis Plan 46
**9.0 Correlative Research**
9.1 Peripheral immunoscore 47
9.2 Immune contexture in residual breast tumor 47
9.3 Sample labeling overview 48
**10.0 Ethical Considerations**
10.1 Institutional Review Board (IRB) 49
10.2 Ethical Conduct of the Study 49
10.3 Subject Information and Consent 49
10.4 Ethical Consideration for Enrollment 49
10.5 Protection of Patient Confidentiality 49
**11.0 References** 49
**12.0 Appendices** 51
Appendix A: ECOG Performance Status Table) 53
Appendix B: Patient Registration Form 54
Appendix C: Data sharing Plan 55
Appendix D: Management Algorithms 57
Appendix E: Quality of Life Questionnaire (EORTC QLQ-C30 66

#### STUDY SYNOPSIS

| | |
|---|---|
| **Title** | OXEL: A pilot study of immune checkpoint or capecitabine or combination therapy as adjuvant therapy for triple negative breast cancer with residual disease following neoadjuvant chemotherapy |
| **Short Title** | OXEL study |
| **Protocol Number** | 2017-1535 (Georgetown University Parent IRB); BMS CA209-8CL |
| **Clinicaltrials.gov number:** | NCT03487666 |
| **Phase** | Phase 2 |
| **Investigational Agents** | Nivolumab, a PD-1 monoclonal antibody (BMS) |
| **Indication** | Advanced triple negative breast cancers |
| **Study Overview** | This is a pilot phase II trial for patients with triple negative breast cancer. |
| **Study Duration** | 36 months |
| **Study Center(s)** | ➢ Georgetown University, Lombardi Comprehensive Cancer Center Washington, DC<br>➢ MedStar Washington Hospital Center, Washington DC<br>➢ Hackensack University Medical Center, Hackensack, NJ<br>➢ One additional participating center to be determined |
| **Objectives** | <u>Primary objective:</u><br>To assess the immunologic effects of capecitabine, nivolumab or the combo, post neoadjuvant and post-surgery, in the adjuvant setting, in patients with high risk TNBC as defined by presence of residual disease (breast and or LNs) at surgery, in terms of the change of PIS.<br><u>Secondary objectives:</u><br>1) To determine the incidence of toxicity graded using the National Cancer Institute CTCAE v. 4.0 until 30 days after last dose of treatment on trial.<br>2) To determine association between changes in PIS from baseline to week 6 and week 12 and clinical outcome variables (DRFS and OS at 3-years).<br>3) Translational endpoints:<br>a) circulating tumor DNA (ct-DNA) during treatment<br><u>Exploratory objectives:</u><br>a) To describe the immune contexture in residual tumors of patients with high risk TNBC after receiving neoadjuvant chemotherapy<br>b) Tissue banking for future studies (NGS of initial biopsy and surgical specimen), plasma, PBMCs |
| **Number of Subjects** | The anticipated number of patients will be 45. |

| | |
|---|---|
| **Diagnosis and Main Inclusion and Exclusion Criteria** | Key Inclusion Criteria: Biopsy proven TNBC (ER and PR defined as ≤5% cells stain positive), HER2 negativity defined as (IHC0, 1+ without in situ hybridization (ISH) HER2/neu chromosome 17 ratio OR IHC 2+ and ISH HER2/neu chromosome 17 ratio non-amplified with ratio <2.0 and if reported average HER2 copy number <6 signals/cells); residual disease of ≥1.0 cm of the primary tumor and/or node positive disease measuring ≥ 0.5cm; receipt of neoadjuvant taxane +/- anthracycline; platinum use allowed; must have completed definitive resection of primary tumor; signed ICF; age ≥18.<br>Key Exclusion Criteria: Stage IV disease; previous exposure to capecitabine, fluorouracil or immunotherapy with anti-PD1, anti PDL1 or anti-CTLA4; active autoimmune disease requiring systemic treatment in the past 2 years; tuberculosis, hepatitis B or C, HIV or other active infection; chronic use of systemic steroids or pregnancy. |
| **Study Design** | Pilot phase II open-label three arm randomized trial of nivolumab, capecitabine or combination as adjuvant therapy for 45 patients with residual TNBC after adequate neoadjuvant chemotherapy followed by definitive surgery. Patients are randomly assigned to 1 of 3 treatment arms: ARM A – Nivolumab 360 mg iv q3weeks for x 6 cycles; ARM B - Capecitabine 1250mg/$m^2$ bid D1-D14 q3 weeks x 6 cycles; ARM C - Nivolumab 360mg iv q3weeks + Capecitabine 1250mg/$m^2$ bid D1-D14 q3 weeks x 6 cycles. We will enroll 15 patients per arm (45 totally for 3 arms). |
| **Duration of therapy** | All drugs will be administered for 18 weeks, or until disease progression or unacceptable toxicity is observed. |
| **Sample size considerations** | The primary objective of the study is to assess the change in PIS, the Peripheral ImmunoScore, at 6 weeks (=2 cycles) from baseline, in each arm. The sample size of 15 per arm (45 totally for 3 arms) is mainly based on the feasibility of patient accrual. A sample size of 15 per arm will have about 85% power to detect an effect size of 1 (the difference of the change in PIS from baseline to week 6 between two arms divided by the standard deviation) at 5% significance level. |

#### 1.0 BACKGROUND AND JUSTIFICATION

##### 1.1. Triple negative breast cancer and residual disease

Breast cancer (BC) is the most common cancer in women in the United States, with an estimated 252,710 new cases of invasive BC anticipated in 2017[1]. Overall, triple negative breast cancer (TNBC) accounts for 10-15% of invasive BC cases and it is associated with a worse prognosis and limited treatment options compared to the other subtypes[2,3]. Patients with TNBC and residual disease at the time of surgery after completing neoadjuvant chemotherapy constitute a group of patients at a high risk of recurrence. Long-term follow-up of neoadjuvant studies consistently demonstrates worse clinical outcomes in patients with TNBC who did not achieve pathologic complete response (pCR), with only 35-40% remaining free of recurrence at 2 years[4-6]. Until recently, the standard of care for patients with early stage TNBC who do not achieve pCR was observation or participation in a clinical trial. More recently, adjuvant capecitabine became an option after the CREATE-X study revealed that the addition of adjuvant capecitabine prolonged disease-free survival (DFS) and overall survival (OS) among patients with HER2-negative BC who had residual invasive disease on pathological testing[7]. This benefit was more striking in patients with TNBC (DFS 69.8% in the capecitabine group vs 56.1% in the control group, HR 0.58; 95% CI, 0.39 to 0.87; and OS 78.8% vs 70.3%, HR 0.52; 95% CI, 0.30 to 0.90).

##### 1.2. Immunotherapy as a treatment option for TNBC

Immunotherapy has shifted the treatment paradigm for many malignancies, but not all cancer types have enjoyed a clinically meaningful response from checkpoint blockade. In BC, most work to date with immunotherapy has focused on TNBC given the greater frequency of tumor infiltrating lymphocytes (TILs)[8]. The overall response rate (ORR) in heavily pretreated patients with TNBC who received pembrolizumab single agent was 18.5%[9]. Other trials in TNBC either in the metastatic[10] or neoadjuvant setting[11], have shown that the combination of immune checkpoint inhibitors plus chemotherapy may be a more promising approach. In the phase I clinical trial of atezolizumab and nabpaclitaxel, in patients with metastatic disease those treated in first line derived the greatest benefit, when compared to patients treated in second or third line (ORR 67% vs 25-29% in 1st line versus 2nd/3rd line respectively)[9]. These findings confirm the overall impression that when chemoimmunotherapy is used earlier in TNBC, the outcome is better. They also support the notion that, in TNBC, the addition of chemotherapy to a checkpoint inhibitor is a promising strategy, contrary to what is observed in less chemosensitive diseases such as melanoma.

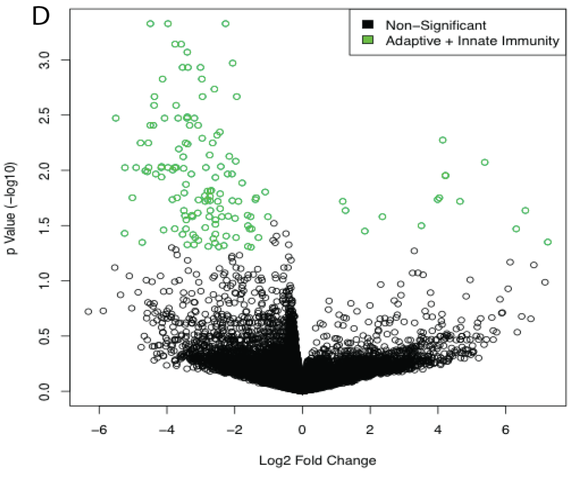

**Figure 1**. Genome-wide shRNA library screen in EO771 TNBC tumors growing in WT or SCID C57Bl/6 mice reveals genes whose knockdown either reduces (left, upper) or increases (right, upper) the representation of the targeted tumor cells in 1

#### 1.3. The immune milieu in TNBC

It is unlikely that immunotherapy alone or in combination with chemotherapy will be effective in all patients with residual TNBC after neoadjuvant chemotherapy. Thus, we need to characterize the immune contexture of the residual tumors in order to select patients who will benefit additional therapy including immunotherapy. Many tools are available to describe the immune contexture in the tumor microenvironment, using powerful technologies such as multiplex immunohistochemistry (IHC), genome-wide association studies (GWAS) and mass cytometry[12]. Recently we employed a genome-wide shRNA screen to identify 709 genes that selectively regulated adaptive anti-tumor immunity in a syngeneic, functionally TNBC model and focused on five genes (CD47, TGFβ1, Sgpl1, Tex9 and Pex14) with the greatest impact as biomarkers of resistance[13] (Fig. 1). We have used this same approach to identify genes that regulate resistance to PD-1 antibody directed immune attack in the same TNBC model (not shown). While work done using The Cancer Genome Atlas (TCGA) data to characterize immune cell infiltrates in TNBC is valuable and foundational[14] it may not accurately reflect the immune milieu of tumors resistant to chemotherapy. This is the rationale to the evaluation of the immune contexture in tumors that are resistant to neoadjuvant chemotherapy, in order to guide future adjuvant therapy approaches for these patients at a high risk of recurrence.

#### 1.4. Evaluating anti-tumor immunity in the absence of measurable disease

A significant challenge in evaluating the effects of experimental adjuvant therapies is the lack of measurable disease to assess therapeutic effects. In this setting, when tumor is not detectable such as in patients with early BC who have undergone surgical resection of their tumor, the identification of biomarkers in the peripheral blood – the most readily available tissue for sampling – to assess effects of the intervention is intuitively attractive. Immune cell subsets within peripheral blood mononuclear cells (PBMC) have been analyzed to determine if a peripheral immunoscore could have any prognostic significance for patients undergoing immunotherapy. Although to date there is no validated FDA-approved circulatory immunological biomarker for patients with breast cancer, the refined peripheral immunoscore (PIS) defined by Farsaci et al[15] (Table 1) was validated in a group of patients with metastatic BC receiving docetaxel and immunotherapy, and shown to be predictive of progression free survival (PFS). When compared to predefined analyses of "classic" immune cell types (CD4, CD8, natural killer cells, regulatory T cells, myeloid-derived suppressor cells, and ratios) that revealed no differences in PFS for either arm

| Criterion 1: Classic cell types | | Criterion 2: Phenotypes of refined cell subsets reflecting immune function | | |
|---|---|---|---|---|
| Description | Expected effect on PFS | Description | Expected effect on PFS | Reference |
| % CD4 | Positive | % Central memory CD4[a] | Positive | (19) |
| | | % CD4 expressing ≥2 suppressive markers[b] | Negative | (20, 21) |
| % CD8 | Positive | % Central memory CD8[a] | Positive | (19) |
| | | % CD8 expressing ≥2 suppressive markers[b] | Negative | (20, 21) |
| % Treg[c] | Negative | % Treg CD49d$^-$ | Negative | (22, 23) |
| % MDSC[d] | Negative | % MDSC Lin$^-$ [e] | Negative | (24, 25) |
| % NK[f] | Positive | % NK CD56$^{br}$CD16$^-$ | Negative | (26, 27) |
| Ratio CD4:Treg | Positive | Ratio central memory[a] CD4:Treg CD49d$^-$ | Positive | |
| Ratio CD8:Treg | Positive | Ratio central memory[a] CD8:Treg CD49d$^-$ | Positive | |
| Ratio CD4:MDSC | Positive | Ratio central memory[a] CD4:MDSC Lin$^-$ | Positive | |
| Ratio CD8:MDSC | Positive | Ratio central memory[a] CD8:MDSC Lin$^-$ | Positive | |
| | | Ratio central memory[a] CD4:NK CD56$^{br}$CD16$^-$ | Positive | |
| | | Ratio central memory[a] CD8:NK CD56$^{br}$CD16$^-$ | Positive | |

NOTE: Frequencies of PBMC subsets represented as a percentage of total PBMCs. [a], CD45RA$^-$CCR7$^+$; [b], CTLA-4, PD-1, Tim-3, 2B4; [c], CD4$^+$CD25$^+$FoxP3$^+$CD127$^-$; [d], CD3$^-$CD16$^-$CD19$^-$CD20$^-$CD56$^-$CD33$^+$HLA-DR$^{-/low}$; [e], CD14$^-$CD15$^-$; [f], CD3$^-$CD56$^+$.

Table 1. Refined peripheral immunoscore

(docetaxel alone or docetaxel with vaccine), predefined analyses of refined immune cell subsets for which a biologic function had been previously reported revealed statistically significant improved PFS (*P* < 0.001) for those receiving docetaxel plus immunotherapy (Fig 2). Based on these results, we selected for our study the PIS as a surrogate of clinical benefit and a new strategy to identify patients with BC who may benefit from immunotherapy to eliminate residual micrometastasis.

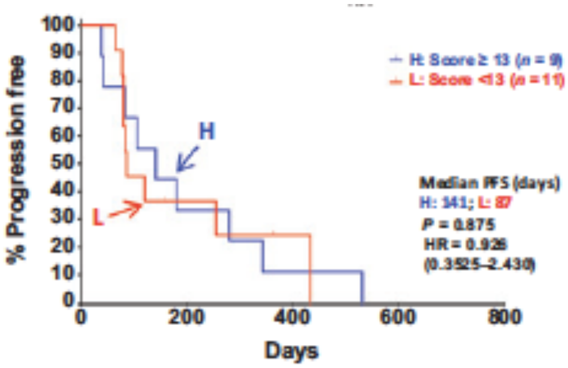

Figure 2.a Docetaxel alone

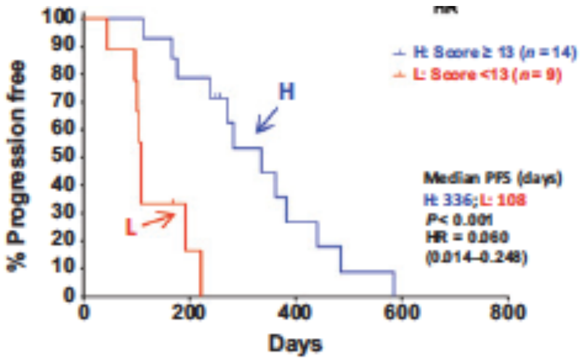

Figure 2.b Docetaxel and vaccine

#### 1.5 Nivolumab Activity and Pharmacokinetic Profile

##### 1.5.1. Introduction

Nivolumab (also referred to as BMS-936558, MDX1106, or ONO-4538) is a human monoclonal antibody (HuMAb; immunoglobulin G4 [IgG4]-S228P) that targets the programmed death-1 (PD-1) cluster of differentiation 279 (CD279) cell surface membrane receptor. PD-1 is a negative regulatory molecule expressed by activated T and B lymphocytes[16]. Binding of PD-1 to its ligands, programmed death–ligands 1 (PD-L1) and 2 (PD-L2), results in the down-regulation of lymphocyte activation. Inhibition of the interaction between PD-1 and its ligands promotes immune responses and antigen-specific T-cell responses to both foreign antigens as well as self-antigens. Nivolumab is expressed in Chinese hamster ovary (CHO) cells and is produced using standard mammalian cell cultivation and chromatographic purification technologies. The clinical study product is a sterile solution for parenteral administration. OPDIVO™ (nivolumab) is approved for the treatment of several types of cancer in multiple regions including the United States (US, Dec-2014), the European Union (EU, Jun-2015), and Japan (Jul-2014). Nivolumab is also being investigated in various other types of cancer as monotherapy or in combination with other therapies, and as single-dose monotherapy for the treatment of sepsis.

##### 1.5.2. Nonclinical Studies

Nivolumab has been shown to bind specifically to the human PD-1 receptor and not to related members of the CD28 family[17,18]. Nivolumab inhibits the interaction of PD-1 with its ligands, PD-L1 and PD-L2, resulting in enhanced T-cell proliferation and interferon-gamma (IFN-γ) release in vitro[19-21]. Nivolumab binds with high affinity to activated human T-cells expressing cell surface PD-1 and to cynomolgus monkey PD-1.2 In a mixed lymphocyte reaction (MLR), nivolumab promoted a reproducible concentration-dependent enhancement of IFN-γ release[22]. In intravenous (IV) repeat-dose toxicology studies in cynomolgus monkeys, nivolumab was well tolerated at doses up to 50 mg/kg, administered weekly for 5 weeks, and at doses up to 50 mg/kg, administered twice weekly for 27 doses. While nivolumab alone was well tolerated in cynomolgus monkeys, combination studies have highlighted the potential for enhanced toxicity when combined with other immunostimulatory agents[23].
In addition, an enhanced pre- and postnatal development (ePPND) study in pregnant cynomolgus monkeys with nivolumab was conducted[24]. Administration of nivolumab at up to 50 mg/kg 2QW was well tolerated by pregnant monkeys; however, nivolumab was determined to be a selective developmental toxicant when administered from the period of organogenesis to parturition at $\geq$ 10 mg/kg (area under the concentration-time curve [AUC] from time zero to 168 hours [AUC(0-168 h)] 117,000 μg•h/mL). Specifically, increased developmental mortality (including late gestational fetal losses and extreme prematurity with associated neonatal mortality) was noted in the absence of overt maternal toxicity. There were no nivolumab-related changes in surviving infants tested throughout the 6-month postnatal period. Although the cause of these pregnancy failures was undetermined, nivolumab-related effects on pregnancy maintenance are consistent with the established role of PD-L1 in maintaining fetomaternal tolerance in mice[25].

<u>1.5.3. Effects in Humans</u>
The pharmacokinetics (PK), clinical activity, and safety of nivolumab have been assessed in subjects with non-small cell lung cancer (NSCLC), melanoma, clear-cell renal cell carcinoma (RCC), classical Hodgkin Lymphoma (cHL), urothelial carcinoma (UC), squamous cell carcinoma of the head and neck (SCCHN), in addition to other tumor types. Nivolumab monotherapy is approved in multiple regions, including the US and EU, for unresectable or metastatic melanoma, previously treated metastatic NSCLC, previously treated advanced RCC, previously treated relapsed or refractory cHL, and previously treated advanced or metastatic UC; it is also approved for the treatment of previously treated recurrent or metastatic SCCHN in the US. In addition, nivolumab has been approved for use in combination with ipilimumab for unresectable melanoma in multiple countries, including the US and EU. Nivolumab is being investigated both as monotherapy and in combination with chemotherapy, targeted therapies, and other immunotherapies for the treatment of several types of cancer.

<u>1.5.4. Clinical Pharmacokinetics</u>

The PK of nivolumab was studied in subjects with cancer over a dose range of 0.1 to 20 mg/kg administered as a single dose or as multiple doses of nivolumab every 2 or 3 weeks. Nivolumab clearance (CL) decreases over time, with a mean maximal reduction (% coefficient of variation [CV%]) from baseline values of approximately 24.5% (47.6%) resulting in a geometric mean steady state clearance (CLss) (CV%) of 8.2 mL/h (53.9%); the decrease in CLss is not considered clinically relevant. The geometric mean volume of distribution at steady state (Vss) was 6.8 L (27.3%), and geometric mean elimination half-life (t1/2) was 25 days (77.5%). Steady-state concentrations of nivolumab were reached by 12 weeks when administered at 3 mg/kg Q2W, and systemic accumulation was approximately 3.7-fold. The exposure to nivolumab increased dose proportionally over the dose range of 0.1 to 10 mg/kg administered every 2 weeks. Additionally, nivolumab has a low potential for drug-drug interactions. The clearance of nivolumab increased with increasing body weight. The PPK analysis suggested that the following factors had no clinically important effect on the CL of nivolumab: age (29 to 87 years), gender, race, baseline LDH, PD-L1 status, solid tumor type, baseline tumor size, and hepatic impairment. Although ECOG status, baseline glomerular filtration rate (GFR), albumin, and body weight had an effect on nivolumab CL, the effect was not clinically meaningful. PPK analysis suggest that nivolumab CL in subjects with cHL was approximately 32% lower relative to subjects with NSCLC; however, the lower CL in cHL subjects was not considered to be clinically relevant as nivolumab exposure was not a significant predictor for safety risks for these patients. When nivolumab is administered in combination with ipilimumab, the CL of nivolumab was increased by 29%, whereas there was no effect on the clearance of ipilimumab. PPK and exposure response analyses have been performed to support use of nivolumab 240 mg Q2W, 360 mg Q3W, and 480 mg Q4W dosing regimens in subjects with cancer in addition to the 3 mg/kg Q2W regimen. A flat dose of nivolumab 240 mg Q2W was selected since it is identical to a dose of 3 mg/kg for subjects weighing 80 kg, the observed median body weight in nivolumab treated cancer patients, while the nivolumab 360 mg Q3W and 480 mg Q4W regimens allow flexibility of dosing with less frequent visits and in combination with other agents using alternative dosing schedules to Q2W. Using a PPK model, the overall distributions of nivolumab exposures (Cavgss, Cminss, Cmaxss, and Cmin1) are comparable after treatment with either nivolumab 3 mg/kg or 240 mg Q2W. Following nivolumab 360 mg Q3W and 480 mg Q4W, Cavgss are expected to be similar to those following nivolumab 3 mg/kg or 240 mg Q2W, while Cminss are predicted to be 6% and ~16% lower, respectively, and are not considered to be clinically relevant. Following nivolumab 360 mg Q3W and 480 mg Q4W, Cmaxss are predicted to be approximately ~23% and ~43% greater, respectively, relative to that following nivolumab 3 mg/kg Q2W dosing. However, the range of nivolumab exposures (median and 90% prediction intervals) following administration of 240 mg flat Q2W, 360 mg Q3W, and 480 mg Q4W regimens across the 35 to 160 kg weight range are predicted to be maintained well below the corresponding exposures observed with the well tolerated 10 mg/kg nivolumab Q2W dosing regimen. In this study we will use the flat dose of 360mg Q3W. Although this is a less frequent dosing regimen, it will allow combination of nivolumab with the capecitabine schedule. This nivolumab dosing regimen was selected using several simulation models to provide approximately equivalent exposures following administration of nivo 3 mg/kg Q2W. The models predicted that following administration of nivo 360 mg Q3W exposures are expected to be similar to those following nivo 3 mg/kg or 240 mg Q2W.

###### 1.5.5. Clinical Efficacy

Nivolumab has demonstrated durable responses exceeding 6 months as monotherapy and in combination with ipilimumab in several tumor types, including NSCLC, melanoma, RCC, cHL, SCLC, gastric cancer, SCCHN, urothelial cancer, HCC, and CRC. In confirmatory trials, nivolumab as monotherapy demonstrated a statistically significant improvement in OS as compared with the current standard of care in subjects with advanced or metastatic NSCLC, unresectable or metastatic melanoma, advanced RCC, or SCCHN. Nivolumab in combination with ipilimumab improved PFS and ORR over ipilimumab alone in subjects with unresectable or metastatic melanoma.

###### 1.5.6. Clinical Safety

The overall safety experience with nivolumab, as a monotherapy or in combination with other therapeutics, is based on experience in approximately 16,900 subjects treated to date. For monotherapy, the safety profile is similar across tumor types. There is no pattern in the incidence, severity, or causality of AEs to nivolumab dose level. In Phase 3 controlled studies, the safety profile of nivolumab monotherapy is acceptable in the context of the observed clinical efficacy, and manageable using established safety guidelines. Clinically relevant AEs typical of stimulation of the immune system were infrequent and manageable by delaying or stopping nivolumab treatment and timely immunosuppressive therapy or other supportive care. Based on preliminary data from an ongoing Phase 1 study, there have been no unexpected safety findings to date in patients with sepsis who received a single dose of nivolumab monotherapy. In several ongoing clinical trials, the safety of nivolumab in combination with other therapeutics such as ipilimumab, cytotoxic chemotherapy, anti-angiogenics, and targeted therapies is being explored. Most studies are ongoing and, as such, the safety profile of nivolumab combinations continues to evolve. The most advanced combination under development is nivolumab + ipilimumab, which is approved in subjects with unresectable or metastatic melanoma, and being studied in multiple tumor types. Results to date suggest that the safety profile of nivolumab+ipilimumab combination therapy is consistent with the mechanisms of action of nivolumab and ipilimumab. The nature of the AEs is similar to that observed with either agent used as monotherapy; however, both frequency and severity of most AEs are increased with the combination. Nivolumab at 360mg q3w has been shown to be safe in combination with capecitabine and oxaliplatin in patients with G/GEJ cancers (Kang YK, et al. Annals of Oncology 2017).

There are other ongoing studies evaluating the combination of capecitabine and nivolumab (NCT02746796; NCT03006705).

#### 1.6 Patient reported outcomes (PRO)

The assessment of patient-reported outcomes (PROs) in clinical research provides important insight into how therapies impact the daily lives of patients. Patient reports regarding health related quality of life have proven more comprehensive than provider-collected data in breast cancer patients (Oberguggenberger et al 2011).

##### 1.6.1 EORTC QLQ-C30

In this study patient-reported disease related symptoms and health-related quality of life (HRQoL) will be evaluated using the validated EORTC QLQ-C30 questionnaire. The EORTC QLQ-C30 questionnaire was developed to assess HRQoL and is the most commonly used cancer-specific tool in oncology.
The EORTC QLQ-C30 comprises 30 questions designed for all cancer types. Questions can be grouped into the following scales:

- 5 multi-item functional scales (physical, role, emotional, cognitive and social)
- 3 multi-item symptom scales (fatigue, pain, nausea vomiting)
- A 2-item global QoL scale
- 5 single items assessing the following common cancer symptoms: dyspnea, loss of appetite, insomnia, constipation, diarrhea
- 1 item on the financial impact of the disease.

All the EORTC scales range from 0 to 100 (through transformation of scores). A high scale score represents a higher response level. Thus a high score for a functional scale represents a high / healthy level of functioning, while a high score for a symptom scale / item represents a high level of symptomatology / problems.

##### 1.6.2 Administration of PRO questionnaires

Paper-based EORTC QLQ-C30 will be administered at baseline (prior to randomization once eligibility is confirmed), at 6 weeks visit and at the end of treatment.
Each center must allocate the responsibility for the administration of the questionnaires to a specific individual (e.g., a research nurse, study coordinator) and if possible assign a back-up person to cover if that individual is absent. The significance and relevance of the data need to be explained carefully to participating patients so that they are motivated to comply with data collection.
The instructions for completion of the PRO questionnaires are as follows:

- They must be completed prior to any other study procedures (following informed consent) and before discussion of disease progress to avoid biasing the patient’s responses to the questions. They must be completed in private by the patient
- The patient should be given sufficient time to complete at their own speed
- The patient should not receive help from relatives, friends or clinic staff to answer the questionnaire.
- On completion of the questionnaire it should be handed back to the person responsible for questionnaires who should check for completeness

 Only one answer should be recorded for each question

#### 1.7. Rationale for Current Study

Recent results presented at ASCO 2017 indicated that a subset of patients with early stage triple negative breast cancer benefit from immune checkpoint inhibitors[11]. It is of critical importance to develop tools to help identify which patients benefit from this strategy. As discussed above, we currently don't have any validated biomarker that allow us to detect this subset of patients. Tumor biopsies at different timepoints are not an option in this population when the only tumor available is the residual tissue removed at the time of surgery. The presence of circulating tumor cells after chemotherapy has been shown to have a negative impact on DFS and OS (Rack B, et al. JNCI 2014) but has not been used to predict benefit from immunotherapy. The presence of PD-L1 expression on tumor cells has been associated with objective responses in metastatic cancer (Topalian, NEJM) but not been evaluated in patients who have completed adjuvant therapy. Presence of TILs has been associated with increased PD-L1 infiltrate, but the association between increased TILs and response to immune checkpoint therapy has not yet been established. For this pilot study, we selected the refined peripheral immunoscore (PIS)[15] defined by Farsaci based on it's being the closest to the particular scenario proposed on this study. His group demonstrated that an immunoscore of refined immune cell subsets showed significant prognostic value in PFS for metastatic breast cancer patients receiving docetaxel plus vaccine ($P < 0.001$) and for metastatic prostate cancer patients receiving radionuclide plus vaccine ($p=0.004$), but not on the same groups when treated with chemotherapy alone.

This pilot study will provide preliminary data regarding the role of PIS in predicting the benefit of immune checkpoint inhibition with or without chemotherapy for high risk patients with TNBC and residual disease after effective neoadjuvant chemotherapy. If the peripheral immunoscore fails as a biomarker, this pilot study would still be very relevant as we will be collecting other exploratory biomarkers in this unique population.

#### 2.0 STUDY OBJECTIVES

2.1 Primary objective:

1) To assess the immunologic effects of capecitabine, nivolumab or the combo, post neoadjuvant and post-surgery, in the adjuvant setting, in patients with high risk TNBC as defined by presence of residual disease (breast and or LNs) at surgery, in terms of the change of PIS.
   a) We hypothesized that among patients with TNBC and residual disease at the time of surgery, the change of the Peripheral ImmunoScore (PIS) from baseline to week 6 will be higher among those who receive post-surgery immunotherapy (ARM A and C), compared to those who receive post-surgery chemotherapy alone (ARM B).

2.2 Secondary objectives:

1). To determine the incidence of toxicity graded using the National Cancer Institute CTCAE v. 4.0 until 30 days after last dose of treatment on trial.

2). To determine association between changes in PIS from baseline to week 6 and week 12 and clinical outcome variables (DRFS and OS at 3-years)). After end of study visit, clinical follow up or telephone communication every 3months.

3). Translational endpoints:

   a) Circulating-tumor DNA (ct-DNA)

Exploratory objectives:

   c) To describe the immune contexture in residual tumors of patients with high risk TNBC after receiving neoadjuvant chemotherapy
   d) Tissue banking for future studies (NGS of initial biopsy and surgical specimen)
   e) To explore the impact of nivolumab on patient-reported outcomes (PROs) of health-related quality of life (HRQoL), as measured by the European Organization for Research and Treatment of Cancer (EORTC) EORTC Quality of Life Questionnaire Core 30 (QLQ-C30) disease related multi-item symptom and functional scales.

#### 3.0 STUDY DESIGN

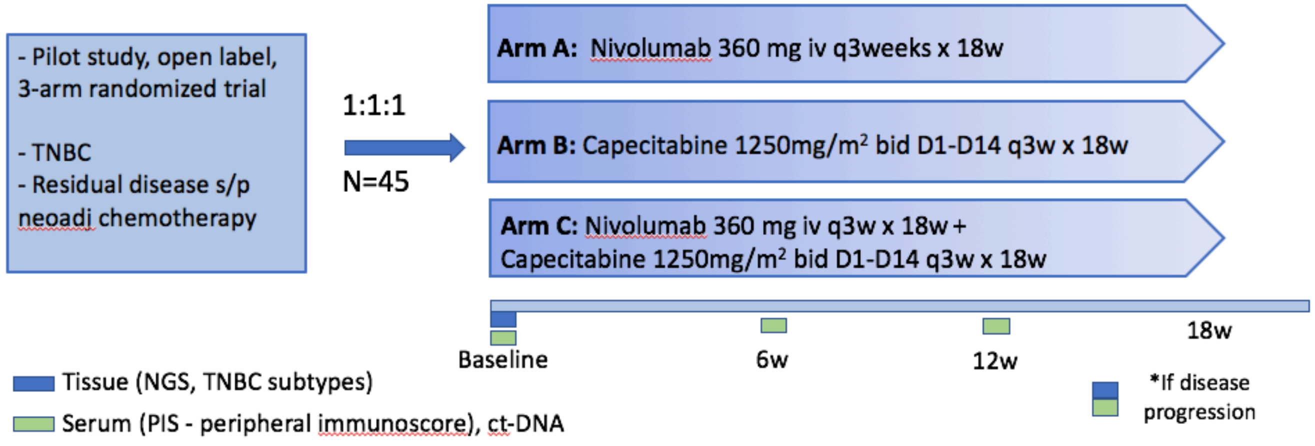

Figure 3. Study design

The OXEL "Opdivo, XELoda or combination therapy as adjuvant therapy for triple negative breast cancer with residual disease following neoadjuvant chemotherapy" is a pilot phase II open-label three arm randomized trial of nivolumab, capecitabine or combination as adjuvant therapy for 45 patients with residual TNBC after adequate neoadjuvant chemotherapy followed by definitive surgery. We will enroll 15 patients per arm (45 totally for 3 arms).

Patients are randomly assigned to 1 of 3 treatment arms:

- ARM A – Nivolumab 360 mg iv q3weeks for x 6 cycles;
- ARM B - Capecitabine 1250mg/m$^2$ bid D1-D14 q3 weeks x 6 cycles;
- ARM C - Nivolumab 360mg iv q3weeks + Capecitabine 1250mg/m$^2$ bid D1-D14 q3 weeks x 6 cycles.

Nivolumab (Opdivo®) is a human programmed death receptor-1 (PD-1) antibody currently approved in different diseases. Capecitabine (Xeloda®) was selected for ARM B (as contemporary control arm) given the recent results from CREATE-X trial[7] and the increasing use by the community (feasibility). Importantly, available data from other scenarios indicates that capecitabine does not have immunosuppressive effects[27]. Given capecitabine low myelosuppressive potential, it was felt to be a reasonable chemotherapy partner with the checkpoint inhibitor in this setting (ARM C). Finally, there is safety data regarding combination of nivolumab and capecitabine in patients with G/GEJ tumors (Kang YK, et al. Annals of Oncology 2017).

**4.0 SUBJECT POPULATION**

**4.1 Subject Population, Number of Subjects and Feasibility**

4.1.1 Subject Population

Patients will have triple negative breast cancer. Enrolled patients will have completed standard neoadjuvant chemotherapy and will have residual disease at the time of definitive breast surgery. Patients will have completed adequate local therapy.

4.1.2 Number of Subjects

45 patients will be enrolled with advanced triple negative breast cancer. Patients who don't have evaluable samples as per protocol will be replaced.

**4.2 Inclusion Criteria**

1. Biopsy proven TNBC:
    - ER- and PR- defined as ≤5% cells stain positive
    - HER2 negativity defined as:
        - IHC 0, 1+ without in situ hybridization (ISH) HER2/neu chromosome 17 ratio OR
        - IHC 2+ and ISH HER2/neu chromosome 17 ratio non-amplified with ratio less than 2.0 and if reported average HER2 copy number < 6 signals/cells
2. Residual disease of 1.0 cm at least of the primary tumor and/or node positive disease (at least ypN1) and/or Residual Cancer Burden (RBC) classification II or III. Tissue availability needs to be confirmed and should be received by the lead institution by day 1 of study treatment. (Note: As long as tissue sample is confirmed shipped, the subject will be eligible)
3. Patients must have completed neoadjuvant chemotherapy; patients must NOT have received capecitabine as part of their neoadjuvant therapy regimen. Acceptable preoperative regimens include an anthracycline or a taxane, or both. Participants who received preoperative therapy as part of a clinical trial may enroll. Participants may not have received adjuvant chemotherapy after surgery prior to randomization. Carboplatin-containing neoadjuvant chemotherapy is also allowed). Patients who cannot complete all planned neoadjuvant treatment cycles for any reason are considered high risk and therefore are eligible for the study if they have residual disease.
4. Recovery of all toxicities from previous therapies to at least grade 1, except alopecia and ≤ grade 2 neuropathy which are allowed.
5. Must have completed definitive resection of primary tumor and have no evidence of unresected or metastatic disease at the time of study entry. For those who don't require radiotherapy, the most recent surgery for breast cancer must have been completed at least 14 days prior to day 1 of study treatment, but no more than 84 days prior to study registration.
    - Negative margins for both invasive and ductal carcinoma in situ (DCIS) are desirable,

however patients with positive margins may enroll if the treatment team believes no further surgery is possible and patient has received radiotherapy; patients with margins positive for lobular carcinoma in situ (LCIS) are eligible

- Either mastectomy or breast conserving surgery (including lumpectomy or partial mastectomy) is acceptable
- Sentinel node biopsy post neoadjuvant chemotherapy (i.e. at the time of definitive surgery) is allowed; axillary dissection is encouraged in patients with lymph node involvement, but is not mandatory
- Whole breast radiotherapy is required for participants who underwent breast-conserving therapy, including lumpectomy or partial mastectomy. Participants must have completed radiotherapy at least 14 days prior (but no more than 84 days prior) to day 1 of study treatment.
- Post-mastectomy radiotherapy is required for all participants with a primary tumor ≥ 5 cm or involvement of ≥ 4 lymph nodes. For participants with primary tumors < 5 cm or with < 4 involved lymph nodes, provision of post-mastectomy radiotherapy is at the discretion of the treating physician.

6. ECOG PS 0-1
7. Patients must not be planning to receive concomitantly other biologic therapy, hormonal therapy, other chemotherapy, surgery, radiation therapy or other anti-cancer therapy while receiving treatment on this protocol.
8. At the time of registration (randomization), patients must have the following laboratory results (obtained within 28 days prior to registration):
   a. A serum TSH prior to registration to obtain a baseline value.
   b. Patients must have adequate bone marrow function as evidenced by all of the following:
      - ANC ≥ 1,500 microliter (mcL);
      - Platelets ≥ 100,000/mcL;
      - Hemoglobin ≥ 9 g/dL.
   c. Patients must have adequate hepatic function as evidenced by the following:
      - Total bilirubin ≤ 1.5 x institutional upper limit of normal (IULN) (except Gilbert's Syndrome, who must have a total bilirubin < 3.0 mg/dL), and
      - SGOT (AST) or SGPT (ALT) and alkaline phosphatase ≤ 2.5 x IULN.
   d. Patients must have adequate renal function as evidenced by ONE of the following:
      - Serum creatinine ≤ IULN OR
      - Measured or calculated creatinine clearance ≥ 60 mL/min.
   e. Normal thyroid function or stable on hormone supplementation (Patients that meet the definition of asymptomatic subclinical hypothyroidism will be eligible to participate)
   f. Women of childbearing potential (WOCBP) must have a negative urine or serum pregnancy test within 28 days prior to registration and within 24h prior to the start of nivolumab. In addition, women of childbearing potential must agree to have a pregnancy test every 4 weeks while on nivolumab.
9. Signed ICF

10. Age ≥18

**4.3 Exclusion criteria**

1. Stage IV disease
2. Receipt of adjuvant chemotherapy
3. Diagnosis of other invasive cancer except for adequately treated cervix cancer or skin cancer, or more than 5 years since other diagnosis of invasive cancer without current evidence of disease
4. Previous exposure to capecitabine, fluorouracil or immunotherapy with anti-PD1, anti-PDL1, anti-CTLA4 or similar drugs.
5. Active autoimmune disease that has required systemic treatment in the past 2 years; replacement therapy is not considered a form of systemic therapy. Autoimmune diseases include but are not limited to myasthenia gravis, myositis, autoimmune hepatitis, systemic lupus erythematosus, rheumatoid arthritis, inflammatory bowel disease, vascular thrombosis associated with antiphospholipid syndrome, Wegener's granulomatosis, Sjogren's syndrome, Guillain-Barre syndrome, multiple sclerosis, vasculitis or glomerulonephritis.
6. TB, active hepatitis B (defined as having a positive hepatitis B surface antigen [HBsAg]), active hepatitis C (defined as positive test for hepatitis C viral load by polymerase chain reaction [PCR]) or other active infection. Patients with past hepatitis B virus (HBV) infection or resolved HBV infection (defined as having a negative HBsAg test and a positive antibody to hepatitis B core antigen [anti-HBc] antibody test) are eligible. Patients with positive hepatitis C antibody and negative quantitative hepatitis C by PCR are eligible. Patients who have completed curative therapy for HCV are eligible. Patients with known HIV infection are eligible if they meet each of the following 3 criteria: CD4 counts ≥ 350 $mm^3$; serum HIV viral load of < 25,000 IU/ml and treated on a stable antiretroviral regimen.
7. History of (non-infectious) pneumonitis that required steroids or evidence of active pneumonia
8. Uncontrolled disease
9. Chronic use of systemic steroids
10. Live vaccine within 30 days prior to registration.
11. Incapacity to provide consent or to follow clinical trial procedures
12. Pregnancy, lactation, or planning to be pregnant

<u>Patients with microsatellite unstable tumors will not be excluded as immunotherapy as adjuvant therapy is not standard for these patients but we will prospective collect this data.</u>

#### 4.4 Additional Study Restrictions

##### 4.4.1 Other Anticancer Therapy

For purposes of this protocol, anti-tumor treatment may be defined as, but is not limited to, anti-cancer agents (endocrine therapy, cytotoxic chemotherapy, immunotherapy, or biologic therapy), radiotherapy, and investigational agents. An investigational agent is any drug or therapy not currently approved for use in humans. No other anticancer therapy is permitted during the course of the study treatment for any patient (the patient can receive a stable dose of corticosteroids during the study as long as these were started at least 4 weeks prior to enrollment and the total daily dose does not exceed 10mg of prednisone, or the equivalent, per exclusion criteria above). If the patient discontinues study treatment, this restriction no longer applies, however the patient will remain enrolled in the study for the purpose of collecting subsequent outcomes. Radiotherapy is not allowed.

##### 4.4.2 Vaccines

Live vaccine administration is not permitted within 30 days prior to the first dose of study treatment and while participating in the study. Examples of live vaccines include, but are not limited to, the following: measles, mumps, rubella, chicken pox, yellow fever, rabies, bacilli Calmette-Guerin (BCG), and typhoid (oral) vaccine. Seasonal influenza vaccines for injection are generally killed virus vaccines and are allowed. Intranasal influenza vaccines (eg, Flu-Mist®) are live attenuated vaccines, however, and are not allowed.

##### 4.4.3 Birth Control

Birth control should be used from the signing of the patient consent form and for 6 months following the last dose of nivolumab and capecitabine. Acceptable methods of birth control include:

- At least one highly effective form of contraception, defined as a contraceptive method with a failure rate of less than 1% per year when used consistently and correctly.
- Permanent sterilization, defined as hysterectomy, bilateral salpingectomy, bilateral oophorectomy, or bilateral orchiectomy
- Postmenopausal, defined as a female patient or sexual partner >45 years of age who has not menstruated for at least 12 consecutive months
- Total sexual abstinence

In addition, men must not donate sperm during nivolumab and capecitabine therapy and for seven months after receiving the last dose of study therapy.

##### 4.4.4 Breast Feeding

Patients must not breastfeed from the first dose and for 1 month following the final dose of nivolumab and capecitabine.

4.4.5 Blood Donation

Patients must not donate blood during the study or for 90 days after the last dose of study treatment.

4.4.6 Immunosuppressive Medication

Immunosuppressive medications including, but not limited to systemic corticosteroids at doses exceeding 10 mg/day of prednisone or equivalent, methotrexate, azathioprine, and TNF-α blockers are not allowed. Use of immunosuppressive medications for the management of investigational product-related AEs or in subjects with contrast allergies is acceptable. In addition, use of inhaled, intranasal, ophthalmic and topical corticosteroids is permitted. A temporary period of steroids will be allowed for different indications, at the discretion of the principal investigator (e.g., chronic obstructive pulmonary disease, radiation, nausea, etc).

#### 4.5 Other Prior and Concomitant Therapy

Any medication or vaccine (including over-the-counter or prescription medicines, vitamins and/or herbal supplements) that the subject is receiving at Screening up to the Final Visit must be recorded in source documents and the case report forms (CRFs). The reason for use, date(s) of administration (including start and end dates), and dosage information (including dose and frequency) must be recorded. Any change in concomitant therapy during the study period must be similarly recorded. Questions regarding prior or concomitant therapy should be directed to one of the investigators.

4.5.1 Prior Surgery

Patients must have fully recovered from all effects of surgery. Patients must have had at least two weeks after minor surgery and four weeks after major surgery before starting therapy. Minor procedures requiring conscious sedation such as endoscopies or mediport placement may only require a 24-hour waiting period, but this must be discussed with an investigator.

4.5.2 Supportive Care

Subjects should receive best supportive care and treatment of symptoms during the study as appropriate, including transfusion of blood and blood products, oxygen therapy, nutritional support, intravenous fluids, and treatment with appropriate medications (antibiotics, antiemetics, antidiarrheals, and analgesics, etc.). Medications that are given for supportive care, such as appetite stimulation, may be given concurrently.

4.5.2.1 Bisphosphonates and denosumab

Bisphosphonates and denosumab are permitted for the treatment of osteopenia or osteoporosis, or as adjuvant treatment to decrease risk of breast cancer recurrence.

4.5.2.2 Hematopoietic growth factors

Hematopoietic growth factors may be used according to the American Society of Clinical Oncology (ASCO) guidelines, but not during the first cycle of the study. The patient must be referred to a hematologist for further evaluation (1) if frequent transfusions are required or (2) if the treatment-related hematologic toxicities have not recovered to CTCAE Grade 1 or less after 4 weeks.

4.5.3 For patients randomized to capecitabine (arms B and C), the approved label of capecitabine should be followed when concomitantly using:

a. Anticoagulants, monitor anticoagulant response (INR or prothrombin time) frequently to adjust the anticoagulant dose as needed.
b. Phenytoin, monitor phenytoin levels in patients.
c. CYP2C9 substrates, caution should be exercised.

#### 4.6 Removal/Replacement of Subjects from Therapy or Assessment

An evaluable patient must meet all inclusion/exclusion criteria, have adequate tissue from definitive breast surgery for initial assessment, and be evaluable for the primary study endpoints of quantification of immune activation and immunomodulation, as well as the safety and tolerability of nivolumab and/or capecitabine.

4.6.1 Screen Failures

Patients will be identified and enrolled after completing definitive breast surgery. All patients must continue to meet the inclusion and exclusion criteria up to and including the first day of treatment. Reasons for patients who have enrolled, but become ineligible could include (but are not limited to):

- The patient is no longer eligible based on laboratory parameters
- The patient's performance status has declined
- The patient does not have adequate tissue for initial assessment

Patients who become ineligible prior to initiation of therapy per protocol will be considered screen failures. Screen failures must be replaced until 45 patients with advanced triple negative breast cancer are enrolled.

4.6.2 Evaluable Patients

4.6.2.1 Immune Response Evaluable

For a patient to be evaluable for immunomodulation by assigned drugs, the patient must be on treatment for 6 weeks (when first assessment of peripheral immunoscore will be conducted). Any patient who is taken off study prior to completion of 2 cycles (=6 weeks) must be replaced.

#### 4.7 Multi-Institutional Trial Coordination

##### 4.7.1 Personnel

At each site, personnel dedicated to this protocol will be:

- A study PI
- A research coordinator
- A data manager

In addition, the Georgetown Multicenter Project management office will play the primary role in coordinating the trial between Lombardi-Georgetown and additional sites. This Multicenter Project Managers will be the main point of contact for Dr. Mainor and the other site PIs for any study related concerns, and to confirm eligibility of each patient being considered for enrollment (Including “remote” confirmation of eligibility for the patients being screened at other sites). The Project Manager(s) will also be the point of contact for the data managers for data entry questions. Finally, the Project managers will play a major role in regulatory coordination of the study, specifically by: 1) Reviewing and confirming all study-related adverse events; 2) Ensuring that sites are submitting all severe adverse event (SAE) reports to the site IRB per local IRB policy ; 3) Gathering and preparing all necessary primary source data for review/audit.

##### 4.7.2 Patient Enrollment

If a patient is being screened for enrollment, the local research coordinator must send an email within 24 hours containing the patient’s initials to the local PI, to Dr. Mainor, and to the Project Managers. If a patient is successfully screened, the local research coordinator must send all supporting documentation to the Project Managers(by secure email). Patients should not start therapy until Dr. Mainor and the Project Managers have reviewed the patient’s records and confirmed that the patient is indeed eligible for enrollment.

##### 4.7.3 Data Collection and Management

Patient data will be entered into the on-line accessible database. This database is housed at Lombardi-Georgetown, but is accessible anywhere there is internet access. The data manager and research coordinator at each site will attend an on-line training session so that they may learn how to enroll data into the database. All screening data should be entered prior to starting therapy, and all ongoing patient data should be entered within 10 business days of each patient visit.

##### 4.7.4 Conference Calls

A biweekly conference call will be held between Lombardi-Georgetown and the other sites to review patient enrollment, toxicity, and response assessment.

4.7.5 Trial Auditing

The multicenter Project Manager will request primary source documents for the patients to be audited. This will include collecting copies of the primary source data for any patients treated at other sites.

#### 5.0 TREATMENT PLAN

If a subject randomizes to Arm A, nivolumab will be provided by the study.

If a subject randomizes to Arm B, medication is not covered by the study and will be handled similarly to standard of care medication administration. Therapy will be based on the patient's actual weight. Dose adjustments will be made according to the approved label and at the discretion of the treating physician.

If a subject randomizes to Arm C, nivolumab will be provided by the study and capecitabine will be covered by patient's insurance.

Administration of nivolumab and/or capecitabine must be initiated within 2 weeks from enrollment in the study arm.

##### 5.1 Dose modifications - General Guidelines

Participants will be monitored continuously for AEs while on study therapy. Participants will be instructed to notify their physician for any and all AEs. Modifications should be applied by the investigator's judgment. In case of an AE relationship assignment to capecitabine alone, dose modifications for capecitabine alone are allowed. In case of doubt, both chemotherapy and nivolumab doses should be modified. Also, in case of assignment of AE relationship to nivolumab alone, dose reduction for chemotherapy is not mandated. Specific algorithms for the management of immune-related AEs are provided in Appendix D and are applicable to immune-related AEs for all immuno-oncology study treatment combinations. For capecitabine chemotherapy, the package insert and local standard of care rules for dose reduction should also be applied.

##### 5.2. Nivolumab

Nivolumab will be given at flat dose of 360 mg iv every 3 weeks for 6 cycles.

5.2.1. Dose Modifications

Immuno-oncology (I-O) agents are associated with AEs that can differ in severity and duration from AEs caused by other therapeutic classes. Early recognition and management of AEs associated with I-O agents may mitigate severe toxicity. Management algorithms have been developed from extensive experience with nivolumab to assist investigators in assessing and managing the following groups of AEs:

- Gastrointestinal
- Renal
- Pulmonary
- Hepatic
- Endocrinopathies
- Skin
- Neurological

Specific algorithms for the management of immune-related AEs are provided in Appendix D and are applicable to immune-related AEs for all immuno-oncology study treatment combinations (Arm A and C).

Nivolumab dose modifications do not apply.

For an AE requiring dose modification, chemotherapy, and/or nivolumab should be interrupted to allow recovery from the AE. Re-initiation of study treatment cannot occur until the AE decreases to ≤ Grade 1 or baseline assessment. In case of delayed recovery to ≤ Grade 1 or baseline from treatment-related AEs that result in a delay of treatment for > 28 days, the participant will not receive additional protocol-related therapy and will be removed from study unless discussed and agreed upon by the Investigator and Sponsor/Medical Monitor that it is in the best interest of the participant to receive additional therapy with nivolumab.

Withhold nivolumab for any of the following:

- Grade 2 pneumonitis
- Grade 2 or 3 colitis
- Aspartate aminotransferase (AST) or alanine aminotransferase (ALT) greater than 3 and up to 5 times upper limit of normal (ULN) or total bilirubin greater than 1.5 and up to 3 times ULN
- Creatinine greater than 1.5 and up to 6 times ULN or greater than 1.5 times baseline
- Any other severe or Grade 3 treatment-related adverse reactions
- Grade 2 hypophysitis
- Grade 2 adrenal insufficiency
- New-onset moderate or severe neurologic signs or symptoms

Resume nivolumab in patients whose adverse reactions recover to Grade 0-1.

Permanently discontinue nivolumab for any of the following:

- Any life-threatening or Grade 4 adverse reaction
- Grade 3 or 4 pneumonitis
- Grade 4 colitis
- AST or ALT greater than 5 times ULN or total bilirubin greater than 3 times ULN
- Creatinine greater than 6 times ULN
- Any severe or Grade 3 treatment-related adverse reaction that recurs
- Inability to reduce corticosteroid dose to 10 mg or less of prednisone or equivalent per day within 12 weeks
- Persistent Grade 2 or 3 treatment-related adverse reactions that do not recover to Grade 0-1 within 12 weeks after last dose of nivolumab
- Grade 3 or 4 adrenal insufficiency
- Immune-mediated encephalitis
- Grade 3 myocarditis

5.2.2. Preparation and Administration

Visually inspect drug product solution for particulate matter and discoloration prior to administration. OPDIVO is a clear to opalescent, colorless to pale-yellow solution. Discard the vial if the solution is cloudy, is discolored, or contains extraneous particulate matter other than a few translucent-to-white, proteinaceous particles. Do not shake the vial.

5.2.3. Preparation

Vials of 100mg will be provided. There are 5 vials per carton. Withdraw the required volume of nivolumab and transfer into an intravenous container. Dilute nivolumab with either 0.9% Sodium Chloride Injection, USP or 5% Dextrose Injection, USP, to prepare an infusion with a final concentration of 1-10 mg/mL, with a maximum total volume of 160 mL for prepared infusions (including overfill). Mix diluted solution by gentle inversion. Do not shake. Discard partially used vials or empty vials of nivolumab.

5.2.4. Storage of Infusion

The product does not contain a preservative.

After preparation, store the nivolumab infusion either:

- at room temperature for no more than 8 hours from the time of preparation. This includes room temperature storage of the infusion in the IV container and time for administration of the infusion or

- under refrigeration at 2°C to 8°C (36°F-46°F) for no more than 24 hours from the time of infusion preparation.

Do not freeze.

5.2.5. Administration

Administer the infusion over 60 minutes through an intravenous line containing a sterile, non pyrogenic, low protein binding in-line filter (pore size of 0.2 micrometer to 1.2 micrometer). Do not coadminister other drugs through the same intravenous line. Flush the intravenous line at end of infusion.

#### 5.3. Capecitabine

Capecitabine will be given at 1250 mg/m2 PO BID for Days 1-14 only of a 21-day cycle for 6 cycles.

Commercially available capecitabine will be used. Locally obtained commercial supplies of capecitabine will be used in accordance with local regulations. As capecitabine is an oral drug available in fixed doses, the dose administered may not exactly match the calculated dose. Determination of the rounding of capecitabine doses for administration should be made according to local institutional practices, with documentation of both the calculated and administered dose. Per package insert, it is recommended that capecitabine be administered with food. Capecitabine should be stored according to the package insert. For capecitabine, the package insert and local standard of care rules for dose reduction should also be applied. Dose modifications will be documented.

5.3.1 Dose modifications for capecitabine for clinical adverse events considered related to capecitabine

| CTCAE Toxicity Grades | During a course of therapy | Dose adjustment for Next Treatment (% of starting dose) |
|---|---|---|
| **Grade 1** | Maintain dose level | Maintain dose level |
| **Grade 2** | | |
| 1st appearance | Interrupt until resolved to Grade ≤ 1 | 100% |
| 2nd appearance | Interrupt until resolved to Grade ≤ 1 | 75% |
| 3rd appearance | Interrupt until resolved to Grade ≤ 1 | 50% |
| 4th appearance | Discontinue permanently | NA |
| **Grade 3** | | |
| 1st appearance | Interrupt until resolved to Grade ≤ 1 | 75% |
| 2nd appearance<br>3rd appearance | Interrupt until resolved to Grade ≤ 1<br>Discontinue permanently | 50%<br>NA |

| Grade 4 | | |
|---|---|---|
| 1st appearance | Discontinue permanently | NA |

### 6.0 STUDY PROCEDURES AND TREATMENT ASSIGNMENT

#### 6.1. Table of events

| | Pre-registration | Screening | On Study (maximum 6 cycles; cycle is every 21 d) | Treatment discontinuation | Surveillance |
|---|---|---|---|---|---|
| | Post-surgery | -30 days | D1 of each cycle +/- 7 days. Each cycle is 21 days. | 30 days (+/-7 days) after last dose | |
| **Assessments** | | | | | |
| Informed consent | X | | | | |
| Medical history and height | | X | | | |
| Physical examination | | X | X^ | X | |
| Weight, BP | | X | X^ | X | |
| ECOG PS | | X | X^ | X | |
| CMP | | X | X | X | |
| Creatinine clearance | | X | X | X | |
| CBC with differential | | X | X | X | |
| Breast imaging (remaining tissue only) | | | | | Yearly +/- 30 days |
| Adverse event and con meds assessment | | X | X^ | | |
| Urine pregnancy or serum HCG for WOCBP only | | X | X^ (every 3 weeks) for Arm A and C | | |
| ACTH and Thyroid Function Tests# | | X | X^ (week 12) for Arm A and C | X | |
| Hepatitis and HIV screening* | | X | | | |
| EORTC QLQ-C30& | | X | X (week 6) | X | |
| Telephone communication or focused physical exam+ | | | | | Every 3-6 months |
| **Treatment** | | | | | |
| | | | Nivolumab 360mg iv q3w; capecitabine 1250mg/$m^2$ D1-14 | | |
| **Correlative Samples** | | | | | |

| | | | | |
|---|---|---|---|---|
| Tumor from initial diagnosis | | X (if available) | | |
| Tumor from definitive surgery – with IHC testing for MSI | X | | | |
| Serum for peripheral immunoscore | | | X^ (baseline, week 6 and 12) | X (if recurrence) |
| Blood for ct-DNA$ | | | X^$ (baseline, week 6 and 12) | X (if recurrence) |
| Tumor from metastatic site | | | | X (if recurrence) |

### Thyroid-stimulating hormone (TSH); free T3 and free T4 at screening and reflex when TSH is abnormal at subsequent visits

* hepatitis B surface antigen, anti-HBc, anti-HBs, hepatitis C antibody (if hep C Ab is positive, reflex to Hep C RNA) or hep C RNA, HIV-1 and -2 antibody (testing for HIV-1 and HIV-2 must be performed at sites where mandated by local requirements)

$ there will be an **optional draw** for ct-DNA 24-48h after the first drug administration

^ +/- 7 days. If a cycle is delayed due to blood counts, no need to repeat physical exam and correlative samples. The correlatives should follow C1D1 as baseline whenever possible and not be delayed in case of delays in treatment.

+ Every 3 months +/- 30 days for the first 2 years, then every 6 months +/- 30 days in year 3

[&] EORTC QLQ-C30 questionnaires to be completed at screening, week 6 and end of treatment

#### 6.2. Pre-registration

Participants who sign informed consent will be pre-registered and tissue from surgery will be requested. Once confirmed availability and sufficient material, patients will undergo screening materials.

#### 6.3. Screening

Within 30 days of study registration:

- Complete medical history and complete physical examination including: height, weight, BP and ECOG performance status
- Complete metabolic profile including serum chemistries and electrolytes
- Calculated creatinine clearance
- Complete blood count with differential
- Urine pregnancy test or serum HCG (only in women of child bearing potential, WOCBP) within 24 hours prior to the start of nivolumab. Women should be counseled regarding acceptable birth control methods to utilize from the time of screening to start treatment. In addition, women of child bearing potential must accept to have a pregnancy test every 3 weeks if they are randomized to a nivolumab arm.
- Adverse event and concomitant medication assessment
- EORTC QLQ-C30 should be completed during screening

- Obtain tumor from definitive breast surgery (mandatory) and from initial biopsy (if tissue is available) and send to the main site (Georgetown). See lab manual for more details.

**Note:** Assessments performed for clinical indications (not exclusively to determine study eligibility) may be used for screening/baseline values even if the studies were done before informed consent was obtained.

**6.4. Randomization**
Eligible patients will be randomized to one of three arms (see 8.3.)

**6.5. On Study Day 1**
Cycle 1 Day 1 testing need not to be repeated if completed within 14 days of starting therapy. Will be obtained blood samples for correlative studies (see Correlative Laboratory Manual for more details). Cycle 1 Day 1 must occur within 2 weeks after receiving randomization results.

**6.6. On Treatment**
Blood samples will be obtained for correlative studies on week 6 and 12 (see Correlative Laboratory Manual for more details).

**6.7. Treatment discontinuation**
A participant will be discontinued from the protocol therapy under the following circumstances:
- if there is evidence of progressive disease
- if the treating physician thinks a chance of therapy would be in the best interest of the participant
- if the participant withdraws consent to protocol treatment
- if the drug exhibits unacceptable adverse events. Participants will be followed until the resolution of these adverse events.
- if a participant becomes pregnant

Patients may temporarily suspend study treatment if they experience an adverse event that requires a dose to be held. If study treatment is held because of adverse events for > 42 days beyond what was planned, then the patient will be discontinued from any study drug and be followed for safety. If, in the judgment of the investigator, the patient is likely to derive clinical benefit from resuming nivolumab and/or capecitabine after a hold > 42 days, study drug may be restarted with the approval of the Medical Monitor.

Dose interruptions for reason(s) other than adverse events, such as surgical procedures, may be allowed with Medical Monitor approval. The acceptable length of interruption will depend on agreement between the investigator and the Medical Monitor.

After treatment discontinuation, in the event of a continuing AE, patients must be followed up until resolution or stabilization of the AE.

#### 6.8. Surveillance

Participants will be monitored by their treating physicians for the development of either local (chest wall, axillary, or supraclavicular nodes) or distant recurrent disease at least once every 3 months for the first two years, then at least every 6 months during year 3. Surveillance will be done through telephone communication, focused physical exam or review of medical records. Surveillance for immune-related adverse events will be included in these follow up visits or telephone calls. The Principal Investigator will notify the Data Safety Monitoring Board of any serious adverse events and immune related adverse events that are believed to be related to prior drug treatment or study procedures that occur at any time after a patient has discontinued treatment.

For those patients that develop a recurrence while on active treatment or during the follow up period (i.e. 3 years), blood correlatives (ctDNA and PIS) and tissue biopsy will be obtained. Tissue for research will only be requested (one block or 13 unstained slides) if a biopsy is done as part of routine care, either to confirm metastatic disease and/or to repeat markers.

### 7.0 SAFETY VARIABLES AND TOXICITY ASSESSMENT

#### 7.1. Serious Adverse Events

A ***Serious Adverse Event (SAE)*** is any untoward medical occurrence that at any dose:

- results in death
- is life-threatening (defined as an event in which the participant was at risk of death at the time of the event; it does not refer to an event which hypothetically might have caused death if it were more severe)
- requires inpatient hospitalization or causes prolongation of existing hospitalization (see **NOTE** below)
- results in persistent or significant disability/incapacity
- is a congenital anomaly/birth defect
- is an important medical event (defined as a medical event(s) that may not be immediately life-threatening or result in death or hospitalization but, based upon appropriate medical and scientific judgment, may jeopardize the subject or may require intervention [eg, medical, surgical] to prevent one of the other serious outcomes listed in the definition above.) Examples of such events include, but are not limited to, intensive treatment in an emergency room or at home for allergic bronchospasm; blood dyscrasias or convulsions that do not result in hospitalization.)

- Suspected transmission of an infectious agent (eg, pathogenic or nonpathogenic) via the study drug is an SAE.

Although pregnancy, overdose, potential drug-induced liver injury (DILI), and cancer are not always serious by regulatory definition, these events must be handled as SAEs.

Any component of a study endpoint that is considered related to study therapy should be reported as an SAE (eg, death is an endpoint, if death occurred due to anaphylaxis, anaphylaxis must be reported).

**NOTE**:

The following hospitalizations are not considered SAEs in BMS clinical studies:

- a visit to the emergency room or other hospital department < 24 hours, that does not result in admission (unless considered an important medical or life-threatening event)
- elective surgery, planned prior to signing consent
- admissions as per protocol for a planned medical/surgical procedure
- routine health assessment requiring admission for baseline/trending of health status (eg, routine colonoscopy)
- Medical/surgical admission other than to remedy ill health and planned prior to entry into the study. Appropriate documentation is required in these cases.
- Admission encountered for another life circumstance that carries no bearing on health status and requires no medical/surgical intervention (eg, lack of housing, economic inadequacy, caregiver respite, family circumstances, administrative reason).
- Admission for administration of anticancer therapy in the absence of any other SAEs (applies to oncology protocols)

All Serious Adverse Events (SAEs) that occur following the subject's written consent to participate in the study through 100* days of discontinuation of dosing must be reported to BMS Worldwide Safety, whether related or not related to study drug. If applicable, SAEs must be collected that relate to any later protocol-specified procedure (eg, a follow-up skin biopsy).

Following the subject's written consent to participate in the study, all SAEs, whether related or not related to study drug, are collected, including those thought to be associated with protocol-specified procedures. The investigator should report any SAE occurring after these aforementioned time periods, which is believed to be related to study drug or protocol-specified procedure.

7.1.1 SAE reporting
An SAE report should be completed for any event where doubt exists regarding its seriousness. If the investigator believes that an SAE is not related to study drug, but is potentially related to the conditions of the study (such as withdrawal of previous therapy or a complication of a study procedure), the relationship should be specified in the narrative section of the SAE Report Form. If the BMS safety address is not included in the protocol document (eg, multicenter studies where events are reported centrally), the procedure for safety reporting must be reviewed/approved by the BMS Protocol Manager. Procedures for such reporting must be reviewed and approved by BMS prior to study activation.

An appropriate SAE form (USA = MedWatch form) should be used to report SAEs to BMS. The BMS protocol ID number must be included on whatever form is submitted by the Sponsor/Investigator.

For studies with long-term follow-up periods in which safety data are being reported, include the timing of SAE collection.

The Sponsor will reconcile the clinical database SAE cases (case level only) transmitted to BMS Global Pharmacovigilance. Frequency of reconciliation should be every 3 months and prior to the database lock or final data summary. BMS GPV&E will email, upon request from the Investigator, the GPV&E reconciliation report. Requests for reconciliation should be sent to. The data elements listed on the GPV&E reconciliation report will be used for case identification purposes. If the Investigator determines a case was not transmitted to BMS GPV&E, the case should be sent immediately to BMS

The MedWatch form is available at:
https://www.fda.gov/downloads/AboutFDA/ReportsManualsForms/Forms/UCM048334.pdf

In accordance with local regulations, BMS will notify investigators of all reported SAEs that are suspected (related to the investigational product) and unexpected (i.e., not previously described in the IB). An event meeting these criteria is termed a Suspected, Unexpected Serious Adverse Reaction (SUSAR). Investigator notification of these events will be in the form of a SUSAR Report.

SAEs, whether related or not related to study drug, and pregnancies must be reported to BMS within 24 hours. SAEs must be recorded on either CIOMS or MedWatch form & pregnancies must be reported on a Pregnancy Surveillance Form or can be submitted on the aforementioned SAE form to BMS.

Other important findings which may be reported by BMS as an Expedited Safety Report (ESR) include: increased frequency of a clinically significant expected SAE, an SAE considered associated with study procedures that could modify the conduct of the study, lack of efficacy that poses significant hazard to study subjects, clinically significant safety finding from a nonclinical (eg, animal) study, important safety recommendations from a study data monitoring committee, or sponsor decision to end or temporarily halt a clinical study for safety reasons.

Upon receiving an ESR from BMS, the investigator must review and retain the ESR with the IB. Where required by local regulations or when there is a central IRB/IEC for the study, the sponsor will submit the ESR to the appropriate IRB/IEC. The investigator and IRB/IEC will determine if the informed consent requires revision. The investigator should also comply with the IRB/IEC procedures for reporting any other safety information.

In addition to the Sponsor Investigator's responsibility to report events to their local HA, suspected serious adverse reactions (whether expected or unexpected) shall be reported by BMS to the relevant competent health authorities in all concerned countries according to local regulations (either as expedited and/or in aggregate reports).

**SAE Facsimile Number:** +1 609-818-3804

If only limited information is initially available, follow-up reports are required. (Note: Follow-up SAE reports should include the same investigator term(s) initially reported.)
If an ongoing SAE changes in its intensity or relationship to study drug or if new information becomes available, a follow-up SAE report should be sent within 24 hours to BMS (or designee) using the same procedure used for transmitting the initial SAE report.
All SAEs should be followed to resolution or stabilization.

**SAE Reporting for Subsites:** Subsites should report all SAEs to Georgetown using the MedWatch form within 24 hours. Georgetown will then forward SAE reports to BMS Safety within 24 hours through email or fax.

#### 7.2. Adverse Events

An Adverse Event (AE) is defined as any new untoward medical occurrence or worsening of a preexisting medical condition in a clinical investigation participant administered study drug and that does not necessarily have a causal relationship with this treatment. An AE can therefore be any unfavorable and unintended sign (such as an abnormal laboratory finding), symptom, or disease temporally associated with the use of investigational product, whether or not considered related to the investigational product.

The causal relationship to study drug is determined by a physician and should be used to assess all adverse events (AE). The casual relationship can be one of the following:

a. Related: There is a reasonable causal relationship between study drug administration and the AE.
b. Not related: There is not a reasonable causal relationship between study drug administration and the AE.

The term "reasonable causal relationship" means there is evidence to suggest a causal relationship.
Adverse events can be spontaneously reported or elicited during open-ended questioning, examination, or evaluation of a subject. (In order to prevent reporting bias, subjects should not be questioned regarding the specific occurrence of one or more AEs.)

7.2.1. Nonserious Adverse Events
Non-serious Adverse Events (AE) are to be provided to BMS in aggregate via interim or final study reports as specified in the agreement or, if a regulatory requirement [eg, IND US trial] as part of an annual reporting requirement.

- Non-serious AE information should also be collected from the start of a placebo lead-in period or other observational period intended to establish a baseline status for the subjects.

A ***non-serious adverse event*** is an AE not classified as serious.

7.2.1.1. Non-serious Adverse Event Collection and Reporting
The collection of non-serious AE information should begin at initiation of study drug. All non-serious adverse events (not only those deemed to be treatment-related) should be collected continuously during the treatment period and for a minimum of 30 days following the last dose of study treatment.

Non-serious AEs should be followed to resolution or stabilization, or reported as SAEs if they become serious. Follow-up is also required for non-serious AEs that cause interruption or discontinuation of study drug and for those present at the end of study treatment as appropriate.

#### 7.3. Adverse Events Associated with the Use of Immune Checkpoint Inhibitor
The immune-related AEs listed below have been reported as related to the use of Nivolumab. Each subject who receives Nivolumab (arms A and C) will be closely monitored by the Investigator and managed, as appropriate, per the respective product labels. In addition, all

immune-related AEs will be evaluated by the Safety Review Committee throughout the conduct of this study.

Subjects in treatment Arms A and C will be monitored for:

- Pneumonitis
- Colitis
- Hepatitis
- Hypophysitis
- Thyroid disorders
- Adrenal insufficiency
- Diabetes mellitus
- Nephritis and renal dysfunction
- Rash
- Encephalitis

#### 7.4. Laboratory Test Abnormalities

All laboratory test results captured as part of the study should be recorded following institutional procedures. Test results that constitute SAEs should be documented and reported to BMS as such.

The following laboratory abnormalities should be documented and reported appropriately:

- any laboratory test result that is clinically significant or meets the definition of an SAE
- any laboratory abnormality that required the participant to have study drug discontinued or interrupted
- any laboratory abnormality that required the subject to receive specific corrective therapy.

It is expected that wherever possible, the clinical rather than laboratory term would be used by the reporting investigator (eg, anemia versus low hemoglobin value).

##### 7.4.1. Potential Drug Induced Liver Injury (DILI)

Specific criteria for identifying potential DILI have not been identified for this protocol. Standard medical practice in identifying and monitoring hepatic issues should be followed.
Wherever possible, timely confirmation of initial liver-related laboratory abnormalities should occur prior to the reporting of a potential DILI event. All occurrences of potential DILIs, meeting the defined criteria, must be reported as SAEs.

Potential drug induced liver injury is defined as:

1) AT (ALT or AST) elevation > 3 times upper limit of normal (ULN)

**AND**

2) Total bilirubin > 2 times ULN, without initial findings of cholestasis (elevated serum alkaline phosphatase)

**AND**

3) No other immediately apparent possible causes of AT elevation and hyperbilirubinemia, including, but not limited to, viral hepatitis, pre-existing chronic or acute liver disease, or the administration of other drug(s) known to be hepatotoxic.

Wherever possible, timely confirmation of initial liver-related laboratory abnormalities should occur prior to the reporting of a potential DILI event. All occurrences of potential DILIs, meeting the defined criteria, must be reported as SAEs.

#### 7.5. Pregnancy

If, following initiation of the investigational product, it is subsequently discovered that a study participant is pregnant or may have been pregnant at the time of investigational product exposure, including during at least 5 half-lives after product administration, the investigational product will be permanently discontinued in an appropriate manner (eg, dose tapering if necessary for participant).

The investigator must immediately notify of this event via either the CIOMS, MedWatch or appropriate Pregnancy Surveillance Form in accordance with SAE reporting procedures. Protocol-required procedures for study discontinuation and follow-up must be performed on the participant. Follow-up information regarding the course of the pregnancy, including perinatal and neonatal outcome and, where applicable, offspring information must be reported on the CIOMS, MedWatch or appropriate Pregnancy Surveillance Form. A BMS Pregnancy Surveillance Form may be provided upon request. Any pregnancy that occurs in a female partner of a male study participant should be reported to BMS. Information on this pregnancy will be collected on the Pregnancy Surveillance Form. In order for Sponsor or designee to collect any pregnancy surveillance information from the female partner, the female partner must sign an informed consent form for disclosure of this information.

#### 7.6. Overdose

An overdose is defined as the accidental or intentional administration of any dose of a product that is considered both excessive and medically important. All occurrences of overdose must be reported as an SAE.

#### 7.7. Other Safety Considerations

Any significant worsening noted during interim or final physical examinations, electrocardiograms, X-rays, and any other potential safety assessments, whether or not these procedures are required by the protocol, should also be recorded as a non-serious or serious AE, as appropriate, and reported accordingly.

#### 7.8. Study Monitoring

##### 7.8.1 Data Safety Monitoring Committee at Georgetown

The Georgetown Lombardi Comprehensive Cancer Center (LCCC) will be responsible for the data and safety monitoring of this trial. As this study is an investigator initiated study utilizing FDA-approved on label, and off label therapies, it is considered a high-risk study which requires real-time monitoring by the PI and study team and quarterly reviews by the LCCC Data and Safety Monitoring Committee (DSMC).

The Principal Investigator, Dr. Mainor, and the Study Chair will review the data including safety monitoring at their monthly teleconferences with participating sites.

All SAEs are required to be reported to Dr. Mainor and the Multicenter Project Manager(s). In additions, SAEs should be reported to the local and/or to the Georgetown IRB per local IRB policy. Based on SAEs, the IRB retains the authority to suspend further accrual pending more detailed reporting and/or modifications to further reduce risk and maximize the safety of participating patients.

Progress on the trial and the toxicities experienced will be reviewed by the LCCC Data and Safety Monitoring Committee every 3 months from the time the first patient is enrolled on the study. Results of the DSMC meetings will be forwarded to the IRB with recommendations regarding need for study closure. DSMC recommendations should be based not only on results for the trial being monitored as well as on data available to the DSMC from other studies. It is the responsibility of the PI to ensure that the DSMC is kept apprised of non-confidential results from related studies that become available. It is the responsibility of the DSMC to determine the extent to which this information is relevant to its decisions related to the specific trial being monitored.

A written copy of the DSMC recommendations will be given to the trial PI and the IRB. If the DSMC recommends a study change for patient safety or efficacy reasons the trial PI must act to implement the change as expeditiously as possible. In the unlikely event that the trial PI does not concur with the DSMC recommendations, then the LCCC Associate Director of Clinical Research must be informed of the reason for the disagreement. The trial PI, DSMC Chair, and the LCCC Associate Director for Clinical Research will be responsible for reaching a mutually acceptable decision about the study and providing details of that decision to the IRB. Confidentiality must be preserved during these discussions. However, in some cases, relevant data may be shared with other selected trial investigators and staff to seek advice to assist in reaching a mutually acceptable decision. If a recommendation is made to change a trial for reasons other than patient safety or efficacy the DSMC will provide an adequate rationale for its decision. If the DSMC recommends that the trial be closed for any reason, the recommendation will be reviewed by the Associate Director for Clinical Research at LCCC. Authority to close a trial for safety reasons lies with the IRB, with the above described input from DSMC and the Associate Director for Clinical Research.

Of note, the DSMC will also review the safety data of the patients enrolled outside of Georgetown University. The Multicenter Project Manager will be tasked with the job of requesting and collecting primary source documentation for patients enrolled outside of Georgetown University. In addition, the data managers at each site will be entering data into the Georgetown database, so that all data will be available for the DSMC at Georgetown to review. Records should be sent to the Multicenter Project Manager(s) and Dr. Mainor via email.

##### 7.9 Protocol Deviations and Violations

Protocol deviations (including violations) are cases of drug administration, radiotherapy, clinical laboratory tests, toxicity evaluation, efficacy evaluation, etc., not being carried out in accordance with the study protocol. There are three categories of deviation, as detailed below.

###### 7.9.1 Violations

In general, violations are deviations from the study protocol that meet any one of the following criteria:

1. The deviation affects evaluation of the study endpoints
2. The party responsible for the deviation is a responsible physician and/or medical institution
3. The deviation was deliberate and/or long-term
4. The deviation presented risks for the patients and/or was major
5. As a result of the violation, the procedure followed was inappropriate for clinical use

Violations must be described in detail in all published reports.

###### 7.9.2 Deviations

This refers to deviations that belong to neither the “violation” nor “acceptable deviation” categories. If a particular deviation occurs numerous times, it is preferable for it to be included in published reports.

###### 7.8.3. Acceptable Deviations

Acceptable deviations deviate from the protocol within a range that has been agreed upon, either beforehand or after the fact, by the study group and the Administrative Office.

#### 8.0. STATISTICAL CONSIDERATIONS

##### 8.1 Objectives

Primary objective:

2) To assess the immunologic effects of capecitabine, nivolumab or the combo, post neoadjuvant and post-surgery, in the adjuvant setting, in patients with high risk TNBC as defined by presence of residual disease (breast and or LNs) at surgery, in terms of the change of PIS.
   a) We hypothesized that among patients with TNBC and residual disease at the time of surgery, the change of the Peripheral ImmunoScore (PIS) from baseline to week 6 will be higher among those who receive post-surgery immunotherapy (ARM A and C), compared to those who receive post-surgery chemotherapy alone (ARM B).

Secondary objectives:

1). To determine the incidence of toxicity graded using the National Cancer Institute CTCAE v. 4.0 until 30 days after last dose of treatment on trial.
2). To determine association between changes in PIS from baseline to week 6 and week 12 and clinical outcome variables (DRFS and OS at 3-years). After end of study visit, clinical follow up or telephone communication every 3months.
3). Translational endpoints:
   a) To assess changes in
   b) circulating tumor DNA (ct-DNA) during treatment

Exploratory objectives:

   a) To describe the immune contexture in residual tumors of patients with high risk TNBC after receiving neoadjuvant chemotherapy
   b) Tissue banking for future studies (NGS of initial biopsy and surgical specimen), plasma, PBMCs

#### 8.2 Sample Size Considerations

The primary objective of the study is to assess the change in PIS, the Peripheral ImmunoScore, at 6 weeks (=2 cycles) from baseline, in each arm.

The sample size of 15 per arm (45 totally for 3 arms) is mainly based on the feasibility of patient accrual. A sample size of 15 per arm will have about 85% power to detect an effect size of 1 (the difference of the change in PIS from baseline to week 6 between two arms divided by the standard deviation) at 5% significance level.

#### 8.3. Randomization and Stratification Factors

Stratified randomization will be used to assign patients into the three arms. There will be three strata: nivolumab (arm A), capecitabine (arm B) and nivolumab/capecitabine combo (arm C). Within each stratum, blocked randomization with randomly selected block sizes will be used. The stratified randomization procedure will be carried out by the biostatistician(s) at the LCCC Biostatistics and Bioinformatics Shared Resource. They will implement the randomization, such as generate the randomization table and hold the randomization key.

**8.4 Endpoints**

8.4.1. Primary Endpoint

a. Quantification of immune activation measured by changes of PIS from baseline to week 6 in each arm.

8.4.2 Secondary Endpoints

a. Quantification of immune activation measured by changes of PIS from baseline to week 12 in each arm.
b. Quantification of grade 3 and 4 toxicities according to the National Cancer Institute Common Toxicity Criteria for Adverse Events Version 4.0 [NCI CTCAE v4.03]
c. Association between changes in PIS from baseline to week 6 and week 12 and clinical outcome variables (DRFS and OS at 3-years). Distant recurrence free survival (DRFS) is defined by time from study enrollment to date of first invasive distant disease recurrence, second invasive primary cancer (breast or not), or death due to any cause.
d. Quantification of ct-DNA at different time points

8.4.3 Exploratory Endpoints

e. Quantification of immune activation by IHC, flow cytometry and ELISA analysis
f. Quantification of antigen-specific responses by intracellular cytokine staining and CD8+ T-cell clonal expansion

**8.5 Analysis Plan**

8.5.1. Analysis for the Primary Endpoint

For the primary endpoint, the observed change of PIS from baseline to week 6 in each arm will be presented with its 95% confidence interval. The difference of the change of PIS between treatment arms will be compared using two-sample t-test.

For a patient to be evaluable for immunomodulation, measured by the peripheral immunoscore, by nivolumab, capecitabine or the combo, the patient must complete the first 2 cycles. Any patient who is taken off study prior to completion of 2 cycles must be replaced.

8.5.2. Analysis for the Secondary Endpoint

The observed incidence of toxicity will be tabulated according to the grade level and the attribution using the National Cancer Institute CTCAE v. 4.0. Fisher's Exact test will be used to compare the incidence rate of toxicity between treatment groups. We expect that among patients with TNBC and residual disease at the time of surgery, those who receive post-surgery immunotherapy (ARM A and C) will have longer invasive disease-free survival (DRFS) time compared to those who receive post-surgery chemotherapy alone (ARM B). Patients will be followed for DRFS and overall survival (OS) for 3 years. The Kaplan-Meier method will be used to analyze the DRFS and OS. The DRFS and OS at year 3 will be presented with their 95% confidence intervals. Log-rank test will be used to compare the DRFS and OS between treatment groups.

#### 9.0 CORRELATIVE RESEARCH

##### 9.1 Peripheral Immunoscore

We selected the refined PIS[15] as a surrogate of clinical benefit and a new strategy to identify patients with TNBC and high-risk disease who may benefit from immunotherapy to eliminate residual micrometastasis. The peripheral immunoscore was calculated based on a predefined analyses of refined immune cell subsets for which a biologic function had been previously reported revealed statistically significant improved PFS ($P < 0.001$) in patients with metastatic breast cancer receiving docetaxel plus immunotherapy compared to docetaxel alone (Fig 2). To calculate the PIS, flow cytometry was performed on PBMC harvested at different time points. A multiparametric flow cytometry platform was applied to measure the frequency of PBMC. Patients' PBMCs by density gradient separation, and then 1 mL of PBMCs was cryopreserved in liquid nitrogen at a concentration of 107 cells/mL per vial. On the day of staining, one vial of cryopreserved PBMCs per patient was defrosted, cells were counted, and then stained with the appropriate monoclonal antibodies. Table 1 shows the classic immune cell types as well as those refined subsets with a phenotype reflecting immune function. Viability of all samples following thawing after cryopreservation was 80% to 95%. The frequency of individual subsets was calculated as a percentage of total PBMCs to help reduce the bias that could occur in the smaller subpopulations with fluctuations in parental leukocyte populations. The PIS defined by Farsaci et al was selected based on being the closest to the particular scenario proposed on the OXEL study. This has been validated in a group of patients with metastatic BC receiving docetaxel and immunotherapy, and shown to be predictive of PFS. While this (and other) immune biomarkers in the blood available today have not been validated in all clinical scenarios – including early BC – available data suggests that this immunoscore constitutes a promising surrogate marker of clinical outcome in this group of patients. The PIS will be performed 40) at Dr. Jeffrey Schlom's lab at NIH, which developed this particular immunoscore.

##### 9.2 Immune contexture in residual tumors

We will evaluate PD-L1 expression in the tumor, expression of other immune checkpoints that may be upregulated (e.g., LAG3, TIM3, ICOS) and expression of the cellular composition (CD8+, activated T cells, CD4+, T regs, MDSC and macrophages) using the Perkin Elmer Vectra 3.0 Automated Quantitative Pathology Imaging System, which is housed in the Lombardi Histopathology and Tissue Shared Resource (HTSR). We will also measure other cytokines and chemokines involved in the immune system activation by Nanostring, as described in Table 2. Based on preclinical studies of immune regulation of tumor growth[13] we will specifically examine CD47, TGFF□1, Sgpl1, Tex9 and Pex14 in the tumor specimens by PCR and IHC. All tissues will be processed in Dr. Louis Weiner's laboratory and assays will be performed in the Lombardi HTSR (IHC) and the Lombardi Genomics and Epigenomics Shared Resource (Nanostring, PCR).

| | Prioritized for Analysis |
|---|---|
| Immunologic profile (Vectra 3.0 IHC) | PDL1, LAG3, TIM3, ICOS, other checkpoints |
| Cellular and molecular composition of residual tumor (Vectra 3.0) | CD8+, CD4+, Tregs, MDSC, PD1, other checkpoints |
| Other cytokines and chemokines (Nanostring) | IL-2, 4, 6, 8, 10, 17; IFN-b, g; TNF-a; STING; |
| Other candidate resistance biomarkers (qPCR, Vectra 3.0 IHC) | CD47, TGFβ1, Sgpl1, Tex9 and Pex14 |

Table 2. Immune Biomarkers to be Studied

#### 9.3. Sample Labeling Overview

Patients will be de-identified and labeled with the following study label (XXX-XXXX-XX-OXEL):

- The first three letters will be the site name.
  - GUH = Georgetown
  - WHC = Washington Hospital Center
  - HCK = Hackensack
  - UOC = Univ of Chicago
  - UAB = Univ Alabama at Birmingham
- The four numbers will refer to the patient number preceded by the site number
  - 1 = Georgetown
  - 2 = Washington Hospital Center
  - 3 = Hackensack
  - 4 = Univ of Chicago
  - 5 = Univ Alabama at Birmingham
- This will be followed by 2 letters that will be the 2 patient initials (first name, last name)
- Finally, this will be followed by the study name OXEL

- Each sample (tissue) will be labeled with the procedure date in MM/DD/YR format and will have a number to indicate the type of collection. Options include:
  - Initial biopsy (tissue) - 1
  - Surgery (tissue) - 2
  - Recurrence (tissue) - 3

See Correlative Laboratory Manual for more details regarding sample handling and processing.

#### 10.0 ETHICAL CONSIDERATIONS

##### 10.1 Institutional Review Board (IRB)

GCP requires that the clinical protocol, any protocol amendments, the Investigator's Brochure, the informed consent and all other forms of subject information related to the study (e.g., advertisements used to recruit subjects) and any other necessary documents be reviewed by an IRB. IRB approval of the protocol, informed consent and subject information and/or advertising, as relevant, will be obtained prior to the authorization of drug shipment to a study site. Any amendments to the protocol will require IRB approval prior to implementation of any changes made to the study design. Any serious adverse events that meet the reporting criteria, as dictated by local regulations, will be reported to the IRB. During the conduct of the study, the investigator should promptly provide written reports to the IRB of any changes that affect the conduct of the study and/or increase the risk to subjects.

##### 10.2 Ethical Conduct of the Study

The study will be conducted in accordance with the protocol, ICH guidelines, applicable regulations and guidelines governing clinical study conduct and the ethical principles that have their origin in the Declaration of Helsinki.

##### 10.3 Subject Information and Consent

Prior to the initiation of any screening or study-specific procedures, the investigator or his/her representative will explain the nature of the study to the subject and answer all questions regarding this study. Each informed consent will be reviewed, signed and dated by the subject and the person who administered the informed consent. A copy of each informed consent will be given to the subject and each original will be placed in the subject's medical record. An entry must also be made in the subject's dated source documents to confirm that informed consent was obtained prior to any study-related procedures and that the subject received a signed copy. Tissue sample collection for analysis will only be performed if the subject has voluntarily consented to participate after the nature of the testing has been explained and the subject has had the opportunity to ask questions.

##### 10.4 Ethical Consideration for Enrollment

Only patients with triple negative breast cancer and residual disease at the time of surgery, who have a high risk of recurrence, will be considered for enrollment. As described above, nivolumab alone or in combination with capecitabine is a rational and promising combination for such patients.

#### 10.5 Protection of Patient Confidentiality

All patient records, questionnaires, and tissue specimens will be de-identified using a letter and number assigned to their case at the time of enrollment on study. No record or specimen will contain information which could identify the patient. The key which connects patient identifiable information with this assigned number will be held by the Principal investigator. For computer records, the key will be protected by a double password protection system. Any paper records will be contained in a locked cabinet within a locked office to ensure patient's privacy is protected.

#### 12.0 APPENDICES

#### APPENDIX A

##### Table 1: ECOG Performance Status Scale

| Grade | ECOG |
|---|---|
| 0 | Fully Active, able to carry on all pre-disease performance without restriction. |
| 1 | Restricted in physically strenuous activity but ambulatory and able to carry out work of a light or sedentary nature, e.g., light housework, office work. |
| 2 | Ambulatory and capable of all self-care but unable to carry out any work activities. Up and about more than 50% of waking hours. |
| 3 | Capable of only limited self care, confined to bed or chair more than 50% of waking hours. |
| 4 | Completely disabled. Cannot carry on any self-care. Totally confined to bed or chair. |

**APPENDIX B: PATIENT REGISTRATION FORM**

OXEL: A pilot study of immune checkpoint or capecitabine or combination therapy as adjuvant therapy for triple negative breast cancer with residual disease following neoadjuvant chemotherapy

Study ID: _________

Instructions: This form should be completed by the research staff before registering the patient into the trial.

□ Georgetown University Medical Center

□ Other Institution ___________________________________________

□ Treating Physician ____________ ____________________________

1. Date Informed Consent signed: ____/____/___

2. Screening Date: ____/____/___

3. Start date for treatment: ____/____/_______

4. Prior therapies (date initiated/type):

_______________________________________________________________

_______________________________________________________________

_______________________________________________________________

_______________________________________________________________

_______________________________________________________________

5. Please fax the following documentation:
   - □ Pathology Report
   - □ Physicians Note validating:
   - □ Previous treatments
   - □ Laboratory Results
   - □ Past Medical History

**APPENDIX C: Data Sharing Plan**

Data sharing for this study will be conducted in compliance with the February 26, 2003 NIH Statement on Data Sharing (NOTICE: NOT-OD-03-032). The collaborating sites for this study generate a wide variety of scientific and clinical data and Data Sharing and Archiving will be handled by in accordance with NIH Statement on Data sharing, institutional internal document retention policies and all application rules, regulations and statutes.

Subject to institutional policies, local IRB guidelines, and local, state and Federal laws and regulations including the Privacy Rule and the Bayh-Dole Act, we will make finished research data available through scientific presentations, publications (paper, web and other), depositing gene sequence, gene expression and other data in searchable electronic repositories, attendance at scientific meetings and extending invitations to scientists from other institutions for discussion. In accordance with the NIH policy, such data shall be made widely and freely available while safeguarding the privacy of participants and protecting Lombardi's confidential and proprietary data.

The participating sites will maintain awareness, and may participate in, discussions between members of multiple scientific and technical disciplines and their professional societies concerning data sharing, standards and best practices, and to create an environment that supports and develops data sharing tools. We will participate in or make ourselves aware of the outcome of any workshops the NIH or AACR will convene to address data sharing and which may address areas such as cleaning and formatting data, writing documentation, redacting data to protect subjects' identities and proprietary information, and estimating costs to prepare documentation and data for sharing.

The NIH has recognized the need to protect patentable and other proprietary data and notes the restrictions on the sharing of data that may be imposed by agreements with third parties. Under the Bayh-Dole Act, grantees have the right to elect and retain title to subject inventions developed with Federal funding, and further, to commercialize any invention to which they retain title. Since its is not the stated intent of the NIH statement on Data Sharing to discourage, impede or prohibit the development of commercial products from federally funded research, our collaborating sites will continue to perform inventive activities, to seek patent protection for inventions that relate to data generated and may choose to defer publication or enter into agreements with third parties that may result in certain restrictions on data sharing. We note that seeking patent protection results in publication of the patent application into the public domain and, thus, may result in the data being broadly disseminated.

All specimens submitted to any of the shared resource laboratories will be bar-coded upon receipt and assigned a unique identifier using a common software program. Tracking of specimens and their utilization will be carried out using a web-based system that will represent a modification of the systems presently in place at Georgetown (including but not limited to G-

DOC, Georgetown Database of Cancer, Medidata Rave, REDCap), which will also give us the ability to track sample utilization. These systems have an "honest broker" interface which ensures HIPPA compliance and protection of any human subjects. As the same sample may well be used for genomic, proteomic, histopathological and clinical interrogation, assignment of a unique identifier as well as a common administrative structure ensures efficient cross-database interrogation.

**Data security**

The rights and privacy of people who participate in sponsored clinical research will be protected at all times. In the event that data is intended for broader use, it will be de-identified and would not permit linkages to individual research participants and variables that could lead to deductive disclosure of the identity of individual subjects. No efforts will be made to identify individual cases, and any shared archive data will not be linked to other identifiable data. The following de-identification and security procedures will be followed to share information with collaborators part of the study:

1) Deletion of 18 HIPAA identifiers
    a. Non-identifiable unique patient ID will be generated in G-DOC
2) Secure netID-based single sign-on (netID is Georgetown's LDAP based secure login system)
3) Users will have to *authenticate* themselves prior to accessing controlled data
4) Furthermore, based on their roles, users will require *authorization* to see specific studies
5) Auditing and security assessments will be performed on a quarterly basis to ensure appropriate de-identification procedures and use of data.

For future studies involving new data types that are not covered in the descriptions above, NIH policy on data sharing will be followed where applicable. For example, Genome Wide Association Studies, if conducted, will comply with *NIH Guidelines* NOT-OD-07-088 (http://grants.nih.gov/grants/gwas/) for data release. Following these guidelines, GWAS data will be submitted to NCBI's dbGAP (http://www.ncbi.nlm.nih.gov/sites/entrez?db=gap) or other tools as the NIH's policy on GWAS evolves.

**Data Disclosure**

GUMC (Georgetown University Medical Center) has a Confidentiality and Non-Disclosure policy that pertains only to proprietary information belonging to GUMC. The disclosure of research information such as microarray analysis results is at the discretion of the faculty and staff. Notably, investigators with NIH funding are expected to make their data and results public in a reasonable time frame (Please see the University plan for sharing and distributing biomedical research resources at http://otc.georgetown.edu/documents/inventors/NIHGrantLetter_ModelOrganismsUpdate_6-27-05.doc . To protect institutional intellectual property, the institution does have an internal review process prior to submission of journal manuscripts. This process will be propagated to other study sites as well.

#### APPENDIX D: MANAGEMENT ALGORITHMS

These general guidelines constitute guidance to the Investigator and may be supplemented by discussions with the Medical Monitor representing the Sponsor. The guidance applies to all immuno-oncology agents and regimens.

A general principle is that differential diagnoses should be diligently evaluated according to standard medical practice. Non-inflammatory etiologies should be considered and appropriately treated.

Corticosteroids are a primary therapy for immuno-oncology drug-related adverse events. The oral equivalent of the recommended IV doses may be considered for ambulatory patients with low grade toxicity. The lower bioavailability of oral corticosteroids should be taken into account when switching to the equivalent dose of oral corticosteroids.

Consultation with a medical or surgical specialist, especially prior to an invasive diagnostic or therapeutic procedure, is recommended.

The frequency and severity of the related adverse events covered by these algorithms will depend on the immuno-oncology agent or regimen being used.

### GI Adverse Event - Management Algorithm

Rule out non-inflammatory causes. If non-inflammatory cause is identified, treat accordingly and continue I-O therapy. Opiates/narcotics may mask symptoms of perforation. Infliximab should not be used in cases of perforation or sepsis.

| Grade of Diarrhea/ Colitis (NCI CTCAE v4) | Management | Follow-up |
|---|---|---|
| **Grade 1**<br>Diarrhea:< 4 stools/day over baseline;<br>Colitis: asymptomatic | • Continue I-O therapy per protocol<br>• Symptomatic treatment | • Close monitoring for worsening symptoms.<br>• Educate patient to report worsening immediately<br>If worsens:<br>• Treat as Grade 2 or 3/4 |
| **Grade 2**<br>Diarrhea: 4-6 stools per day over baseline; IV fluids indicated <24 hrs; not interfering with ADL<br>Colitis: abdominal pain; blood in stool | • Delay I-O therapy per protocol<br>• Symptomatic treatment | If improves to grade 1:<br>• Resume I-O therapy per protocol<br>If persists > 5-7 days or recur:<br>• 0.5-1.0 mg/kg/day methylprednisolone or oral equivalent<br>• When symptoms improve to grade 1, taper steroids over at least 1 month, consider prophylactic antibiotics for opportunistic infections, and resume I-O therapy per protocol.<br>If worsens or persists > 3-5 days with oral steroids:<br>• Treat as grade 3/4 |
| **Grade 3-4**<br>Diarrhea (G3): ?7 stools per day over baseline; incontinence; IV fluids ?24 hrs; interfering with ADL<br>Colitis (G3): severe abdominal pain, medical intervention indicated, peritoneal signs<br>G4: life-threatening, perforation | • Discontinue I-O therapy per protocol<br>• 1.0 to 2.0 mg/kg/day methylprednisolone IV or IV equivalent<br>• Add prophylactic antibiotics for opportunistic infections<br>• Consider lower endoscopy | If improves:<br>• Continue steroids until grade 1, then taper over at least 1 month<br>If persists > 3-5 days, or recurs after improvement:<br>• Add infliximab 5 mg/kg (if no contraindication). Note: Infliximab should not be used in cases of perforation or sepsis |

Patients on IV steroids may be switched to an equivalent dose of oral corticosteroids (e.g. prednisone) at start of tapering or earlier, once sustained clinical improvement is observed. Lower bioavailability of oral corticosteroids should be taken into account when switching to the equivalent dose of oral corticosteroids.

### Renal Adverse Event - Management Algorithm

Rule out non-inflammatory causes. If non-inflammatory cause, treat accordingly and continue I-O therapy

| Grade of Creatinine Elevation (NCI CTCAE v4) | Management | Follow-up |
|---|---|---|
| **Grade 1**<br>Creatinine > ULN and > than baseline but s 1.5x baseline | • Continue I-O therapy per protocol<br>• Monitor creatinine weekly | If returns to baseline:<br>•Resume routine creatinine monitoring per protocol<br>If worsens:<br>•Treat as Grade 2 or 3/4 |
| **Grade 2-3**<br>Creatinine > 1.5x baseline to s 6x ULN | • Delay I-O therapy per protocol<br>• Monitor creatinine every 2-3 days<br>• 0.5 to 1.0 mg/kg/day methylprednisolone IV or oral equivalent<br>• Consider renal biopsy with nephrology consult | If returns to Grade 1:<br>•Taper steroids over at least 1 month, consider prophylactic antibiotics for opportunistic infections, and resume I-O therapy and routine creatinine monitoring per protocol<br><br>If elevations persist > 7 days or worsen:<br>•Treat as Grade 4 |
| **Grade 4**<br>Creatinine > 6x ULN | • Discontinue I-O therapy per protocol<br>• Monitor creatinine daily<br>• 1.0-2.0 mg/kg/day methylprednisolone IV or IV equivalent<br>• Consult nephrologist<br>• Consider renal biopsy | If returns to Grade 1 : Taper steroids over at least 1 month and add prophylactic antibiotics for opportunistic infections |

Patients on IV steroids may be switched to an equivalent dose of oral corticosteroids (e.g. prednisone) at start of tapering or earlier, once sustained clinical improvement is observed. Lower bioavailability of oral corticosteroids should be taken into account when switching to the equivalent dose of oral corticosteroids

### Pulmonary Adverse Event - Management Algorithm

Rule out non-inflammatory causes. If non-inflammatory cause, treat accordingly and continue I-O therapy. Evaluate with imaging and pulmonary consultation.

| Grade of Pneumonitis (NCI CTCAE v4) | Management | Follow-up |
|---|---|---|
| **Grade 1**<br>Radiographic changes only | • Consider delay of I-O therapy<br>• Monitor for symptoms every 2-3 days<br>• Consider Pulmonary and ID consults | • Re-image at least every 3 weeks<br>If worsens:<br>• Treat as Grade 2 or 3-4 |
| **Grade 2**<br>Mild to moderate new symptoms | • Delay I-O therapy per protocol<br>• Pulmonary and ID consults<br>• Monitor symptoms daily, consider hospitalization<br>• 1.0 mg/kg/day methyl-prednisolone IV or oral equivalent<br>• Consider bronchoscopy , lung biopsy | • Re-image every 1-3 days<br>If improves:<br>• When symptoms return to near baseline, taper steroids over at least 1 month and then resume I-O therapy per protocol and consider prophylactic antibiotics<br>If not improving after 2 weeks or worsening:<br>• Treat as Grade 3-4 |
| **Grade 3-4**<br>Severe new symptoms; New/worsening hypoxia; Life-threatening | • Discontinue I-O therapy per protocol<br>• Hospitalize<br>• Pulmonary and ID consults<br>• 2-4 mg/kg/day methylprednisolone IV or IV equivalent<br>• Add prophylactic antibiotics for opportunistic infections<br>• Consider bronchoscopy, lung biopsy | If improves to baseline:<br>• Taper steroids over at least 6 weeks<br>If not improving after 48 hours or worsening:<br>• Add additional immunosuppression |

Patients on IV steroids may be switched to an equivalent dose of oral corticosteroids (e.g. prednisone) at start of tapering or earlier, once sustained clinical improvement is observed. Lower bioavailability of oral corticosteroids should be taken into account when switching to the equivalent dose of oral corticosteroids.

### Hepatic Adverse Event - Management Algorithm

Rule out non-inflammatory causes. If non-inflammatory cause, treat accordingly and continue I-O therapy. Consider imaging for obstruction.

| Grade of Liver Test Elevation (NCI CTCAE v4) | Management | Follow-up |
|---|---|---|
| **Grade 1**<br>AST or ALT > ULN to 3.0 x ULN <u>and/or</u> T. bili > ULN to 1.5 x ULN | • Continue I-O therapy per protocol | • Continue LFT monitoring per protocol<br><u>If worsens:</u><br>• Treat as Grade 2 or 3-4 |
| **Grade 2**<br>AST or ALT > 3.0 to s 5 x ULN <u>and/or</u> T. bili > 1.5 to s 3 x ULN | • Delay I-O therapy per protocol<br>• Increase frequency of monitoring to every 3 days | <u>If returns to baseline</u>:<br>•Resume routine monitoring, resume I-O therapy per protocol<br><br><u>If elevations persist > 5-7 days or worsen</u> :<br>• 0.5-1 mg/kg/day methylprednisolone or oral equivalent and when LFT returns to grade 1 or baseline, taper steroids over at least 1 month, consider prophylactic antibiotics for opportunistic infections, and resume I-O therapy per protocol |
| **Grade 3-4**<br>AST or ALT > 5 x ULN <u>or</u> T.bili >3 x ULN | • Discontinue I-O therapy*<br>• Increase frequency of monitoring to every 1-2 days<br>• 1.0 to 2.0 mg/kg/day methylprednisolone IV or IV equivalent**<br>• Add prophylactic antibiotics for opportunistic infections<br>• Consult gastroenterologist | <u>If returns to grade 2</u>:<br>• Taper steroids over at least 1 month<br><br><u>If does not improve in >3-5 days, worsens or rebounds</u>:<br>• Add mycophenolate mofetil 1 g BID<br>• If no response within an additional 3-5 days, consider other immunosuppressants per local guidelines |

Patients on IV steroids may be switched to an equivalent dose of oral corticosteroids (e.g. prednisone) at start of tapering or earlier, once sustained clinical improvement is observed. Lower bioavailability of oral corticosteroids should be taken into account when switching to the equivalent dose of oral corticosteroids.

***I-O therapy may be delayed rather than discontinued if AST/ALTs 8 x ULN or T.bilis 5 x ULN.**

**The recommended starting dose for grade 4 hepatitis is 2 mg/kg/day methylprednisolone IV.

### Endocrinopathy - Management Algorithm

Rule out non-inflammatory causes. If non-inflammatory cause, treat accordingly and continue I-O therapy. Consider visual field testing, endocrinology consultation and imaging.

**Asymptomatic TSH elevation** →

- Continue I-O therapy per protocol
- If TSH < 0.5 x LLN, or TSH > 2 x ULN, or consistently out of range in 2 subsequent measurements: include fT4 at subsequent cycles as clinically indicated; consider endocrinology consult

**Symptomatic endocrinopathy** →

- Evaluate endocrine function
- Consider pituitary scan

Symptomatic with abnormal lab/pituitary scan:
- Delay I-O therapy per protocol
- 1-2 mg/kg/day methylprednisolone IV or PO equivalent
- Initiate appropriate hormone therapy

No abnormal lab/pituitary MRI scan but symptoms persist:
- Repeat labs in 1-3 weeks / MRI in 1 month

→ If improves (with or without hormone replacement):
- Taper steroids over at least 1 month and consider prophylactic antibiotics for opportunistic infections
- Resume I-O therapy per protocol
- Patients with adrenal insufficiency may need to continue steroids with mineralocorticoid component

**Suspicion of adrenal crisis (e.g. severe dehydration, hypotension, shock out of proportion to current illness** →

- Delay or discontinue I-O therapy per protocol
- Rule out sepsis
- Stress dose of IV steroids with mineralocorticoid activity
- IV fluids
- Consult endocrinologist
- If adrenal crisis ruled out, then treat as above for symptomatic endocrinopathy

Patients on IV steroids may be switched to an equivalent dose of oral corticosteroids (e.g. prednisone) at start of tapering or earlier, once sustained clinical improvement is observed. Lower bioavailability of oral corticosteroids should be taken into account when switching to the equivalent dose of oral corticosteroids.

### Skin Adverse Event - Management Algorithm

Rule out non-inflammatory causes. If non-inflammatory cause, treat accordingly and continue I-O therapy.

| Grade of Rash (NCI CTCAE v4) | Management | Follow-up |
| --- | --- | --- |
| **Grade 1-2**<br>Covering s 30% BSA* | • Symptomatic therapy (e.g. antihistamines, topical steroids)<br>• Continue I-O therapy per protocol | <u>If persists > 1-2 weeks or recurs:</u><br>• Consider skin biopsy<br>• Delay I-O therapy per protocol<br>• Consider 0.5-1.0 mg/kg/day methylprednisolone IV or oral equivalent. Once improving , taper steroids over at least 1 month, consider prophylactic antibiotics for opportunistic infections, and resume I-O therapy per protocol<br><u>If worsens:</u><br>• Treat as Grade 3-4 |
| **Grade 3-4**<br>Covering >30% BSA; Life threatening consequences*^ | • Delay or discontinue I-O therapy per protocol<br>• Consider skin biopsy<br>• Dermatology consult<br>• 1.0-2.0 mg/kg/day IV methylprednisolone IV or IV equivalent | <u>If improves to Grade 1:</u><br>• Taper steroids over at least 1 month and add prophylactic antibiotics for opportunistic infections<br>• Resume I-O therapy per protocol |

Patients on IV steroids may be switched to an equivalent dose of oral corticosteroids (e.g. prednisone) at start of tapering or earlier, once sustained clinical improvement is observed. Lower bioavailability of oral corticosteroids should be taken into account when switching to the equivalent dose of oral corticosteroids.

*Refer to NCI CTCAE v4 for term-specific grading criteria.

^If SJS/TEN is suspected, withhold I-O therapy and refer patient for specialized care for assessment and treatment. If SJS or TEN is diagnosed, permanently discontinue I-O therapy.

### Neurological Adverse Event Management Algorithm

Rule out non-inflammatory causes. If non-inflammatory cause, treat accordingly and continue I-O therapy.

| Grade of Neurological Toxicity (NCI CTCAE v4) | Management | Follow-up |
|---|---|---|
| **Grade 1**<br>Asymptomatic or mild symptoms; Intervention not indicated | • Continue I-O therapy per protocol | Continue to monitor the patient.<br>If worsens:<br>• Treat as Grade 2 or 3-4 |
| **Grade 2**<br>Moderate symptoms; Limiting instrumental ADL | • Delay I-O therapy per protocol<br>• Treat symptoms per local guidelines<br>• Consider 0.5 to 1.0 mg/kg/day methylprednisolone IV or PO equivalent | If improves to baseline:<br>•Resume I-O therapy per protocol when improved to baseline<br>If worsens:<br>• Treat as Grade 3-4 |
| **Grade 3-4**<br>Severe symptoms; Limiting self-care ADL; Life-threatening | • Discontinue I-O therapy per protocol<br>• Obtain neurology consult<br>• Treat symptoms per local guidelines<br>• 1.0-2.0 mg/kg/day IV methylprednisolone IV or IV equivalent<br>• Add prophylactic antibiotics for opportunistic infections | If improves to Grade 2:<br>• Taper steroids over at least 1 month<br><br>If worsens or atypical presentation:<br>• Consider IVIG or other immunosuppressive therapies per local guidelines |

Patients on IV steroids may be switched to an equivalent dose of oral corticosteroids (e.g. prednisone) at start of tapering or earlier, once sustained clinical improvement is observed. Lower bioavailability of oral corticosteroids should be taken into account when switching to the equivalent dose of oral corticosteroids

#### APPENDIX E: Quality of Life Questionnaire (EORTC QLQ-C30)

### EORTC QLQ-C30 (version 3)

We are interested in some things about you and your health. Please answer all of the questions yourself by circling the number that best applies to you. There are no "right" or "wrong" answers. The information that you provide will remain strictly confidential.

Please fill in your initials: □□□□
Your birthdate (Day, Month, Year): □□□□□□□□
Today's date (Day, Month, Year): 31 □□□□□□□□

---

| | Not at All | A Little | Quite a Bit | Very Much |
|---|---|---|---|---|
| 1. Do you have any trouble doing strenuous activities, like carrying a heavy shopping bag or a suitcase? | 1 | 2 | 3 | 4 |
| 2. Do you have any trouble taking a long walk? | 1 | 2 | 3 | 4 |
| 3. Do you have any trouble taking a short walk outside of the house? | 1 | 2 | 3 | 4 |
| 4. Do you need to stay in bed or a chair during the day? | 1 | 2 | 3 | 4 |
| 5. Do you need help with eating, dressing, washing yourself or using the toilet? | 1 | 2 | 3 | 4 |

| **During the past week:** | Not at All | A Little | Quite a Bit | Very Much |
|---|---|---|---|---|
| 6. Were you limited in doing either your work or other daily activities? | 1 | 2 | 3 | 4 |
| 7. Were you limited in pursuing your hobbies or other leisure time activities? | 1 | 2 | 3 | 4 |
| 8. Were you short of breath? | 1 | 2 | 3 | 4 |
| 9. Have you had pain? | 1 | 2 | 3 | 4 |
| 10. Did you need to rest? | 1 | 2 | 3 | 4 |
| 11. Have you had trouble sleeping? | 1 | 2 | 3 | 4 |
| 12. Have you felt weak? | 1 | 2 | 3 | 4 |
| 13. Have you lacked appetite? | 1 | 2 | 3 | 4 |
| 14. Have you felt nauseated? | 1 | 2 | 3 | 4 |
| 15. Have you vomited? | 1 | 2 | 3 | 4 |
| 16. Have you been constipated? | 1 | 2 | 3 | 4 |

Please go on to the next page

| **During the past week:** | **Not at All** | **A Little** | **Quite a Bit** | **Very Much** |
|---|---|---|---|---|
| 17. Have you had diarrhea? | 1 | 2 | 3 | 4 |
| 18. Were you tired? | 1 | 2 | 3 | 4 |
| 19. Did pain interfere with your daily activities? | 1 | 2 | 3 | 4 |
| 20. Have you had difficulty in concentrating on things, like reading a newspaper or watching television? | 1 | 2 | 3 | 4 |
| 21. Did you feel tense? | 1 | 2 | 3 | 4 |
| 22. Did you worry? | 1 | 2 | 3 | 4 |
| 23. Did you feel irritable? | 1 | 2 | 3 | 4 |
| 24. Did you feel depressed? | 1 | 2 | 3 | 4 |
| 25. Have you had difficulty remembering things? | 1 | 2 | 3 | 4 |
| 26. Has your physical condition or medical treatment interfered with your family life? | 1 | 2 | 3 | 4 |
| 27. Has your physical condition or medical treatment interfered with your social activities? | 1 | 2 | 3 | 4 |
| 28. Has your physical condition or medical treatment caused you financial difficulties? | 1 | 2 | 3 | 4 |

**For the following questions please circle the number between 1 and 7 that best applies to you**

29. How would you rate your overall health during the past week?

1 2 3 4 5 6 7

Very poor Excellent

30. How would you rate your overall quality of life during the past week?

1 2 3 4 5 6 7

Very poor Excellent

© Copyright 1995 EORTC Quality of Life Group. All rights reserved. Version 3.0
